# Comprehensive genetic diagnosis of tandem repeat expansion disorders with programmable targeted nanopore sequencing

**DOI:** 10.1101/2021.09.27.21263187

**Authors:** Igor Stevanovski, Sanjog R. Chintalaphani, Hasindu Gamaarachchi, James M. Ferguson, Sandy S. Pineda, Carolin K. Scriba, Michel Tchan, Victor Fung, Karl Ng, Andrea Cortese, Henry Houlden, Carol Dobson-Stone, Lauren Fitzpatrick, Glenda Halliday, Gianina Ravenscroft, Mark R. Davis, Nigel G. Laing, Avi Fellner, Marina Kennerson, Kishore R. Kumar, Ira W. Deveson

## Abstract

Short-tandem repeat (STR) expansions are an important class of pathogenic genetic variants. Over forty neurological and neuromuscular diseases are caused by STR expansions, with 37 different genes implicated to date. Here we describe the use of programmable targeted long-read sequencing with Oxford Nanopore’s ReadUntil function for parallel genotyping of all known neuropathogenic STRs in a single, simple assay. Our approach enables accurate, haplotype-resolved assembly and DNA methylation profiling of expanded and non-expanded STR sites. In doing so, the assay correctly diagnoses all individuals in a cohort of patients (*n* = 27) with various neurogenetic diseases, including Huntington’s disease, fragile X syndrome and cerebellar ataxia (CANVAS) and others. Targeted long-read sequencing solves large and complex STR expansions that confound established molecular tests and short-read sequencing, and identifies non-canonical STR motif conformations and internal sequence interruptions. Even in our relatively small cohort, we observe a wide diversity of STR alleles of known and unknown pathogenicity, suggesting that long-read sequencing will redefine the genetic landscape of STR expansion disorders. Finally, we show how the flexible inclusion of pharmacogenomics (PGx) genes as secondary ReadUntil targets can identify clinically actionable PGx genotypes to further inform patient care, at no extra cost. Our study addresses the need for improved techniques for genetic diagnosis of STR expansion disorders and illustrates the broad utility of programmable long-read sequencing for clinical genomics.

**One sentence summary:** This study describes the development and validation of a programmable targeted nanopore sequencing assay for parallel genetic diagnosis of all known pathogenic short-tandem repeats (STRs) in a single, simple test.

## INTRODUCTION

A short tandem repeat (STR) is a short DNA sequence motif, typically 2-6 bp, repeated consecutively at a given position in the genome. STRs make up ∼7% of the human genome sequence and are highly polymorphic, commonly varying in length between unrelated individuals *(1, 2)*.

Unusually long or ‘expanded’ STR alleles are an important class of pathogenic variants in human populations. To date, STR expansions in over forty genes have been shown to cause heritable disorders, with the majority of these exhibiting primary neurological or neuromuscular presentations *(3, 4)*. These include Huntington’s disease (*HTT*), fragile X syndrome (*FMR1*), the hereditary cerebellar ataxias (*RFC1, FXN* and others), the myotonic dystrophies (*DMPK, CNBP*), the myoclonic epilepsies (*CSTB, SAMD12, STARD7* and others), and *C9orf72*-related frontotemporal dementia and amyotrophic lateral sclerosis *(3, 4)*. With each of the > 40 STR-associated neurogenetic diseases estimated to affect ∼1-10 individuals per 100,000, their collective prevalence is high *(5-8)*. Moreover, the list of disorders in which STR expansions are implicated continues to grow and many new pathogenic STR genes have been described recently *(9-13)*.

Given: (*i*) the wide variety and collectively high prevalence of STR expansion disorders; (*ii*) the large number of genes involved; (*iii*) the frequent identification of new genes; (*iv*) the diversity in size and sequence conformation of pathogenic STR expansion alleles and; (*v*) the many gaps in our understanding of their basic biology; there is a growing need for improved methods for molecular characterisation of STRs. Established molecular techniques (e.g. Southern blot and repeat-primed PCR) are relatively slow, labour-intensive, imprecise and require a separate assay with specific primers/probes for every different STR *(14)*. This is problematic when multiple different STR expansions can manifest in a similar phenotype (locus heterogeneity) *(3, 4)* and is a major barrier to the implementation of tests for newly identified STR genes. Next-generation sequencing (NGS) has some utility for analysis of STR expansions *(15-19)*. However, the large size, low sequence-complexity and high GC-content of many pathogenic STR expansions make them refractory to analysis by short-read NGS platforms (e.g., Illumina) *(14)*.

Long-read sequencing technologies from Oxford Nanopore Technologies (ONT) and Pacific Biosciences (PacBio) can be used to genotype large and complex STR expansions *(12, 20-23)*. However, whole-genome analysis remains prohibitively expensive on either platform. A cas9-based approach for targeted enrichment of STR loci and long-read sequencing was recently developed *(20, 24)*. However, this suffers from the same limitation as existing molecular techniques, in that a unique set of cas9 guide-RNAs are needed for every different STR, requiring careful design and experimental optimisation.

An alternative approach to targeted long-read sequencing is ONT’s ‘ReadUntil’ functionality, whereby an ONT sequencing device can be programmed to recognise and accept/reject specific DNA sequence fragments during a sequencing experiment *(25, 26)*. Target selection is fully flexible and requires no additional laboratory processes beyond standard library preparation. Here we demonstrate that ONT ReadUntil can be used to achieve accurate molecular characterisation of all known neuropathogenic STRs in a single assay. Using a custom panel comprising 37 STR loci associated with neurological and neuromuscular disease, we performed targeted ONT sequencing on 27 patient-derived DNA samples to identify and fully characterise a diverse range of STR expansions. Our study establishes the analytical validity of programmable ONT sequencing for the genetic diagnosis of STR expansion disorders and showcases the numerous advantages of this approach.

## RESULTS

### Programmable targeted nanopore sequencing of pathogenic STRs

ONT’s ReadUntil function has the potential to enable simple, cost-effective sequencing of all known pathogenic STR loci but is a largely untested technology. To evaluate the use of ReadUntil for STR profiling, we designed a custom panel encompassing all genes known to harbour pathogenic STR expansions implicated in primary neurological and neuromuscular diseases (*n* = 37; **Supplementary Table 1**). For each gene, the entire locus was targeted, including 50 kb of flanking sequence in either direction (**Figure 1A**). The panel included a range of additional clinically informative loci and covered ∼50.5 Mbase in total, equating to ∼1.6% of the human reference genome (hg38; see **Supplementary Table 2**).

**Figure 1.**
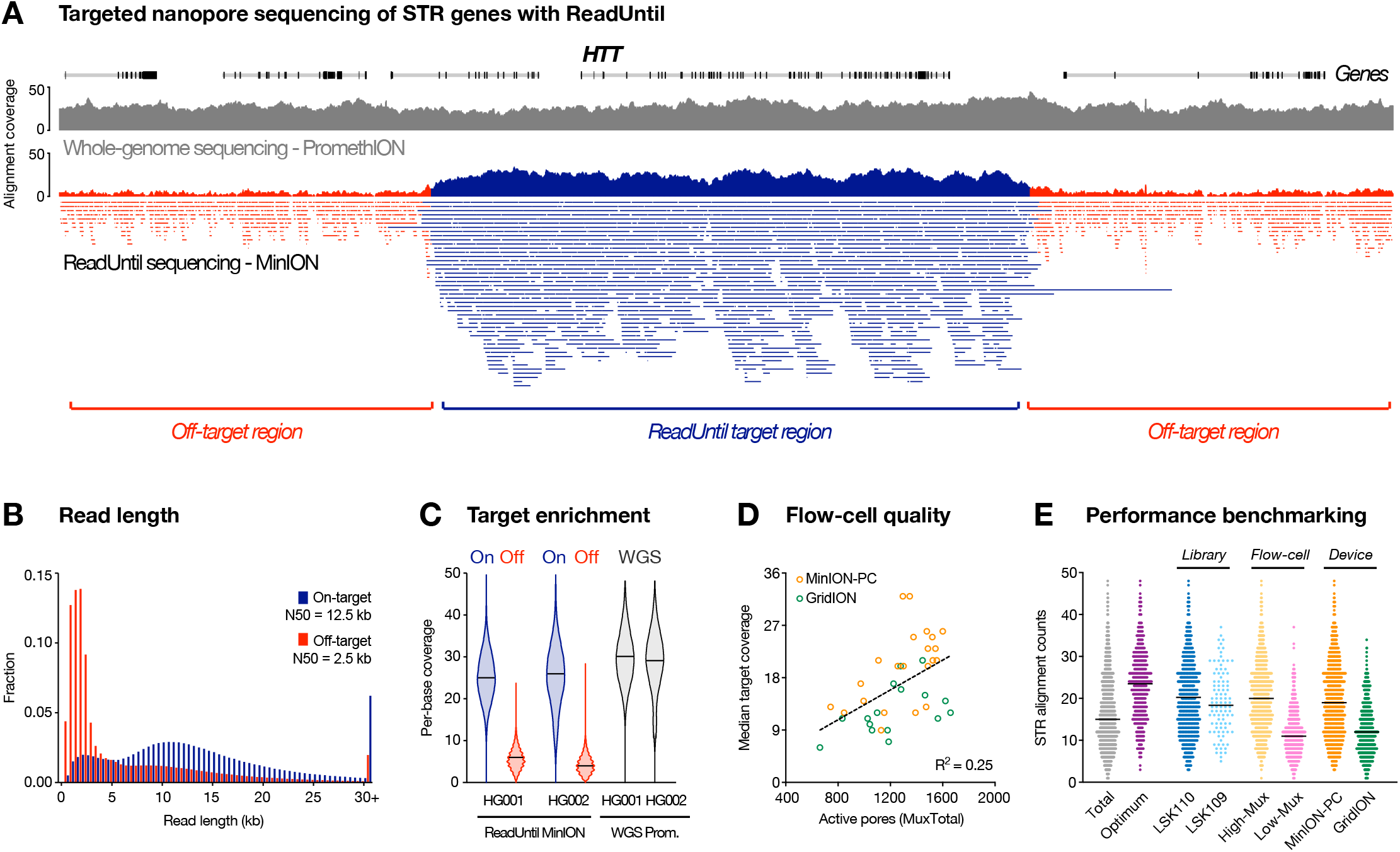
Targeted sequencing of pathogenic STR sites with ONT ReadUntil. (**A**) Genome browser view shows sequencing alignments to the *HTT* locus and surrounding regions for a typical ONT ReadUntil experiment (lower track). Location of ReadUntil target region for *HTT* is marked below and on-target (navy) vs off-target alignments (red) are distinguished by colour. For comparison, a coverage track is also shown for a typical whole-genome ONT sequencing experiment (grey). (**B**) Histograms compare read-length distribution for on-target (navy; N50 = 12.5kb) vs off-target (red; N50 = 2.5kb) alignments. Data is averaged over all ReadUntil experiments from the study (n = 38 runs). (**C**) Violin plots show per-base coverage distributions within on-target regions (navy) vs randomly selected off-target genes (red) during ReadUntil sequencing of HG001 and HG002 reference samples, with data from whole-genome sequencing (WGS; grey) shown for comparison. (**D**) Scatter plot shows median coverage across on-target regions, relative to the starting number of active pores (MuxTotal) on each ONT flow-cell. Colours distinguish ReadUntil experiments run on an ONT GridION (green; NVIDIA Quadro GV100 GPU; n = 16) or a high-spec desktop PC (orange; NVIDIA 3090 GPU; n = 22); see **Supplementary Table 5** for full specs. (**E**) Dot plots show the number of alignments spanning STR sites (n = 37) across all ONT ReadUntil experiments (n = 38). Colours distinguish runs performed with LSK110 (navy) vs LSK109 (blue) library preparation kit, high quality (MuxTotal >1,200 pores; yellow) vs low-quality (MuxTotal <1,200 pores; pink) and GridION (green) vs MinION-PC device (orange). Data from runs with ‘optimum’ parameters (LSK110 + MuxTotal >1,200 pores + MinION-PC device; purple) yielded a median 24 alignments spanning target STR sites.

We used the open-source software package *Readfish (26)*, to execute targeted sequencing on 27 genomic DNA samples obtained from reference catalogs or collected from consenting patients (**Supplementary Table 3**). Genomic DNA was sheared to ∼15-25 kb fragments before library preparation and sequenced on an ONT MinION flow-cell (see **Methods**). We observed a consistent reduction in read-length for off-target reads (N50 = 2.5kb) compared to on-target reads (N50 = 12.5kb), indicating successful rejection of fragments outside the target regions (**Figure 1B**). This resulted in a median 4.5-fold enrichment in sequencing depth within target regions, yielding ∼9-32X median target coverage across the cohort (**Figure 1C**; **Supplementary Table 4**). Notably, ONT ReadUntil achieved similar coverage depth and evenness to whole-genome ONT sequencing of matched samples on a high-output PromethION flow-cell, performed at more than three times the cost (**Figure 1C**).

To determine optimum workflow settings, we evaluated the impact of various parameters, including the choice of ONT library preparation kit, flow-cell quality and the computer used to execute ReadUntil (see **Methods**). This revealed: (*i*) superior on/off-target enrichment with LSK109 library prep chemistry, but greater total output and on-target coverage with LSK110 (**Fig. S1A**,**B**); (*ii*) the initial number of live pores on a flow-cell is an important determinant of final target coverage achieved (**Figure 1D**; **Fig. S1C**,**D**); (*iii*) shorter average read-rejection time and improved target coverage when running *Readfish* on a powerful desktop PC, compared to an ONT GridION device, thanks to its superior Graphics Processing Unit (GPU; **Figure 1D**; **Fig. S1E**,**F**; **Supplementary Table 5**). Across all sequencing experiments with optimum workflow parameters (LSK110, MinION-PC, >1200 pores) we obtained a median of 24 sequence alignments spanning targeted STR sites (**Figure 1E**), demonstrating that ONT ReadUntil can be used to achieve effective targeted STR sequencing.

### Validity & utility of programmable targeted STR sequencing

To establish the validity and utility of our targeted nanopore sequencing assay, we analysed a range of patient-derived reference DNA samples and consenting patients with neurogenetic diseases, including Huntington’s disease (HD), fragile X syndrome (FXS), cerebellar ataxia, neuropathy, and vestibular areflexia syndrome (CANVAS), spinal and bulbar muscular atrophy of Kennedy (SBMA), myotonic dystrophy 1 (DM1), neuronal intranuclear inclusion disease (NIID), Friedreich ataxia (FRDA) and amyotrophic lateral sclerosis (ALS), as well as unaffected individuals. Samples were also subject to independent molecular testing in accredited genetic pathology laboratories, or using standard approaches (see **Methods**). This allowed ONT sequencing data to be evaluated against current best-practices.

#### Huntington’s disease (HD)

HD is an autosomal dominant neurodegenerative disorder caused by a poly-glutamine STR expansion of ≥ 36 ‘CAG’ motifs within the gene *HTT*, with complete penetrance at ≥ 40 copies *(27, 28)*. STR expansion size is correlated with disease severity, as is the absence of characteristic ‘CAA’ interrupting motifs (also coding for glutamine) within expanded alleles *(29, 30)*. Genetic diagnosis of HD therefore requires accurate, allele-specific STR sizing and internal sequence determination.

Targeted sequencing with ReadUntil yielded a median of 18 spanning alignments at the STR site in *HTT* exon 1 (**Fig. S2Ai**). This was sufficient to phase and assemble both STR alleles in every patient, identifying ‘CAG’ repeats ranging from 12 to 71 copies across the cohort (**Figure 2A**; **Fig. S2Ai**; see **Methods**). In all patients affected by HD (*n* = 5), a single expanded STR allele was detected with length in the known pathogenic range, whereas no pathogenic expansions were detected in unaffected individuals (*n* = 22). The lengths of both expanded and non-expanded STR alleles determined by ONT sequencing were closely concordant with clinical testing (R^2^ = 0.996; **Figure 2B**). A single ‘CAA’ interruption was detected within both STR alleles of all HD-affected and non-affected individuals, with six of the latter individuals harbouring a double ‘CAA’ interruption (**Figure 2A**; **Fig. S2Ai**). Our assay also clearly resolved the boundary between the disease-associated ‘CAG’ poly-glutamine repeat and a ‘CCG’ poly-proline repeat located immediately downstream within *HTT* exon1, which similarly varied in size across our cohort. While polymorphism in this adjacent repeat is not considered relevant to the HD phenotype, it is an important technical variable that can confound molecular assays for sizing the disease-associated STR *(31)*.

**Figure 2.**
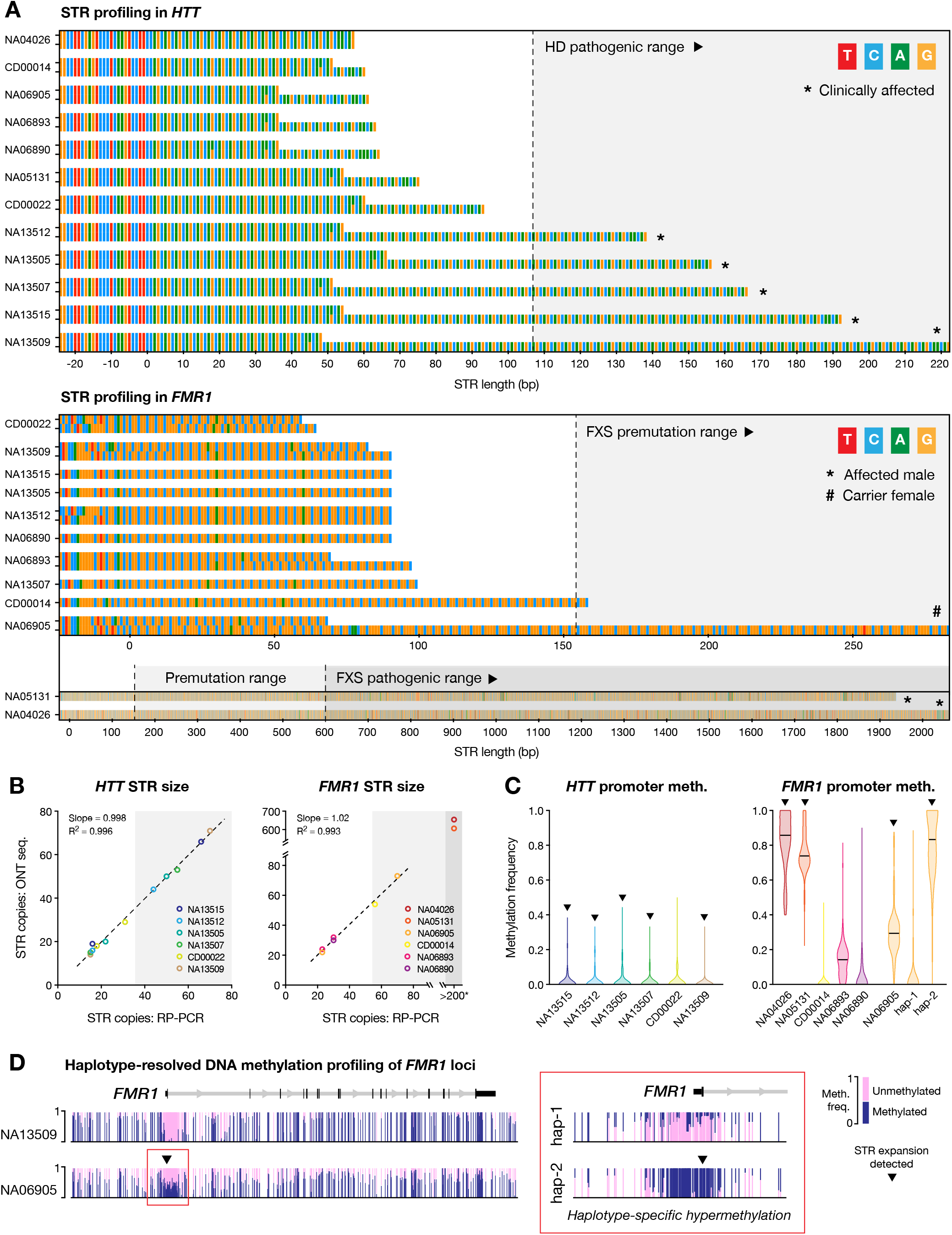
Haplotype-resolved assembly and DNA methylation profiling of *HTT* & *FMR1*. (**A**) Sequence-barcharts show *HTT* (upper) and *FMR1* (lower) STR alleles, including 25 bp of upstream flanking sequence, assembled from ONT sequencing of relevant Coriell reference DNA samples (n = 12; see **Supplementary Table 3**). Two alleles are shown for each individual, excepting *FMR1* for male individuals, where only one copy is present. Clinically affected and premutation-carrier individuals are marked, based on clinical information from Corriell. Further individuals are shown in **Fig. S2**. (**B**) Scatter plots show lengths of STR alleles in *HTT* (CAGn; left) and *FMR1* (CGGn; right) as determined by ONT sequencing vs repeat-primed PCR (data from Coriell). For *FMR1*, two samples exceeded the upper limit of RP-PCR genotyping (∼CGG_200_). (**C**) For the same samples, violin plots show distribution of DNA methylation frequencies recorded at CpG sites within the promoter regions of *HTT* (left) and *FMR1* (right). Triangles indicate which samples contained pathogenic STR expansions. For sample NA06905, differential methylation was observed between the two *FMR1* haplotypes; these are shown separately. (**D**) Genome browser view shows examples of DNA methylation profiles across the complete *FMR1* locus for two samples: NA13509 (female with no STR expansion in *FMR1*) and NA06905 (female carrier of *FMR1* premutation). Inset shows haplotype-specific promoter methylation in NA06905.

HD-like syndromes may be caused by other STR genes that were also included on our targeted sequencing panel (i.e., *C9orf72, PRNP, JPH3, TBP, ATXN8, FXN* and *ATN1*) *(3, 4)*. Parallel sequencing of these genes showed that all HD-affected patients harboured STR alleles within healthy ranges (**Fig. S2**). The capacity to rule out confounding or co-occurring STR expansions in these genes, without the need for additional molecular tests, is a clear advantage of our multi-gene assay.

#### Fragile X syndrome (FXS)

FXS is the most common cause of inherited intellectual disability and single-gene cause of autism spectrum disorder in males *(32)*. FXS is caused by large (>200) ‘CGG’ STR expansions within the chrX-linked gene *FMR1 (33)*. Premutation alleles of 55-200 ‘CGG’ repeats are also associated with late-onset fragile-X-associated tremor/ataxia syndrome (FXTAS) in males and primary ovarian insufficiency in females *(5)*. Interrupting ‘AGG’ motifs are reported to stabilise STR alleles to protect against full expansion *(34, 35)*. DNA methylation (5mC) is also implicated in the pathogenic mechanism of *FMR1*-related disorders, with expanded alleles typically exhibiting promoter-hypermethylation and silencing of *FMR1 (36, 37)*. Therefore, complete genetic diagnosis of *FMR1*-related disorders requires DNA methylation profiling, in addition to STR sizing and internal sequence determination.

At the STR site in *FMR1* exon 1, we obtained a median of 19 spanning alignments in females and 9 in males, or ∼9 alignments per allele overall (**Fig. S2Bi**). All alleles were successfully assembled into ‘CGG’ STRs ranging from 20 to 654 copies across the cohort (**Figure 2A**; **Fig. S2Bi**). The length of both expanded and non-expanded alleles was closely concordant with clinical testing (R^2^ = 0.993; **Figure 2B**). Results from ONT sequencing correctly distinguished affected male individuals (*n* = 2) and a female carrier (*n* = 1) from unaffected individuals, as well as distinguishing pre-mutation alleles (*n* = 2) from full pathogenic STR expansions (*n* = 2; **Figure 2A**). Within both individuals that harboured a pre-mutation allele, we detected two protective ‘AGG’ motif interruptions and similar interruptions were common across non-expanded STR alleles in unaffected individuals (**Figure 2A**; **Fig. S2Bi**). Parallel sequencing of *FMR2* (*AFF2*), an STR expansion gene with partial phenotypic overlap to *FMR1 (38)*, ruled out pathogenic expansions in all individuals (**Fig. S2**).

DNA methylation profiling revealed hypermethylation of the *FMR1* promoter region in both FXS-affected males, who harboured full STR expansions (CGG_654_ & CGG_606_), with > 75% median methylation frequencies among local CpG sites (**Figure 2C**; see **Methods**). By contrast, promoter CpG methylation frequencies were low among males with normal and pre-mutation STR alleles (< 25% median freq.; **Figure 2C**). Females also showed low methylation frequencies, with the exception of the single pre-mutation carrier (NA0695). In this individual we observed differential methylation between the two *FMR1* haplotypes, with the pre-mutation haplotype being predominantly methylated (CGG_73_; 83% median freq.) but the normal haplotype unmethylated (GCC_23_; 0% median freq.; **Figure 2C,D**). DNA hypermethylation in *FMR1* premutation alleles has been reported previously, is correlated to STR repeat size, and may account for variability in phenotypic expression in premutation carriers *(39, 40)*. In contrast to *FMR1*, and in line with expectations, we did not observe DNA hypermethylation in the promoter of *HTT* on either expanded or non-expanded STR alleles for any individuals, further confirming the reliability of the analysis (**Figure 2C**). The capacity to obtain haplotype-resolved DNA methylation profiles, in addition to STR size and interruption status, all in a single, simple assay is a clear advantage of our approach.

#### Cerebellar ataxia, neuropathy and vestibular areflexia syndrome (CANVAS)

CANVAS is a neurodegenerative movement disorder shown recently to be caused, in the majority of cases, by large biallelic ‘AAGGG’ STR expansions in the gene *RFC1 (9, 41)*. Expanded STR alleles in *RFC1* are relatively common and exist in several different motif conformations. The motifs ‘AAAAG_exp_’ and ‘AAAGG_exp_’ are considered non-pathogenic, regardless of size. In addition to the canonical pathogenic motif ‘AAGGG_exp_’, a rare ‘ACAGG_exp_’ motif and mixed ‘AAAGG_10-25_AAGGG_exp_’ conformation are both considered pathogenic, while the pathogenicity of various other observed conformations is currently unknown *(9, 42-44)*. The requirement to resolve very large STR expansions with diverse motifs, in an allele-specific fashion, makes genetic diagnosis of CANVAS patients challenging, often yielding inconclusive results.

Targeted sequencing with ReadUntil obtained a median of 15 spanning alignments at the STR site within the second intron of *RFC1*. Haplotype-resolved STR assembly revealed a variety of different pentanucleotide repeats ranging in size from 8 to 1070 copies across the cohort, with strong concordance to molecular testing by Southern blot and/or RP-PCR (R^2^ = 0.946; **Figure 3A,B**; **Figure 4A**). Large biallelic STR expansions of the pathogenic motif (AAGGG_410-1070_) were detected in 5/6 CANVAS patients but not unaffected individuals (*n* = 21; **Figure 3A**; **Figure 4A**).

**Figure 3.**
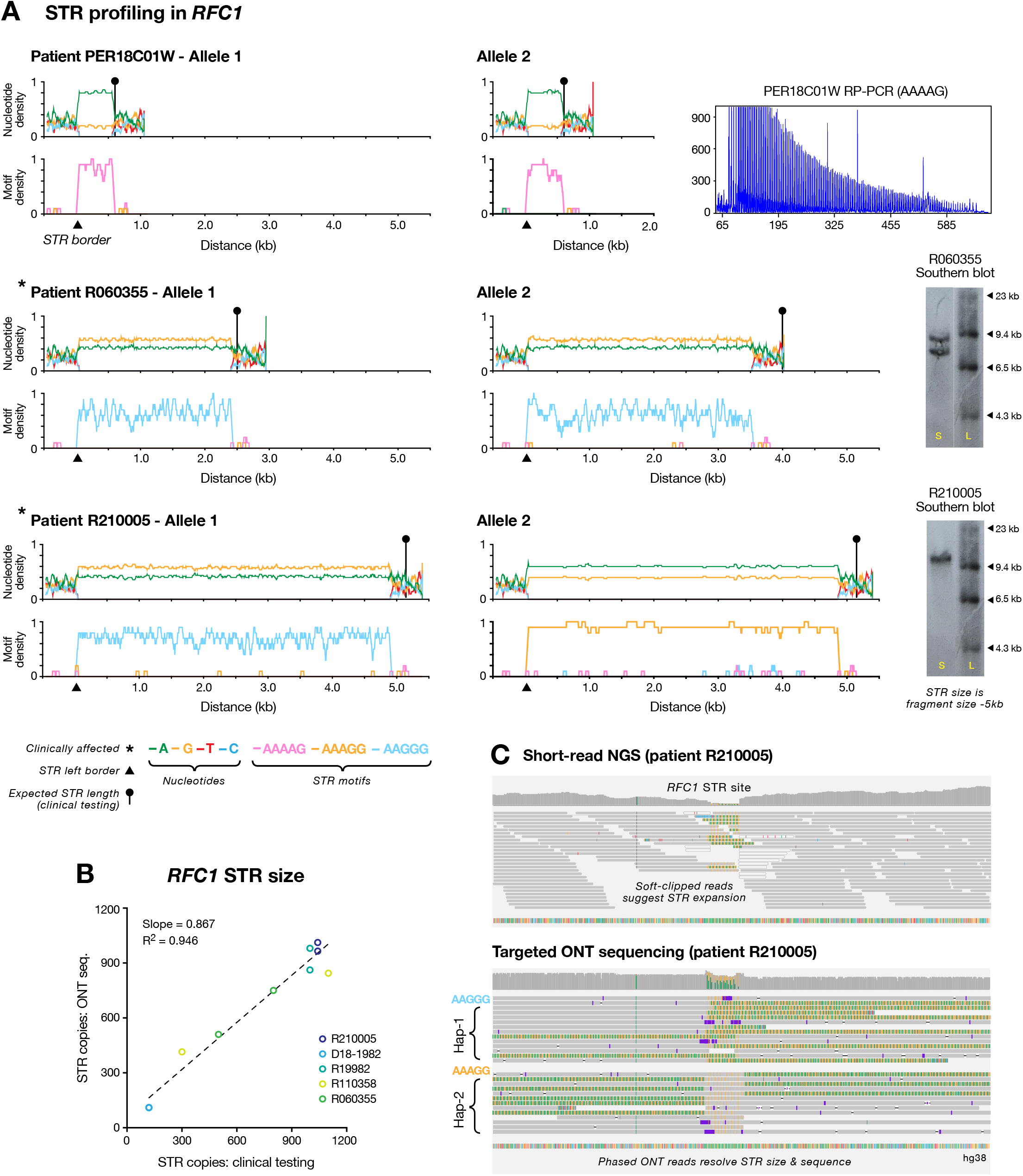
Haplotype-resolved assembly of pathogenic STR site *RFC1*. (**A**) Line plots show nucleotide content (upper) and density of pentanucleotide STR motifs (lower) enumerated in a 50bp sliding window across assembled STR alleles (including 1 kb up/downstream flanking sequences). Data is shown for three consenting individuals that were subject to clinical testing for STR expansions in *RFC1*. Relevant molecular testing data (RP-PCR or Southern blot) is shown for each individual (see **Supplementary Table 3**). Asterisks indicate CANVAS-affected patients, triangles show position of left border of assembled STRs and circular markers show the expected length of STR alleles, as determined by clinical testing. (**B**) Scatter plot shows lengths of pentanucleotide STR alleles in RFC1 in consenting individuals (n = 5), as determined by ONT sequencing vs molecular testing. (**C**) For patient R210005, genome browser views show short-read NGS alignments (upper) at the pathogenic STR site in *RFC1*. The presence of soft-clipped bases suggests an STR expansion is present, but the size, sequence and allelicity cannot be directly determined. The lower panel shows phased ONT alignments from the same sample. Long reads directly measure the STR expansion size and reveal distinct motif conformations on the two *RFC1* alleles.

**Figure 4.**
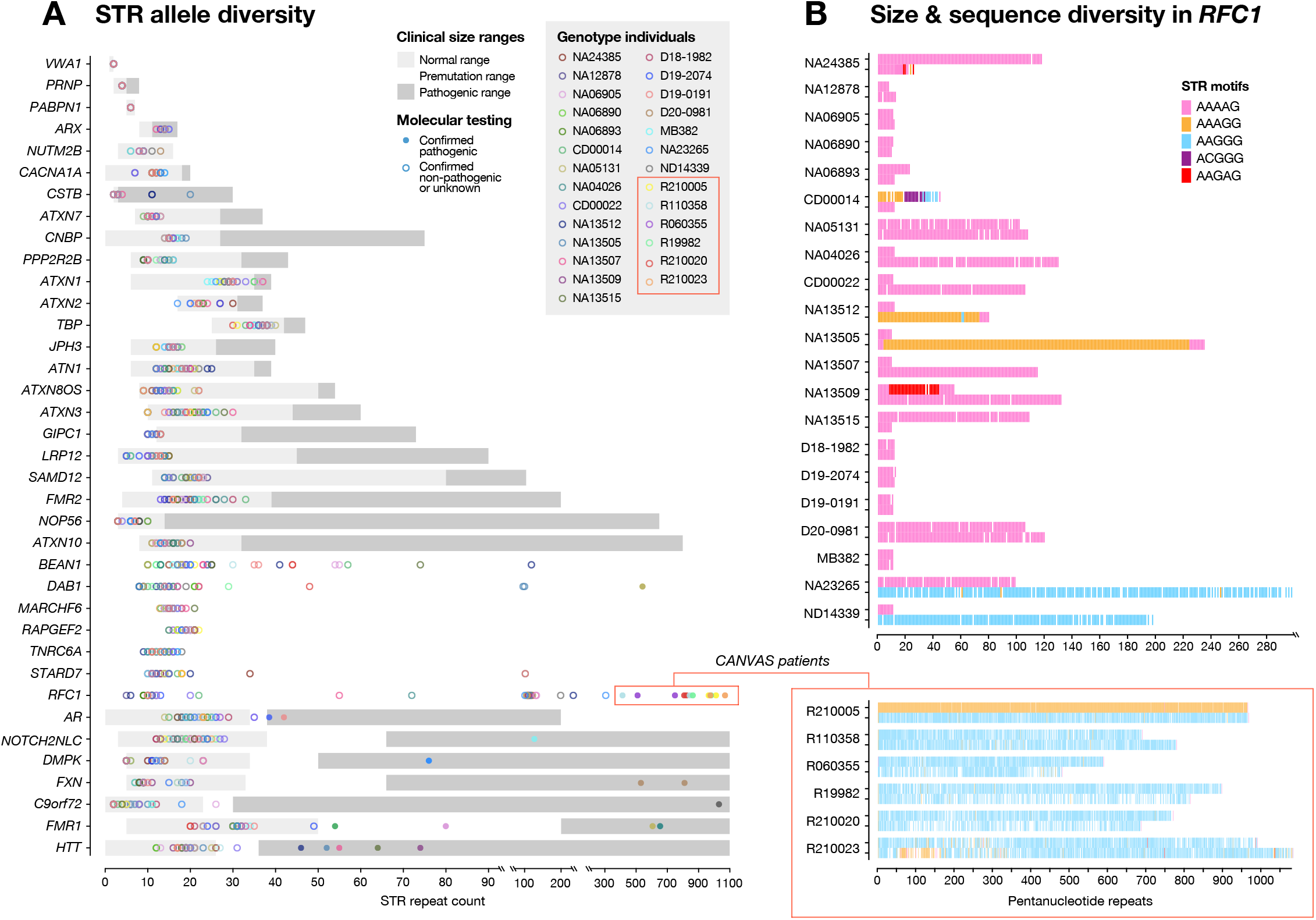
Diversity of STR alleles across the study. (**A**) Dot plot shows observed sizes of STR alleles for each gene (n = 37) in all individuals assessed during our study (n = 28). Grey boxes mark expected size ranges for normal, premutation and pathogenic STR alleles for each gene, where known. Filled circles indicate pathogenic alleles confirmed by clinical molecular testing, and empty circles were confirmed as non-pathogenic or were not tested. Full results for each individual gene are provided in **Fig. S2**. (**B**) Motif-barcharts show observed sizes and motif conformations of STR alleles assembled for the *RFC1* gene in each individual (n = 28). Red frame identifies CANVAS-affected patients, where large STR expansions in *RFC1* were detected by clinical testing and ONT sequencing.

In the single remaining CANVAS patient (R210005, previously reported as R19955 *(45)*), we detected a large (∼5kb) biallelic expansion at the *RFC1* STR site. However, the pathogenic ‘AAGGG’ motif was found on only one allele, with the other harbouring the ‘AAAGG’ motif that is currently considered non-pathogenic *(9)* (**Figure 3A**). Because the two alleles are equivalent in size and the pathogenic motif is present, standard molecular testing suggested that this individual possessed a biallelic pathogenic expansion (**Figure 3A**). Since both STR alleles are much greater than the read length of short-read NGS platforms, analysis by clinical whole-genome sequencing was also inconclusive (**Figure 3C**). In contrast, our assay readily distinguished the two STR alleles, identifying distinct ‘AAGGG_1010_’ and ‘AAAGG_960_’ conformations (**Figure 3A,C**). This highlights the utility of long-read sequencing for profiling large, complex STRs. In addition, this finding suggests potential pathogenicity of the *RFC1* ‘AAAGG’ expansion, as opposed to the current assumption that this motif is non-pathogenic, regardless of size *(9)*.

##### SBMA, DM1, NIID, FRDA & ALS

To further demonstrate the broad utility of our assay we analysed patients affected with SBMA (*n* = 2), DM1 (*n* = 1), NIID (*n* = 1), FRDA (*n* = 1) and ALS (*n* = 1). In all cases, we were able to correctly genotype the relevant STR (**Fig. S2Ci-Gi**). Detailed description of the results pertaining to each disorder are provided in a **Supplementary Note**.

In summary, the results described above demonstrate accurate, haplotype-resolved sizing, sequence determination and DNA methylation profiling of neuropathogenic STR loci using targeted ONT sequencing. This establishes analytical validity for genetic diagnosis of STR expansion disorders and highlights numerous advantages to this approach.

### Resolving STR diversity

STR sequences are highly polymorphic *(1, 2)*, yet their true diversity is likely under-appreciated due to limitations in current genotyping methods *(3, 4)*. Clinical interpretation relies on our ability to distinguish pathogenic alleles from the diversity of STRs encountered in healthy individuals. By determining the size and sequence of every allele of every disease-associated STR site in every individual tested, our targeted sequencing assay provides valuable data to help define the genetic landscape of STRs in human populations.

We observed a diverse array of STR alleles across our cohort (*n* = 27), which are visualised in full for each gene in **Fig. S2Ai-Jii** and summarised in **Figure 4A**. The diversity of STR sizes among clinically non-affected individuals was most evident in the pentanucleotide-repeat genes *RFC1* (8-298 copies), *DAB1* (8-541 copies), *BEAN1* (10-119 copies) and *STARD7* (10-102 copies; **Figure 4A,B**). These genes also harboured multiple unique STR motif conformations. For example, among 21 individuals not affected by CANVAS, we identified two carriers of pathogenic ‘AAGGG’ STR alleles in *RFC1*, and two carriers of noncanonical ‘ACGGG’ and ‘AAGAG’ motifs that are of unknown pathogenicity, while the remaining individuals had non-pathogenic ‘AAAGG’ and ‘AAAAG’ alleles (**Figure 4B**).

Other recently discovered pentanucleotide-repeat genes showed similar polymorphism. An ‘ATTTT’ STR site within *STARD7* was recently linked to familial adult myoclonus epilepsy 2 (FAME2), with affected individuals harbouring inserted ‘ATTTC’ or ‘AAATG’ motifs *(46)*. While we did not genotype any FAME2 patients, we observed novel ‘AAACT’ (*n* = 3) and ‘AACAT’ (*n* = 4) *STARD7* alleles in our cohort (**Fig. S2Hi**). Spinocerebellar ataxia 31 (SCA31) is caused by insertion of ‘TGGAA’ motifs immediately upstream from a ‘TAAAA’ STR in *BEAN1 (47)*. It has been reported that > 99% of healthy individuals possess an ‘TAAAA_8-20_’ allele at this site, although other motifs have been observed. Surprisingly, of 27 individuals in our cohort, only 16 harboured two ‘normal’ (TAAAA_8-20_) alleles. Of the remaining individuals, seven harboured expanded ‘TAAAA’ alleles (> 20 copies) and two had expanded alleles of predominantly ‘AAACT’ and ‘AACAT’ motifs (**Fig. S2Ii**). The allelic diversity of these genes was in contrast to other pentanucleotide-repeat genes *ATXN10, SAMD12, MARCHF6, RAPGEF2* and *TNRC6A*, where STRs alleles were largely uniform in size and sequence across the cohort (**Figure 4A**; **Fig. S2**).

Our analysis also revealed consistent detection of internal STR motif interruptions (**Fig. S2**; **Supplementary Table 6**). In addition to ‘AAG’ interruptions in *FMR1* and ‘CAA’ interruptions in *HTT* (discussed above), we detected known ‘CAT’ interruptions in *ATXN1*, ‘CAA’ interruptions in *ATXN2* and ‘CAA’ interruptions in *ATXN3* (**Fig. S2Bii-Dii**). We also detected motif interruptions in several other genes, including *ATXN10* (‘ATTGT’ interruptions; *n* = 6), *TBP* (‘CAA’ interruptions; *n* = 27), *CNBP* (‘GCTG/TCTG/GGCT’ interruptions; *n* = 27), *NOTCH2NLC* (‘GGA’ interruptions; *n* = 26) and *AR* (‘CAA’; *n* = 1; **Fig. S2**).

While our study does not encompass a sufficiently large and unbiased cohort to draw general conclusions, it is clear that targeted long-read sequencing will help to describe a currently under-appreciated diversity of STR alleles, and likely redefine the features of non-pathogenic alleles for several genes. Moreover, it will facilitate more detailed investigation of genotype-phenotype correlations as well as potential disease modifiers of pathogenic repeat expansion alleles, such as interruptions.

### Informative secondary targets: pharmacogenomics genes

In addition to disease-associated STR genes, our ReadUntil sequencing panel included a range of other targets that may provide further clinical insights (see **Supplementary Table 2**). Since target selection is programmable, such secondary targets come at no additional cost and can be flexibly included/excluded on an individual basis. As an example, we analysed 28 pharmacogenomics (PGx) genes included on the panel (**Supplementary Table 2**). PGx genotyping can anticipate an individual’s capacity to metabolise specific drugs and prevent adverse drug reactions *(48)*.

Many PGx genes, including the prototypical example *CYP2D6* (**Figure 5A**), are highly polymorphic, frequently harbour structural variation and possess close homologs, making them difficult to genotype using standard molecular techniques or short-read NGS *(48, 49)*. Despite their complex architectures, ReadUntil targeted enrichment was effective within PGx genes, achieving a median 25X coverage depth (**Figure 5B**). Notably, the breadth and evenness of coverage by unique ONT sequencing alignments in PGx genes was superior to whole-genome short-read NGS on matched samples (**Figure 5B,C**). This is best illustrated at the *CYP2D6* locus, where ONT ReadUntil achieved complete coverage and phasing of *CYP2D6* and its neighbouring pseudogene *CYP2D7*, unlike short-read NGS (**Figure 5A**). This emphasises the advantages of long-read sequencing at complex genomic loci.

**Figure 5.**
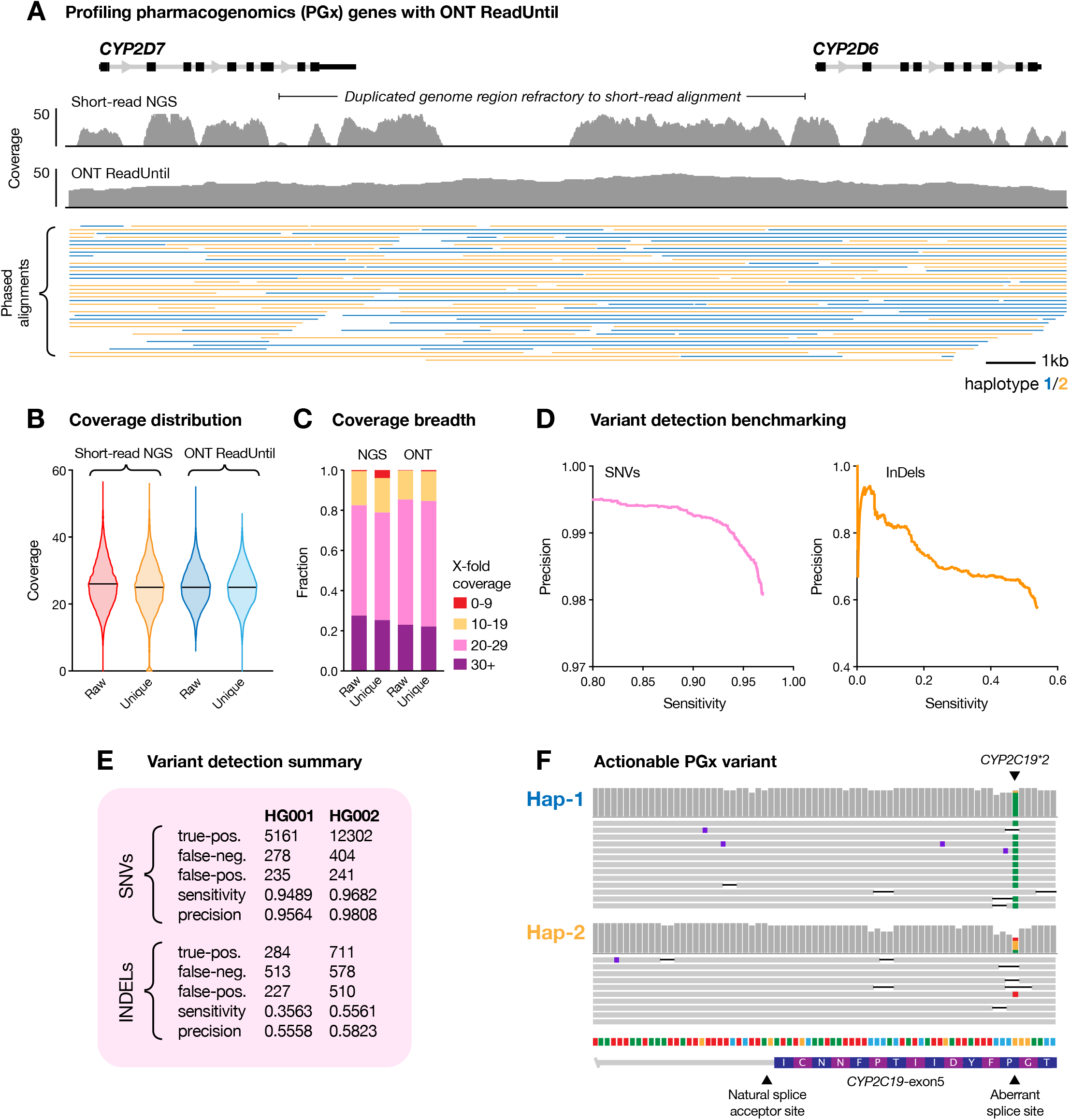
Targeted ONT sequencing of pharmacogenomics (PGx) genes. (**A**) Genome browser view shows coverage distribution for uniquely aligned reads (MapQ>=30) at the *CYP2D6* gene and neighbouring pseudogene *CYP2D7*. The upper track shows data for short-read whole-genome sequencing (Illumina NovaSeq) of the human reference sample HG001/NA12878 (see **Supplementary Table 3**). The lower track shows coverage and phased alignments for ONT ReadUntil targeted sequencing on the same sample. (**B**) Violin plots show coverage distribution across PGx gene targets (n = 28) with short-read NGS (left) and targeted ONT sequencing (right). For each technology, both raw alignment coverage and unique alignments (MapQ>=30) are shown. (**C**) For the same datasets, stacked barcharts show the fraction of PGx target regions covered at different sequencing depths (red 0-9X, yellow 10-19X, pink 20-29X & purple >=30X). (**D**) Precision-recall curves show accuracy of variant detection within PGx gene targets for SNVs (left; pink) and InDels (right; orange) using ONT ReadUntil on reference sample HG002. Precision recall curves were used to determine optimum Nanopolish parameter settings. (**E**) Summary table showing final variant detection statistics within PGx targets. While InDel accuracy is poor, SNVs were detected with relatively high sensitivity and precision. (**F**) Genome browser view showing an example of a clinically-actionable PGx allele (*CYP2C19*2*) detected using ONT ReadUntil in HG001.

We analysed the reference human DNA samples HG001 & HG002 (NA12878 & NA24385) and compared the results to high confidence annotations from the Genome in a Bottle (GIAB) project to evaluate variant detection within PGx loci (see **Methods**). After parameter optimisation with the software *Nanopolish (50)*, we observed accurate detection for SNVs on both samples (95%, 97% sensitivity and 96%, 98% precision; **Figure 5D,E**). However, the high frequency of insertion-deletion errors in ONT reads meant that indel variants could not be reliably detected (sensitivity 36%, 56% and precision 56%, 58%; **Figure 5D,E**).

In HG001 and HG002, respectively, we detected 40 and 43 annotated PharmVar SNVs in key PGx genes *CYP2B6, CYP2C9, CYP2C19, CYP2D6, DPYD*, several of which show evidence of pharmacogenomic phenotypes *(51)*. In HG001, for example, we identified a single SNV (c.681G>A) in *CYP2C19* that is known to introduce an aberrant splice-acceptor site and truncation of the canonical ORF (**Figure 5F**) *(52)*. This allele (*CYP2C19*2*) affects the metabolism of multiple medications and is associated with toxicity for the common antidepressants citalopram and escitalopram (Level 1A evidence) *(53)*. Given that prescription of antidepressants to patients with repeat expansion disorders such as HD is relatively common *(54)*, this example demonstrates the potential utility of parallel PGx genotyping in STR patients. While this is not the primary purpose of our targeted sequencing assay, secondary findings of this nature may better inform patient care at no extra cost, underscoring the appealing flexibility of programmable sequencing with ONT ReadUntil.

## DISCUSSION

This study demonstrates the validity and utility of programmable targeted nanopore sequencing for genetic diagnosis of STR expansion disorders. Unlike existing single-gene molecular techniques, our approach enabled unbiased sizing and sequence determination of all known neuropathogenic STR sites in a single targeted assay. Haplotype-resolved assembly of ONT reads was used to solve large and complex STR expansions, such as those described in *RFC1*, that eluded characterisation by standard molecular testing and short-read NGS. Moreover, we identified motif interruptions within STR sequences and local DNA methylation profiles that further inform pathogenicity. Given these capabilities, we propose that targeted sequencing with ONT ReadUntil *(25, 26)* can address the pressing need for improved methods for molecular characterisation of STR expansions.

Our multi-gene assay has the additional benefit of informing consenting patients of carrier status for pathogenic expansions not associated with their primary diagnosis (e.g. *RFC1* ‘AAGGG’ pathogenic expansion in the DM1 patient NA23265). The use of long read sequencing also enables the detection and phasing of heterozygous SNVs nearby pathogenic STR sites. Phased SNVs are useful genetic markers that distinguish pathogenic and non-pathogenic haplotypes during family genetic studies or preimplantation genetic diagnosis (PGD) *(55)*, further supporting the utility of our assay.

ONT ReadUntil permits the flexible inclusion of virtually any additional secondary targets for genetic analysis. In this study, targeted sequencing of 28 pharmacogenomics (PGx) genes identified clinically actionable PGx alleles that may be used to guide personalised selection of medications. The value of such secondary targets, which come at no extra cost, will continue to increase with ongoing improvement in nanopore sequencing accuracy *(56)*, particularly if this enables reliable detection of indel variants that were beyond current capabilities.

While the potential benefits for genetic diagnosis of patients with STR expansion disorders are clear, targeted STR sequencing will be similarly useful as a research tool. STRs are highly polymorphic and exhibit pathogenicity through an array of different mechanisms. Much remains to be learnt about the basic biology of STR expansion disorders and the distinction between a benign/pathogenic allele, particularly for newly discovered STR disease genes like *RFC1 (9), GIPC1 (10), LRP12 (11), NOTCH2NLC (11, 12), VWA1 (13)*, etc. By resolving the full diversity of STR sizes and motif conformations in clinically affected and non-affected individuals our targeted ONT sequencing assay may be applied to better define pathogenic boundaries and investigate the role of phenotypic modifiers, such as internal STR interruptions. For example, in our relatively small cohort, we observed several novel STR alleles in genes such as *STARD7* and *BEAN1*, the physiological relevance of which are currently unknown. We anticipate that elucidation of the full complement of STR expansion size, motif and interruptions in health and disease will reveal new genotype-phenotype correlations, enabling better understanding of the pathomechanisms and facilitating rationale treatment design.

Haplotype-resolved DNA methylation profiling of STR expansion genes by ONT sequencing will also shed new light on their epigenetic regulation and potential roles in pathogenic mechanisms. DNA methylation has been extensively studied in males with Fragile X Syndrome, wherein expanded ‘CGG’ STR alleles trigger hypermethylation of the *FMR1* promoter, resulting in gene silencing *(36, 37)*. Consistent with this, we observed promoter hypermethylation in FXS-affected males. Furthermore, we identified haplotype-specific hypermethylation of the FMR1 promoter in one female who was a FXS premutation carrier. While previously observed, the significance of DNA methylation in premutation carriers is not fully understood *(39, 40)* and can only be detected using haplotype-aware methodologies, such as ONT sequencing. DNA hypermethylation has also been described in some carriers of pathogenic STR expansions in *C9orf72*, however the association between methylation status, repeat size and clinical phenotype is not clear *(57)*, warranting further interrogation with improved molecular tools. Little is known about the epigenetic regulation of most other genes on our targeted sequencing panel, highlighting the need for further investigation, with ONT sequencing being a powerful tool for this purpose.

Finally, by resolving STR expansions that are not amenable to existing techniques, long-read sequencing promises to accelerate the discovery of new repeat expansion genes and disorders *(3, 4)*. While targeted sequencing is not suitable for discovery of genes with no prior evidence, the flexible nature of ReadUntil sequencing is ideal for profiling tens/hundreds of candidate genes/regions, such as those identified by linkage mapping in affected families *(12, 23, 58)*. Moreover, repeat expansions need not be the only pathogenic variants discovered by this approach, with targeted long read sequencing also suitable for the detection of other types of structural variation *(59)*. We anticipate that this will be a powerful approach to STR gene discovery and provide molecular diagnoses for many previously unsolved cases in the future.

## MATERIALS & METHODS

### Sample collection, processing & molecular testing

Patient-derived genomic DNA reference samples (NA* and CD* prefixes) and accompanying clinical notes were obtained from the Coriell Institute (https://www.coriell.org/). Molecular testing results were available for STR expansions in *HTT, FMR1, DMPK* or *C9orf72*, as relevant for each individual’s phenotype and/or family history. Upon receiving, genomic DNA was resuspended in nuclease free water at ∼100 ng/µL and stored at −20C.

Patients consulting at neurology clinics in New South Wales (NSW) were consented for genomic analysis by nanopore sequencing under St Vincent’s Hospital Human Research Ethics Committee protocol 2019/ETH12538. Patients in Western Australia (WA) were consented under Human Research Ethics Committee of the University of Western Australia protocol 2019/RA/4/20/1008. Deidentified patient samples were subject to diagnostic testing in certified clinical laboratories, according to current clinical practice, and molecular test data was provided for this study. STR sites in *RFC1* (for CANVAS patients), *NOTCH2NLC* (for NIID), *FXN* (for FRDA) and *AR* (for SBMA) were tested by repeat-primed PCR and large expansions in *RFC1* and *FXN* were further analysed by Southern blot to determine STR sizes. High-molecular weight (HMW) genomic DNA was extracted from patient blood samples using Qiagen Gentra PureGene Blood Kit (NSW) or QIAsymphony DSP DNA Midi Kit (WA) and suspended in nuclease free water. De-identified samples were transferred to the Kinghorn Centre for Clinical Genomics for nanopore sequencing analysis.

Full sample descriptions are provided in **Supplementary Table 3**.

### ONT library preparation and sequencing

Prior to ONT library preparations, the DNA was sheared to ∼15kb fragment size using Covaris G-tubes and visualised, post-shearing, on an Agilent TapeStation. Nanopore sequencing libraries were prepared from ∼1.5-5ug of HMW DNA, using native library prep kits (SQK-LSK109 or SQK-LSK110), according to the manufacturer’s instructions. Each sample was loaded onto an ONT MinION flow cell (R9.4.1) and sequenced on either an ONT GridION or ONT MinION device with live target selection/rejection executed by the *ReadFish* software package *(26)* (see below). Samples were run for a maximum duration of 72 hours, with nuclease flushes and library reloading performed at approximately 24 and 48 hour timepoints to maximise sequencing yield.

### Programmable target selection

Targeted sequencing was performed using the open-source software package *Readfish (26)* that internally uses the ONT ReadUntil API for live rejection of off-target sequencing fragments. The targets used in this study were whole gene loci +/-50kb of flanking genome sequence to ensure coverage of promoter and UTR regions, and other local regulatory elements. Our targeted sequencing panel included genes containing pathogenic STR sites associated with neurological disease (*n* = 37; see **Supplementary Table 1**), as well as other potentially informative targets, such as Pharmacogenomics genes exhibiting genotype-drug relationships designated as clinically actionable by the Clinical Pharmacogenetics Implementation Consortium (*n* = 28) and genes harbouring clinically actionable mendelian mutations, as designated by the American College of Medical Genetics (n = 59). A full description of ReadUntil targets is provided in **Supplementary Table 2**.

We Installed *Readfish* version 0.0.5a1 inside a Python virtual environment (with system default Python version given in **Supplementary Table 5**) using *pypi*. When the study was commenced, this *Readfish* version (0.0.5a1) was the version that supported the available MinKNOW-core and *Guppy* versions (major version 4). However, the dependencies (*ont-pyguppy-client-lib* and *pyguppyclient*) installed with *Readfish* v0.0.5a1 by default were incompatible with the *Guppy* versions on our machines. That is, the support for the NVIDIA 3090 GPU card on our workstation was only introduced in *Guppy* v4.2.2 and we did not want to interfere with the *Guppy* v4.2.3 already installed on the GridION. Therefore, we manually installed *ont-pyguppy-client-lib* and *pyguppyclient* (versions in **Supplementary Table 5**) with minor source code changes to accommodate the *Guppy* versions.

We created a new minKNOW configuration profile under */opt/ont/minknow/conf/package/sequencing/sequencing_MIN106_DNA_readfish_real*.*toml* with equivalent configuration values except that the value for the *break_reads_after_seconds* attribute was changed from 1.0 to 0.4, as instructed in the *Readfish* documentation. The reference index for *Readfish* was created using *minimap2 (60)* with *-x map-ont* profile using the hg38 genome with alternate contigs excluded. Experiments run for this study use the high *Guppy* accuracy base-call configuration. *Readfish* parameters were configured such that: (i) if the query sequence aligns to a one or more locations within the target genome regions, sequencing proceeds with no further checking (*single_on = “stop_receiving”, multi_on = “stop_receiving”*); (ii) if the query sequence aligns to one or more regions not in the desired list, the read is rejected by ReadUntil (*single_off = “unblock”, multi_off = “unblock”*); (iii) if the base-called sequence is unavailable or if the query sequence is unmappable, sequence continues and the read is re-checked in the subsequent round (*no_seq = “proceed”, no_map = “proceed”*).

### Haplotype-resolved assembly & methylation profiling of STR sites

Raw ONT sequencing data was base-called using *Guppy* (4.4.1) and reads with mean quality < 7 were excluded from further analysis. Resulting FASTQ files were aligned to the hg38 reference genome using *minimap2* (v2.14-r883) *(60)*.

To analyse STR sites, we retrieved all alignments within a 50kb window centred on a given STR site. These were converted back to FASTQ format, assembled *de novo* using *Flye* (v2.8.1-b1676) *(61)* and polished using *Racon* (v1.4.0) *(62)* to generate a pseudo-haploid contig encompassing the STR region. Starting reads were realigned to this contig and then assigned to separate alleles/haplotypes via each of three methods: (i) phasing by consideration of heterozygous SNVs within the STR region using *Longshot* (v0.4.1) *(63)*; (ii) phasing by consideration of heterozygous SVs at the STR site w.r.t. the assembled contig using *Sniffles* (v1.0.9) *(64)*; (iii) a custom all-vs-all pairwise sequence alignment and similarity clustering method that identifies allelic differences in STR size and sequence, based on read clustering, with no prior information (see **Data & Code Availability**). After phasing, the initial assembled contig was re-polished separately with reads from either allele/haplotype using *Racon* (v1.4.0) *(62)*, yielding two haploid contigs encompassing the STR site. The position of the relevant STR site was identified in each contig by mapping 150 bp unique flanking sequences (extracted from hg38) using *minimap2* (2.14-r883) *(60)*. STR size, motif and summary statistics were retrieved using *Tandem Repeat Finder* (4.09) *(65)*, followed by manual inspection and motif counting. This process was performed separately for each STR site in each individual sample.

For DNA methylation analysis, FAST5 files containing ONT signal data were converted to compressed binary SLOW5 format *(66)* using *slow5tools* (https://github.com/hasindu2008/slow5tools). DNA methylation calling was run on all on-target alignments (w.r.t. hg38) using *F5C call-methylation* (v0.6) *(67)*, which is a GPU accelerated version of the popular *Nanopolish* software package *(50)*. Meth-calling results were retrieved for all phased reads at each STR site (see above) based on read IDs and methylation frequencies were computed separately for each allele/haplotype, as well as for total unphased reads, using the *F5C meth-freq*. When enumerating the local methylation status of a given STR site, we considered all CpGs within 1500 bp with at least 5 meth-called reads on each allele/haplotype. Methylation frequency profiles were converted to bigwig format for visualisation in IGV, using *kentutils* (https://github.com/ENCODE-DCC/kentUtils).

### Genotyping pharmacogenomics (PGx) genes

Raw ONT sequencing data was base-called using *Guppy* (v4.4.1) and reads with mean quality < 7 were excluded from further analysis. Resulting FASTQ files were aligned to the hg38 reference genome using *minimap2* (v2.14-r883) *(60)* and filtered for unique alignments (MapQ>=30). Variant calling within PGx regions (*n* = 28) was performed using *Nanopolish* (v0.13.2) *(50)*, with candidate variants requiring a minimum read depth of 5 (-d 5). We compared the resulting variant candidates detected in HG001 (NA12878) and HG002 (NA24385) to high-confidence variant annotations available for these samples from the Genome in a Bottle (GIAB) project. The comparisons were made with the RTG tools *vcfeval* utility (v3.11) with the --squash-ploidy parameter selected. Precision recall curves created by RTG tools were used to determine an optimum variant filtering parameters of QUAL >= 30 & BCRV >= 5. Performance summary statistics were calculated after applying these filters, using the following definitions:

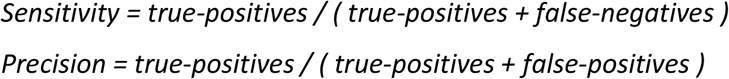

To identify actionable PGx variants, we queried filtered variant candidates against PharmVar variant annotations *(51)*.

## Data Availability

All software used in this study is free and open source, with the exception of the proprietary ONT base-calling software Guppy. Where permitted under patient ethics protocols, sequencing data from this study has been uploaded to the NCBI Sequence Read Archive (SRA) and will be open for public access at the time of publication.

## DATA & CODE AVAILABILITY

All software used in this study is free and open source, with the exception of the proprietary ONT base-calling software *Guppy*. Custom code used for all-vs-all phasing of STR alignments will be made available in a public Github repository at the time of publication. Where permitted under patient ethics protocols, sequencing data from this study has been uploaded to the NCBI Sequence Read Archive (SRA) and will be open for public access at the time of publication.

## ACKNOWLEDGEMENTS

We thank Michael Halmagyi for support in patient recruitment. We thank Mike Vella and NVIDIA for their kind donation of a world-class GPU that was used for ReadUntil experiments. We thank Derrick Lin and Garvan’s DICE team for providing excellent HPC support. We acknowledge the following funding support: Australian Medical Research Futures Fund (MRFF) Investigator Grant MRF1173594 (to I.W.D.), Australian National Health and Medical Research Council (NHMRC) Fellowship APP1122952 (to G.R.), MRFF Genomics Future Health Missions Grant 2007681 (to N.G.L.), Australian Government Research Training Program (RTP) Scholarship (to C.S.), philanthropic support from The Kinghorn Foundation (to I.W.D.), Margaret and Terry Orr Memorial Fund (to N.G.L.), Paul Ainsworth Family Foundation (to K.R.K.) and a Working Group Co-Lead Award from the Michael J. Fox Foundation, Aligning Science Across Parkinson’s (ASAP) initiative (to K.R.K.).

## AUTHOR CONTRIBUTIONS

S.R.C., S.S.P., K.R.K. & I.W.D. conceived the project, designed the targeted sequencing panel and planned experiments. K.N., M.H., M.T., V.F., A.C., C.K.S., M.R.D., N.G.L., G.R., M.K., S.S.P. & K.R.K. were involved in patient recruitment and clinical interpretation. C.K.S., C.DS., H. H. & A.C. performed diagnostic molecular assays. I.S. processed samples and prepared ONT libraries. H.G. & J.M.F. designed and built custom computer hardware for use in ONT ReadUntil experiments. I.S. & H.G. performed ReadUntil experiments. I.S., S.R.C, H.G., J.M.F. & I.W.D. performed bioinformatics analysis. I.S., S.R.C. & I.W.D. prepared the figures. I.S., S.R.C., K.R.K. & I.W.D. prepared the manuscript with support from all authors.

## DISCLAIMERS & COMPETING INTERESTS

I.W.D. manages a fee-for-service sequencing facility at the Garvan Institute of Medical Research that is a customer of Oxford Nanopore Technologies but has no further financial relationship. H.G. & J.M.F. have received travel and accommodation expenses to speak at Oxford Nanopore Technologies conferences. The authors declare no other competing financial or non-financial interests.

## Notes

### Author Declarations

Patients consulting at neurology clinics in New South Wales (NSW) were consented for genomic analysis by nanopore sequencing under St Vincent's Hospital Human Research Ethics Committee protocol 2019/ETH12538. Patients in Western Australia (WA) were consented under Human Research Ethics Committee of the University of Western Australia protocol 2019/RA/4/20/1008. Approval was granted in both cases.

