## Supplementary_Materials for "Comprehensive genetic diagnosis of tandem repeat expansion disorders with programmable targeted nanopore sequencing"

### LIST OF SUPPLEMENTARY MATERIALS

**Supplementary Table 1**. Gene-centric list of neuropathogenic short-tandem repeat expansion disorders.

**Supplementary Table 2.** Genome coordinates of ReadUntil targeted sequencing panel w.r.t. the hg38 reference genome.

**Supplementary Table 3.** List of individuals analysed during the study and relevant clinical information.

**Supplementary Table 4.** List of ReadUntil targeted nanopore sequencing runs and performance metrics.

**Supplementary Table 5.** Technical specifications of computers used to run ReadUntil targeted sequencing experiments.

**Supplementary Table 6.** Gene-centric summary statistics for short-tandem repeat (STR) assembly.

**Supplementary Note.** Description of analysis pertaining to spinal and bulbar muscular atrophy of Kennedy (SBMA), myotonic dystrophy 1 (DM1), neuronal intranuclear inclusion disease (NIID), Friedreich ataxia (FRDA) and amyotrophic lateral sclerosis (ALS).

**Figure S1.** Performance benchmarking of ReadUntil targeted STR sequencing.

**Figure S2.** Haplotype-resolved assembly of disease-associated STR sites.

### SUPPLEMENTARY NOTE

##### Spinal and bulbar muscular atrophy (SBMA)

SBMA (commonly known as Kennedy's Disease) is a chrX-linked progressive lower motor neuron neurodegenerative disease. SBMA is caused by 'CAG' STR expansions of > 34 copies within the *AR* gene *(68)*. Fully penetrant pathogenic alleles harbour ≥ 38 repeats, while 36-37 repeats show incomplete penetrance and the clinical significance of 35 repeats is unclear *(69, 70)*. There is an inverse correlation between STR size and rate of disease progression *(68)*. We sequenced two individuals that were positively genotyped for STR expansions in *AR*: one SBMA-affected male (PER19S02A) and one carrier female (PER19S01C). Consistent with results from RP-PCR testing, ONT sequencing identified a 'CAG' expansion of 53 repeats in the affected male and a heterozygous 44/26 'CAG' expansion in the carrier female (**Fig. S2Ci**). No pathogenic STR expansions were identified in the remaining individuals (*n* = 25). ONT sequencing also identified one unaffected individual (MB382) harbouring a double 'CAA' interruption within the 'CAG' STR (**Fig. S2Ci**), which has not been described previously *(71)*.

##### Myotonic dystrophy type 1 (DM1)

DM1 is a progressive neuromuscular disease caused by 'CTG' STR expansions of ≥ 50 copies within the *DMPK* gene*.* The pathogenic repeat-size range is broad and accounts for varying disease severity; STR alleles of 50-150 repeats typically manifest in late-onset mild cataracts and myotonia while much larger expansions cause infantile-onset hypotonia, respiratory involvement and intellectual disability *(72)*. Our cohort included one DM1 patient (NA23265), in which we identified a heterozygous 'CTG' STR expansion of 76 copies within *DMPK*, consistent with clinical testing by RP-PCR (**Fig. S2Di**). All other individuals (*n* = 26) harboured CTG repeats of < 35 copies (**Fig. S2Di**). Previous studies suggest 3-5% of DM1 patients carry motif interruptions within the DMPK (CCG, GGC, CTC, CAG) that are thought to be protective and delay onset of symptoms by lowering progression of somatic instability over time *(73)*. We detected no such interruptions within our cohort of predominantly non-DM1 individuals, suggesting these are likely rare in healthy populations.

##### Neuronal intranuclear inclusion disease (NIID)

A heterozygous 'GGC' STR expansion of > 65 copies within the gene *NOTCH2NLC* has recently been implicated with several phenotypes including neuronal intranuclear inclusion disease (NIID), leukoencephalopathy, essential tremor, oculopharyngodistal myopathy type 3 (OPDM3), Parkinson's disease, multiple system atrophy (MSA), neuropathy and ALS *(11, 12, 74-80)*. Our cohort included one NIID patient (MB382), in which we identified a heterozygous 'GGC' STR expansion of 125 copies in *NOTCH2NLC*, consistent with clinical testing by RP-PCR (**Fig. S2Ei**). All other individuals (*n* = 26) harboured GGC repeats of < 65 copies. We consistently detected a 'GGA' interruption in all non-affected individuals (**Fig. S2Ei**). This supports previous studies suggesting the most common healthy allele conformation contains two 'GGA' interruptions *(81)*. 'GGA', 'AGC' and 'ACG' motif interruptions have also been reported, and may serve as a phenotypic modifiers but none of these were detected in our cohort *(76, 81)*.

##### Friedrich's ataxia (FRDA)

Friedrich's ataxia (FRDA) is the most common form of recessive hereditary ataxia, typically characterised by early onset, slowly progressive ataxia. FRDA is caused by a biallelic 'GAA' STR expansion within intron 1 of the *FXN* gene *(82)*. The pathogenic range is generally reported as 66-1300 'GAA' repeats, however this is not definitive: the exact differentiation between premutation alleles and pathogenic alleles remains poorly defined, as there are cases of disease-causing alleles within a premutation range of 44-66 'GAA' repeats *(83)*. STR length and the presence of interruptions are both considered phenotypic modifiers, with shorter, uninterrupted STR alleles associated with decreased disease severity. Our cohort included one FRDA patient (PER20F02A), in which we identified a biallelic 'GAA' STR expansion sized at 812 and 529 repeat copies, consistent with diagnostic testing by Southern blot (**Fig. S2Fi**). All other individuals (*n* = 26) harboured GAA repeats of < 21 copies, with no motif interruptions detected (**Fig. S2Fi**).

**C9orf72-related disorders**

A hexanucleotide 'GGGGCC' expansion within the gene *C9orf72* is the most common cause of familial amyotrophic lateral sclerosis (ALS) and/or frontotemporal dementia (FTD) *(84)*. The same STR expansion has also been linked to a range of other phenotypes including ataxia, myoclonus, chorea, parkinsonism, schizophrenia and bipolar disorder *(85–87)*. Healthy individuals usually carry alleles with < 30 repeat copies, while disease-causing alleles typically harbour several hundreds to thousands. However, a grey area exists for small expansions (up to 200 repeats), some carriers of which develop disease while others do not *(88)*. Our cohort included one ALS patient (ND14339), in which we identified a heterozygous hexanucleotide STR expansion in *C9orf72* of ~8kb in size, or ~1300 repeat copies, consistent with clinical testing (**Fig. S2Gi**). We obtained relatively few sequencing alignments spanning the expanded allele, which prevented accurate determination of the internal STR sequence. Deeper sequencing is required to confidently determine the motif conformation and potential presence of interruptions or other features.

Intermediate-sized STR expansion alleles (27-33 copies) within another STR gene on our panel, *ATXN2,* have also been associated with an increased risk of ALS, and it has been suggested that these may drive the clinical phenotype of C9-related disorders towards ALS rather than FTD *(88)*. We identified one such allele (PER18C01W; 27 repeats) within our sample population (*n* = 27; see **Fig. S2Cii**).

### EXTENDED REFERENCES

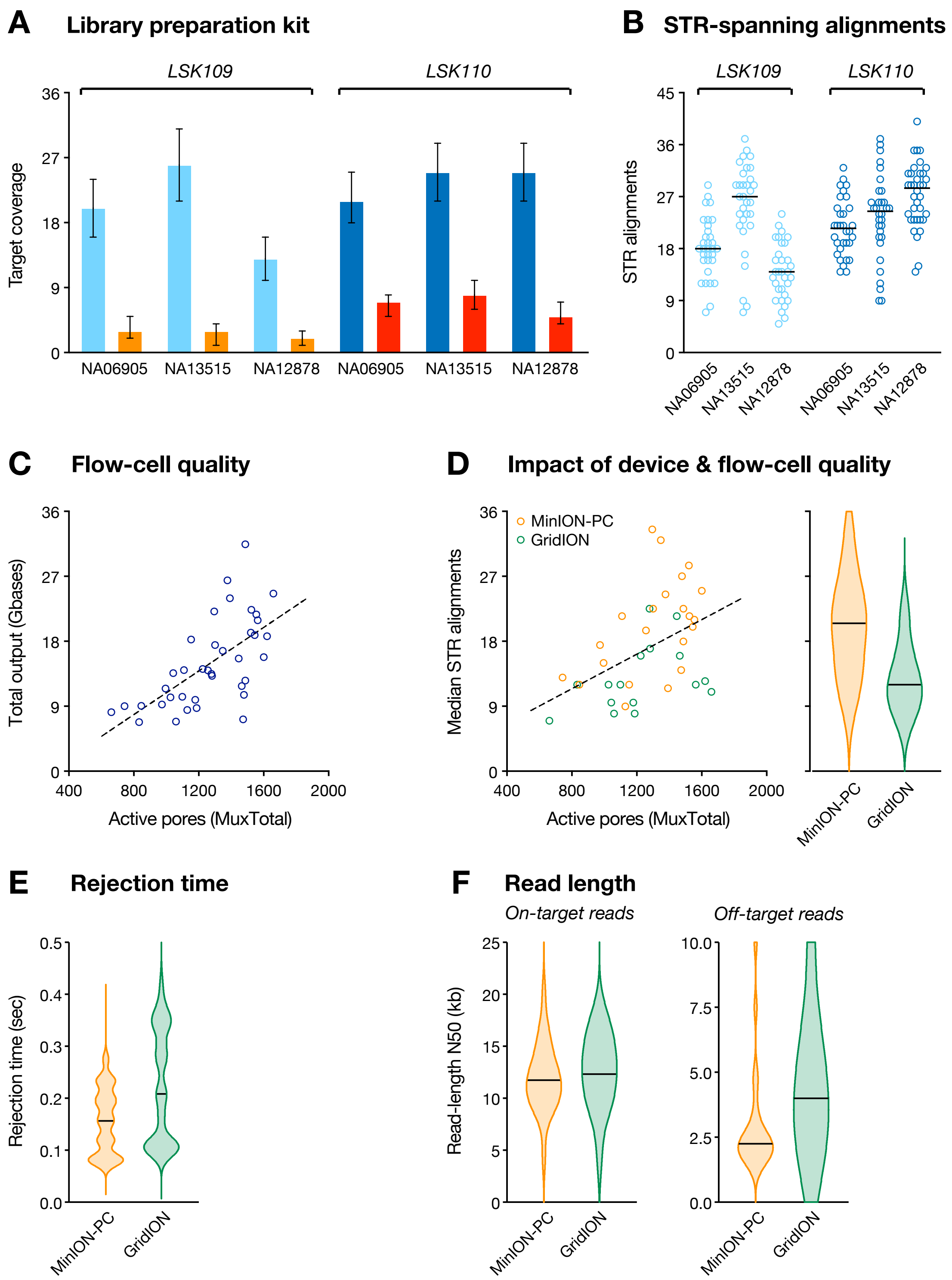


**Figure S1. Performance benchmarking of ReadUntil targeted STR sequencing.** (**A**) Bar chart shows per-base coverage within on-target regions (navy & blue) and randomly selected off-target genes (orange & red) during ReadUntil sequencing of three reference samples (NA06905, NA13515, NA12878; see **Supplementary Table 3**) run with both LSK109 (left) and LSK110 (right) library prep chemistry. Error bars show median +/- interquartile range. (**B**) For the same runs, dot plots show the number of alignments spanning STR sites (n = 37). Bars show median value within each experiment. (**C**) Scatter plot shows total sequencing output (Gbases) generated in ONT ReadUntil experiments (n = 38), relative to the starting number of active pores (MuxTotal) on a given ONT MinION flow-cell. (**D**) Scatter plot shows median number of alignments spanning target STR sites, relative to the starting number of active pores (MuxTotal) on a given ONT MinION flow-cell. Colours distinguish ReadUntil experiments run on an ONT GridION (green; NVIDIA Quadro GV100 GPU; n = 16) or a high-spec desktop PC (orange; NVIDIA 3090 GPU; n = 22); see **Supplementary Table 5** for full specs. Violin plots summarise the distribution of STR alignment counts in ONT ReadUntil experiments run GridION vs MinION-PC devices. (**E**) Violin plots summarise rejection times (i.e. time taken to reject a given off-target read) recorded during ONT ReadUntil experiments run on GridION vs MinION-PC devices. Rejection speed is influenced by the GPU on the device executing ONT base-calling, with the superior GPU on the desktop PC achieving superior speeds. (**F**) Violin plots summarise read-length N50s for on-target vs off-target alignments in ReadUntil experiments run on GridION vs MinION-PC devices. The superior read-rejection speeds on the MinION-PC device lead to shorter off-target reads and results in superior on-target coverage enrichment.

ai

*HTT*

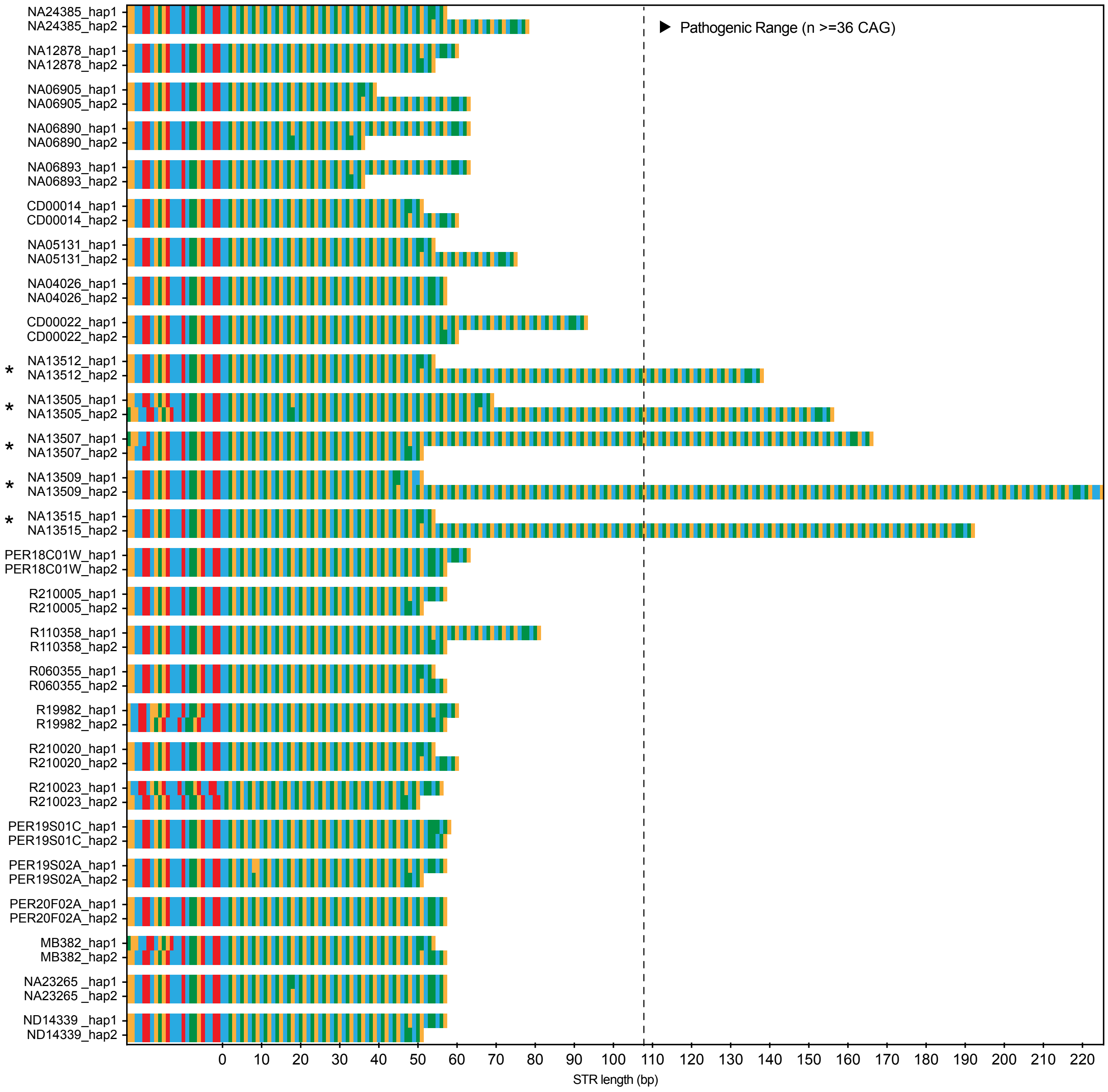


**READ DEPTH**
Range: 8-43x
Median: 18x

**REPEAT LENGTHS (#)**
Min: 12
Max: 74

**SEQUENCE**
Motifs Seen: CAG
Interruptions Seen: CAA

T C A G

* Clinically affected

**Figure S2. Haplotype-resolved assembly of disease-associated STR sites.** (**ai-jii**) Sequence-barcharts show STR alleles, including 25 bp of upstream flanking sequence, assembled from ONT sequencing of all individuals in the study (n = 27; see **Supplementary Table 3**). Each disease-associated STR gene (n = 37) is shown as a separate panel and two alleles are shown for each individual, excepting chrX-linked genes for male individuals, where only one copy is present. Clinically affected and premutation-carrier individuals are marked. Basic summary statistics for each gene are shown below.

**bi**

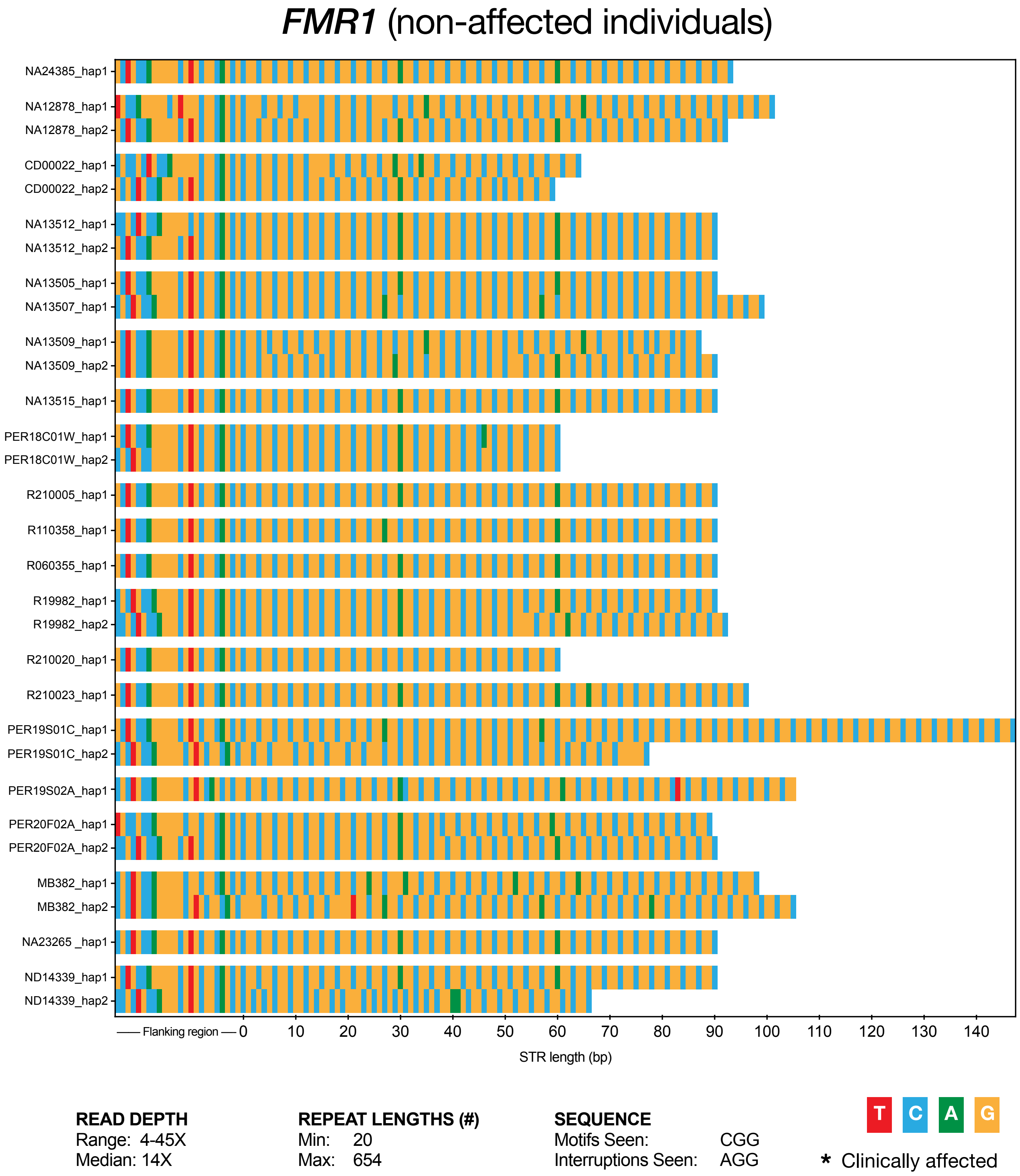
*FMR1* (non-affected individuals)
NA24385_hap1
NA12878_hap1
NA12878_hap2
CD00022_hap1
CD00022_hap2
NA13512_hap1
NA13512_hap2
NA13505_hap1
NA13507_hap1
NA13509_hap1
NA13509_hap2
NA13515_hap1
PER18C01W_hap1
PER18C01W_hap2
R210005_hap1
R110358_hap1
R060355_hap1
R19982_hap1
R19982_hap2
R210020_hap1
R210023_hap1
PER19S01C_hap1
PER19S01C_hap2
PER19S02A_hap1
PER20F02A_hap1
PER20F02A_hap2
MB382_hap1
MB382_hap2
NA23265 _hap1
ND14339_hap1
ND14339_hap2
Flanking region
0
10
20
30
40
50
60
70
80
90
100
110
120
130
140
STR length (bp)
READ DEPTH
Range: 4-45X
Median: 14X
REPEAT LENGTHS (#)
Min: 20
Max: 654
SEQUENCE
Motifs Seen: CGG
Interruptions Seen: AGG
T
C
A
G
* Clinically affected

**ci**

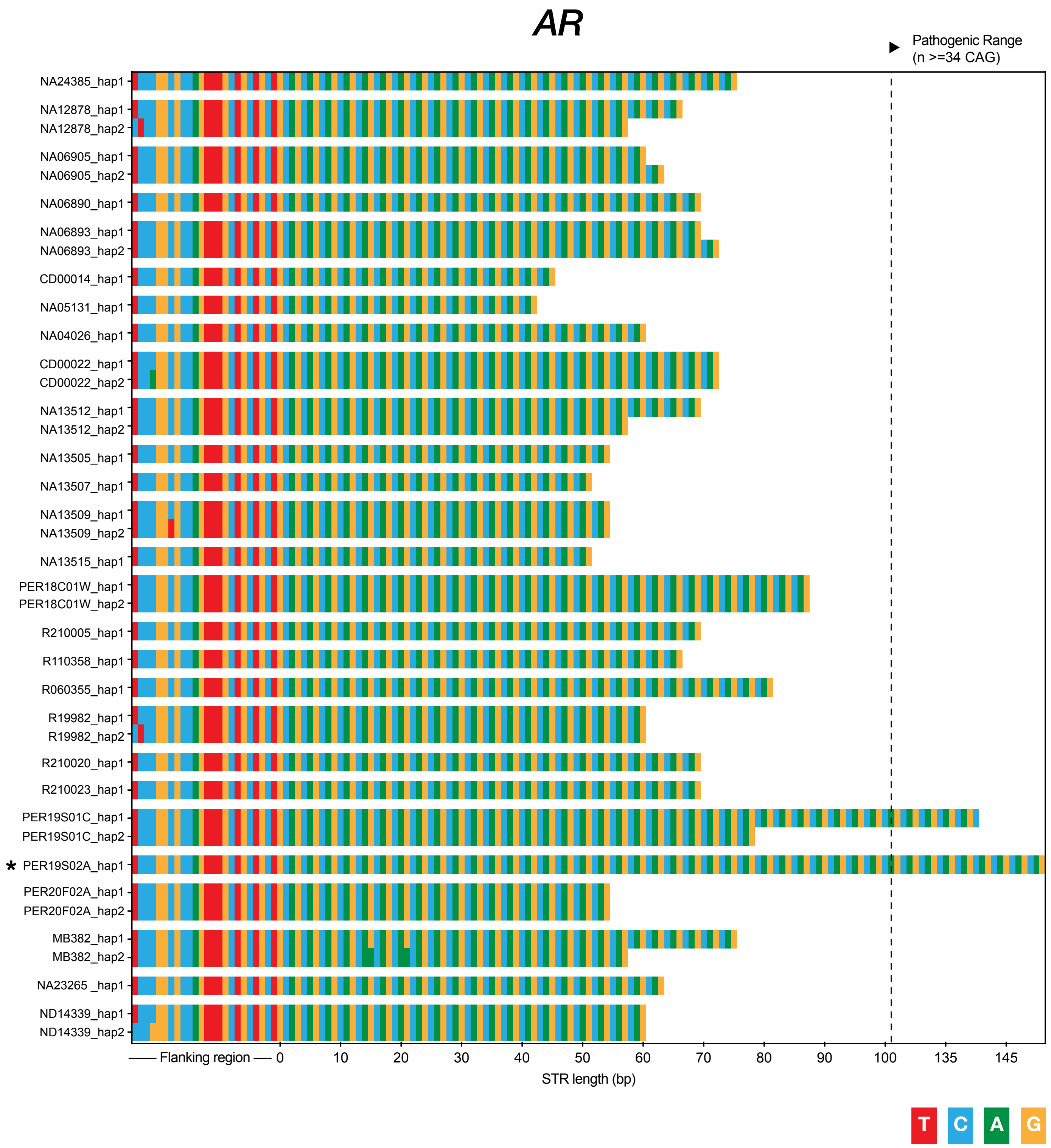


**READ DEPTH**
Range: 4-29x
Median:13.6x

**REPEAT LENGTHS (#)**
Min: 14
Max: 42

**SEQUENCE**
Motifs Seen: CAG
Interruptions Seen: CAA

di

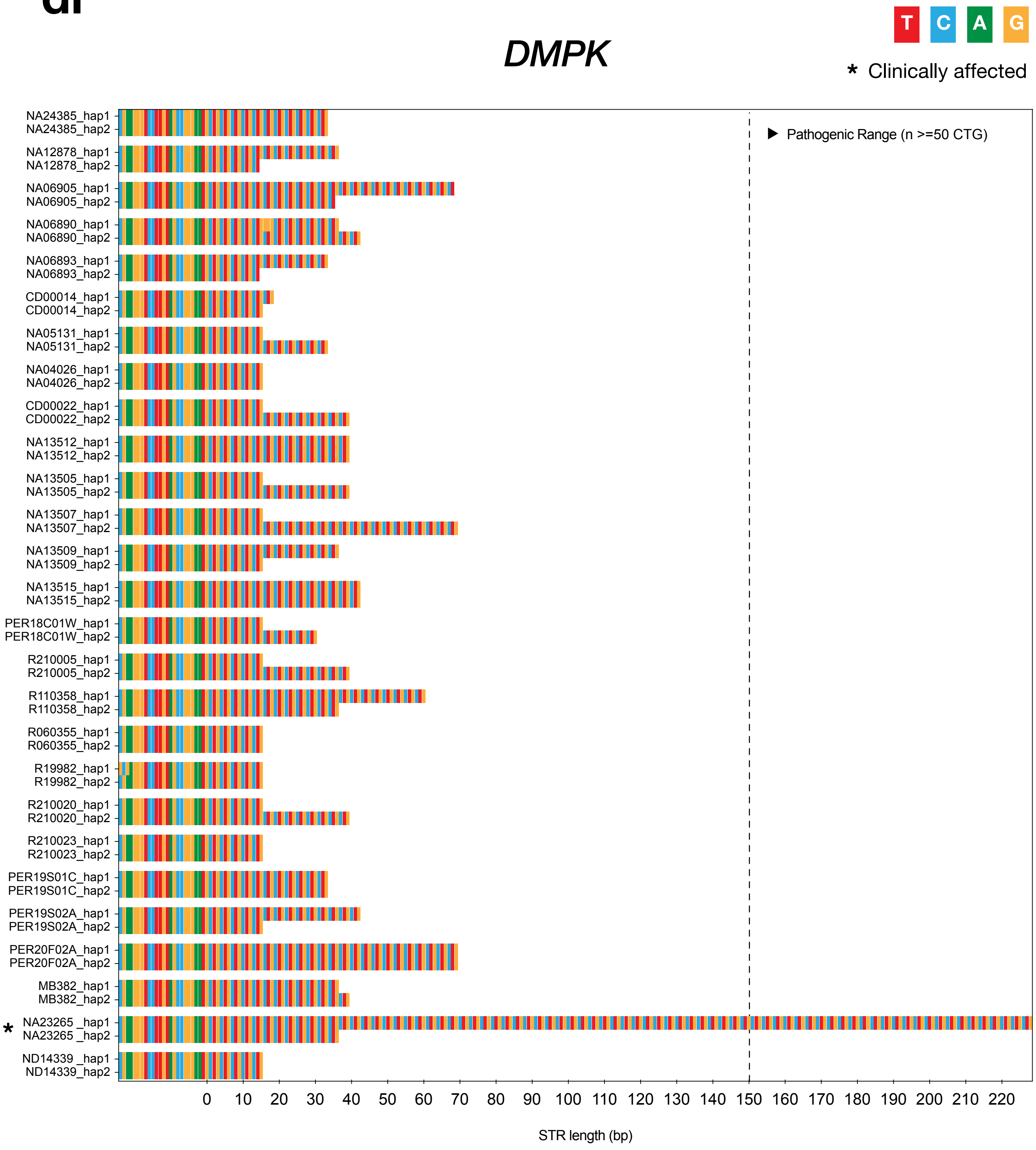


**READ DEPTH**
Range: 7-44X
Median: 17X

**REPEAT LENGTHS (#)**
Min: 5
Max: 76

**SEQUENCE**
Motifs Seen: CTG
Interruptions Seen:

**ei**

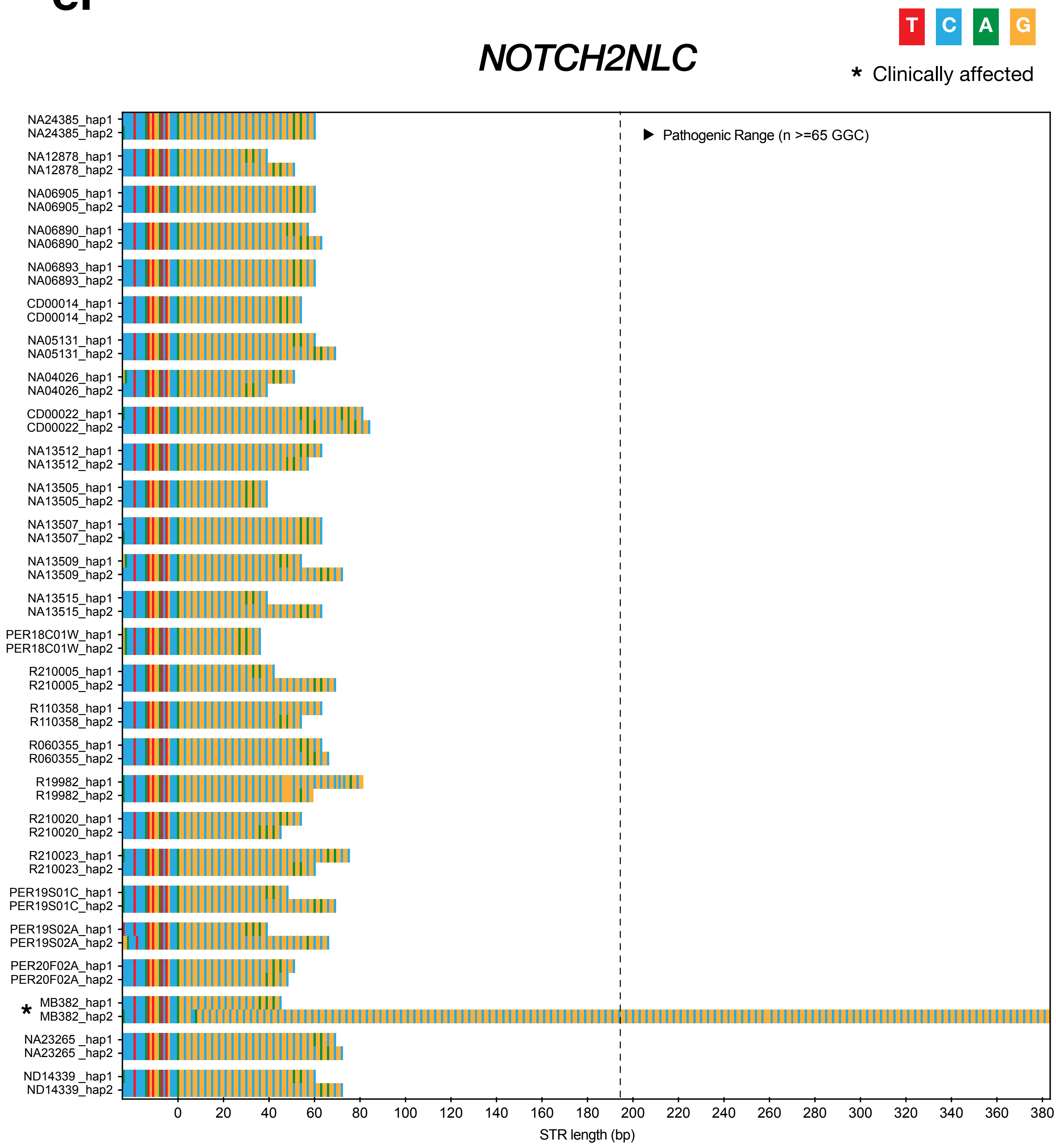


**READ DEPTH**
Range: 5-43x
Median: 20x

**REPEAT LENGTHS (#)**
Min: 12
Max: 127

**SEQUENCE**
Motifs Seen: GGC
Interruptions Seen: GGA, GGG

fi

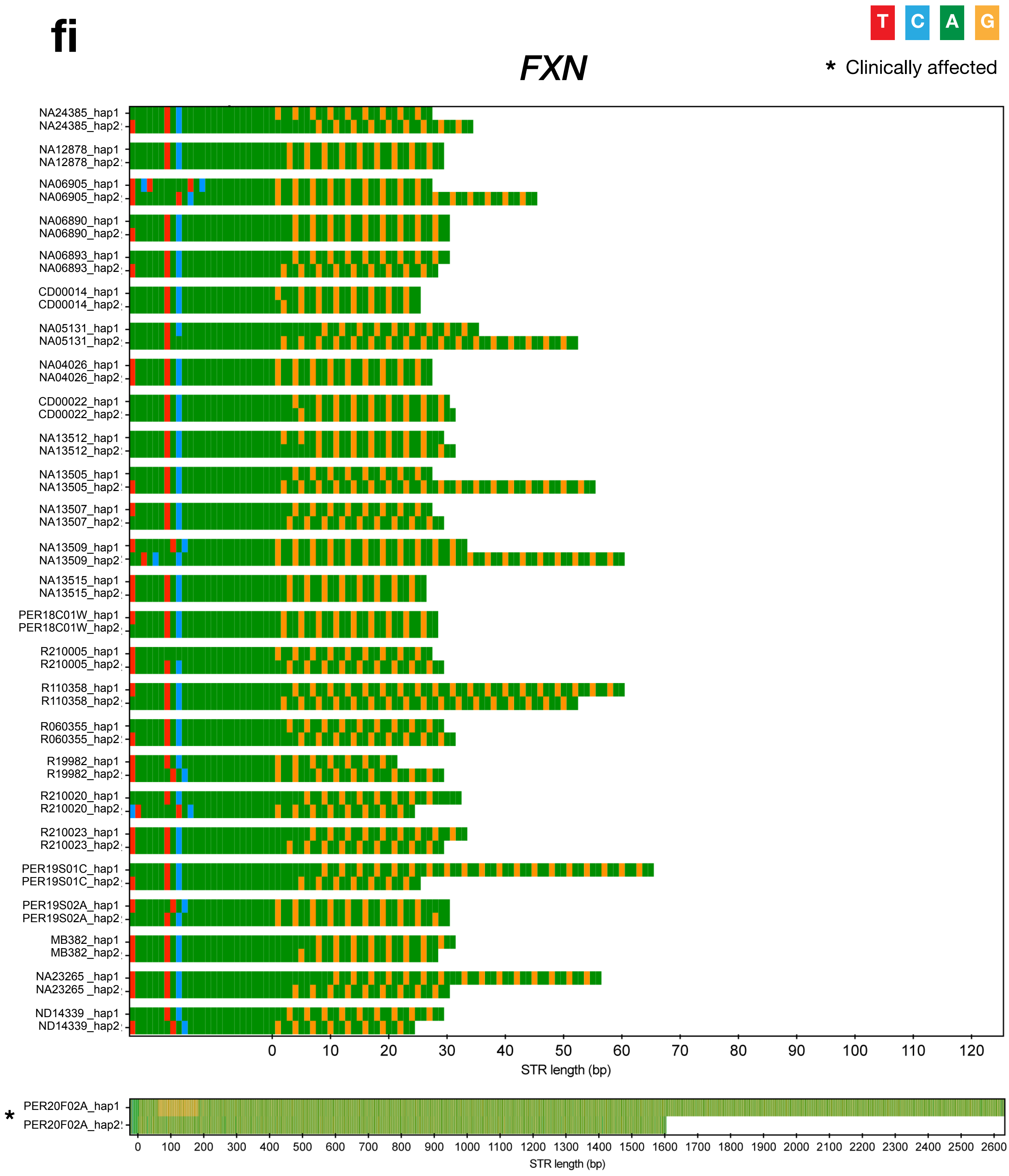


**READ DEPTH**
Range: 5-41x
Median: 22x

**REPEAT LENGTHS (#)**
Min: 7
Max: 812

**SEQUENCE**
Motifs Seen: GAA
Interruptions Seen:

# gi

T C A G

### *C9orf72*

* Clinically affected

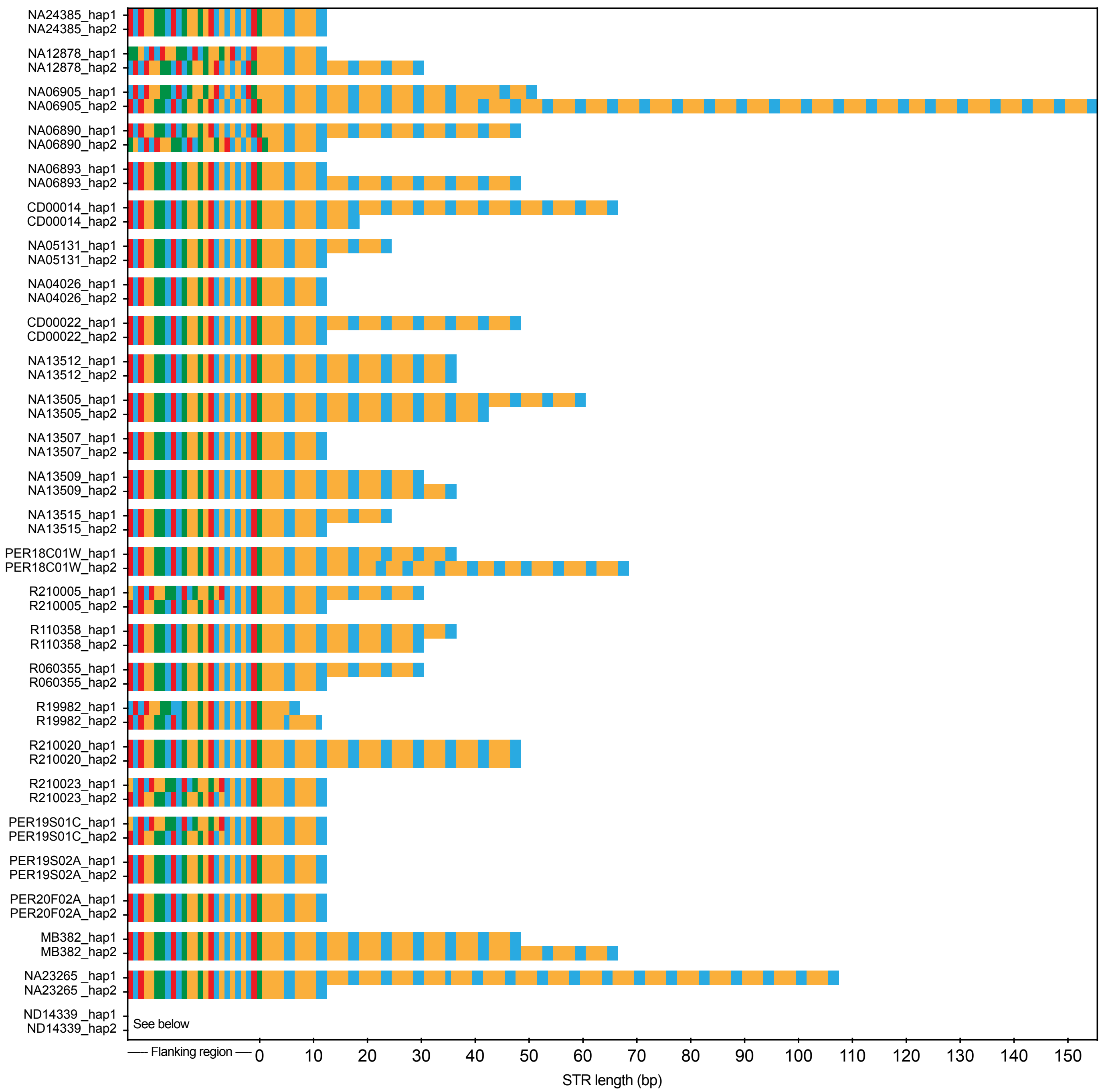


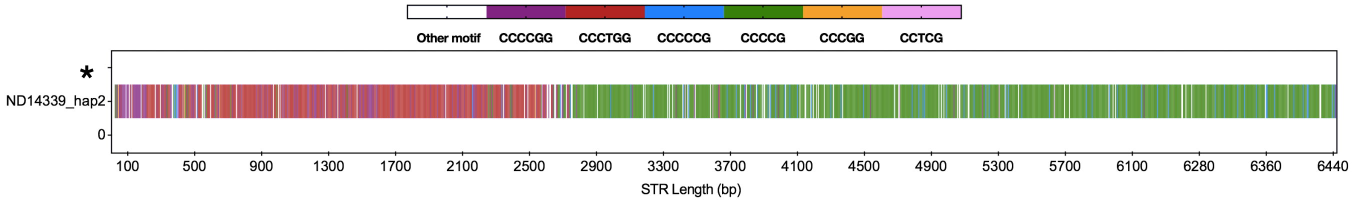


**READ DEPTH**
Range: 6-43x
Median: 21x

**REPEAT LENGTHS (#)**
Min: 2
Max: 1070

**SEQUENCE**
Motifs Seen: GGGGCC, ?CCCTGG, ?CCCGG
Interruptions Seen:

hi

### *STARD7*

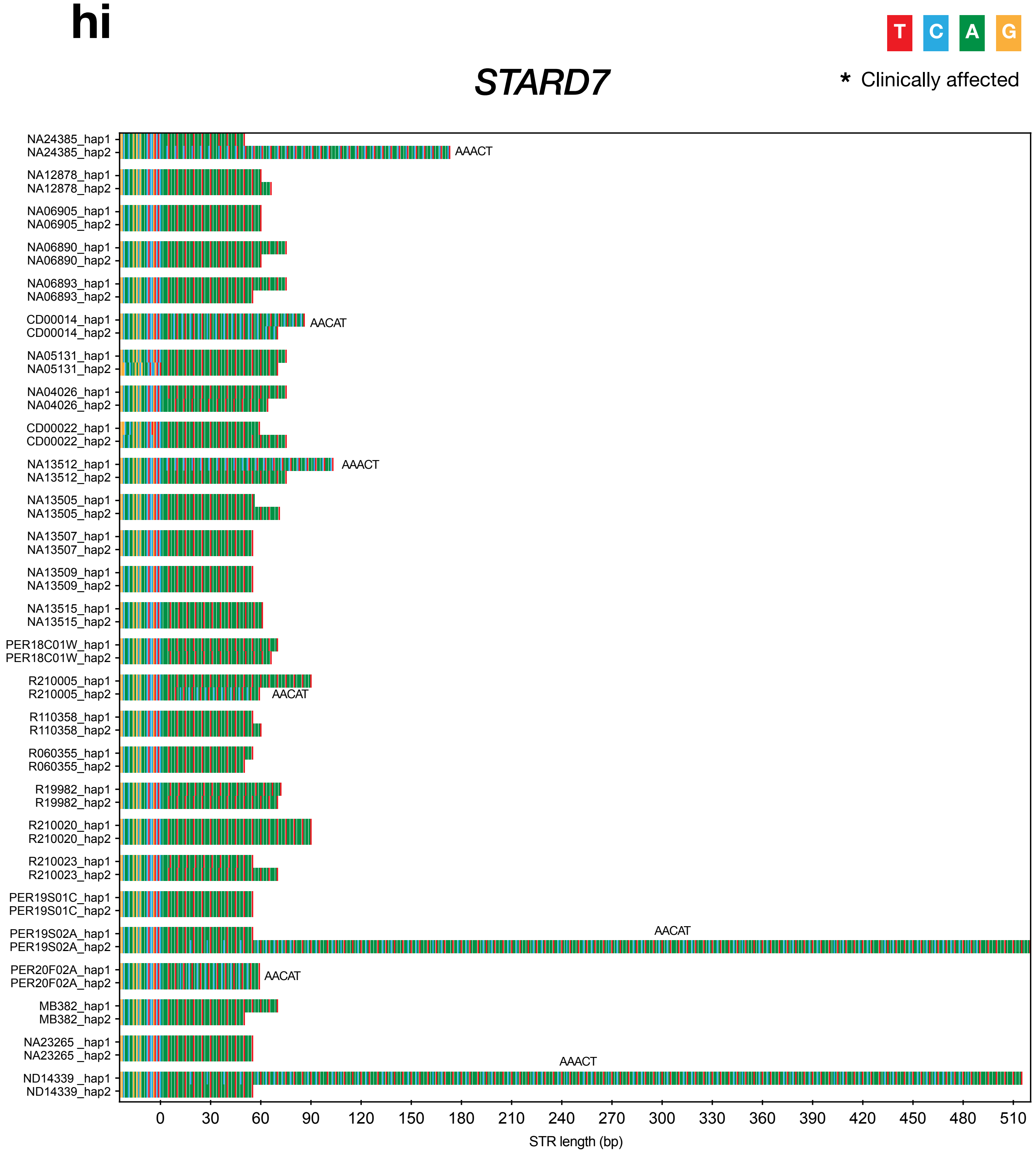


**READ DEPTH**
Range: 9-35x
Median: 17x

**REPEAT LENGTHS (#)**
Min: 10
Max: 102

**SEQUENCE**
Motifs Seen: AAAAT, AACAT, AAACT
Interruptions Seen:

**ii**

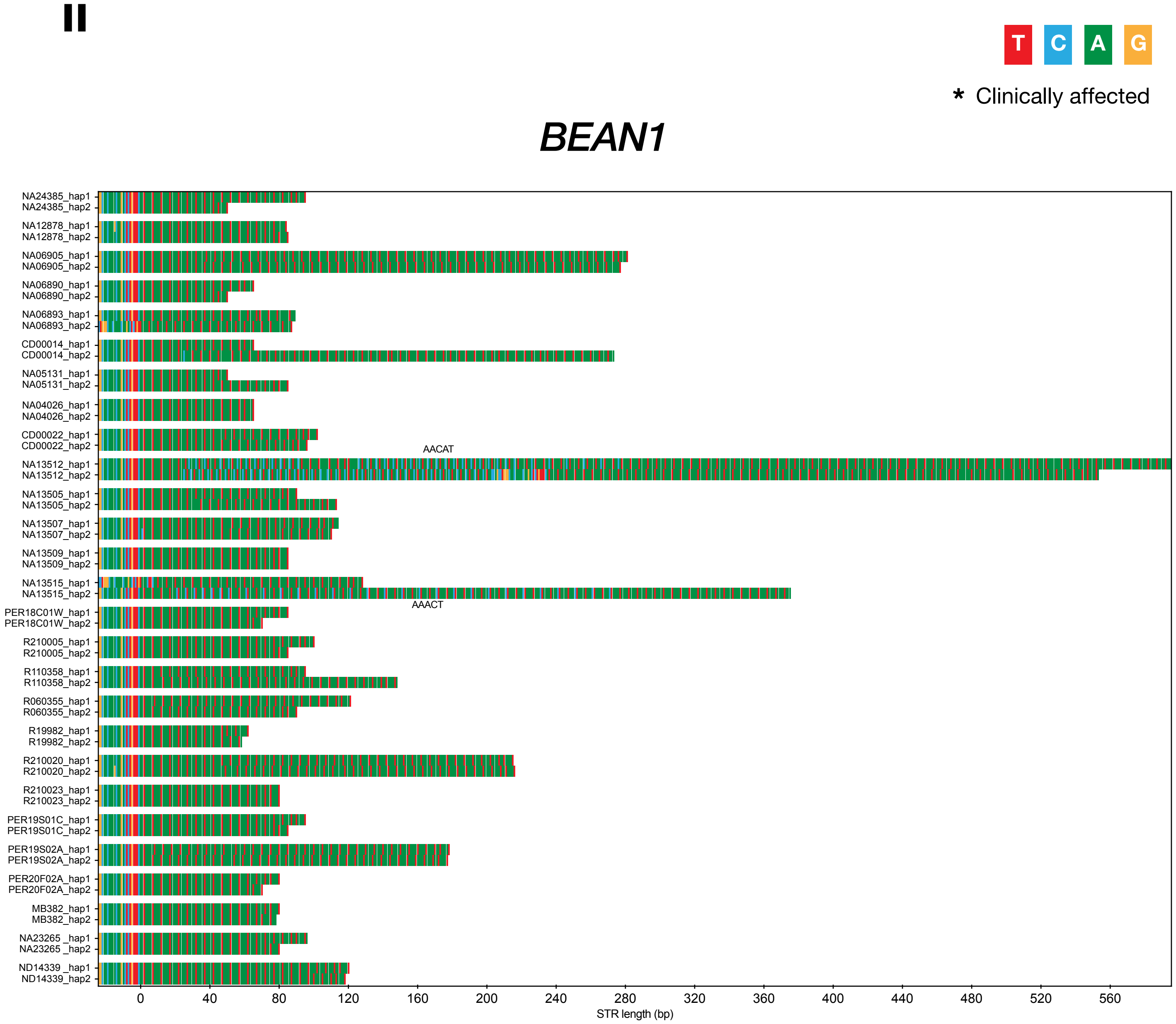


**READ DEPTH**
Range: 4-36x
Median: 20x

**REPEAT LENGTHS (#)**
Min: 10
Max: 119

**SEQUENCE**
Motifs Seen: AAAAT, AACAT, AAACT
Interruptions Seen:

**ji**

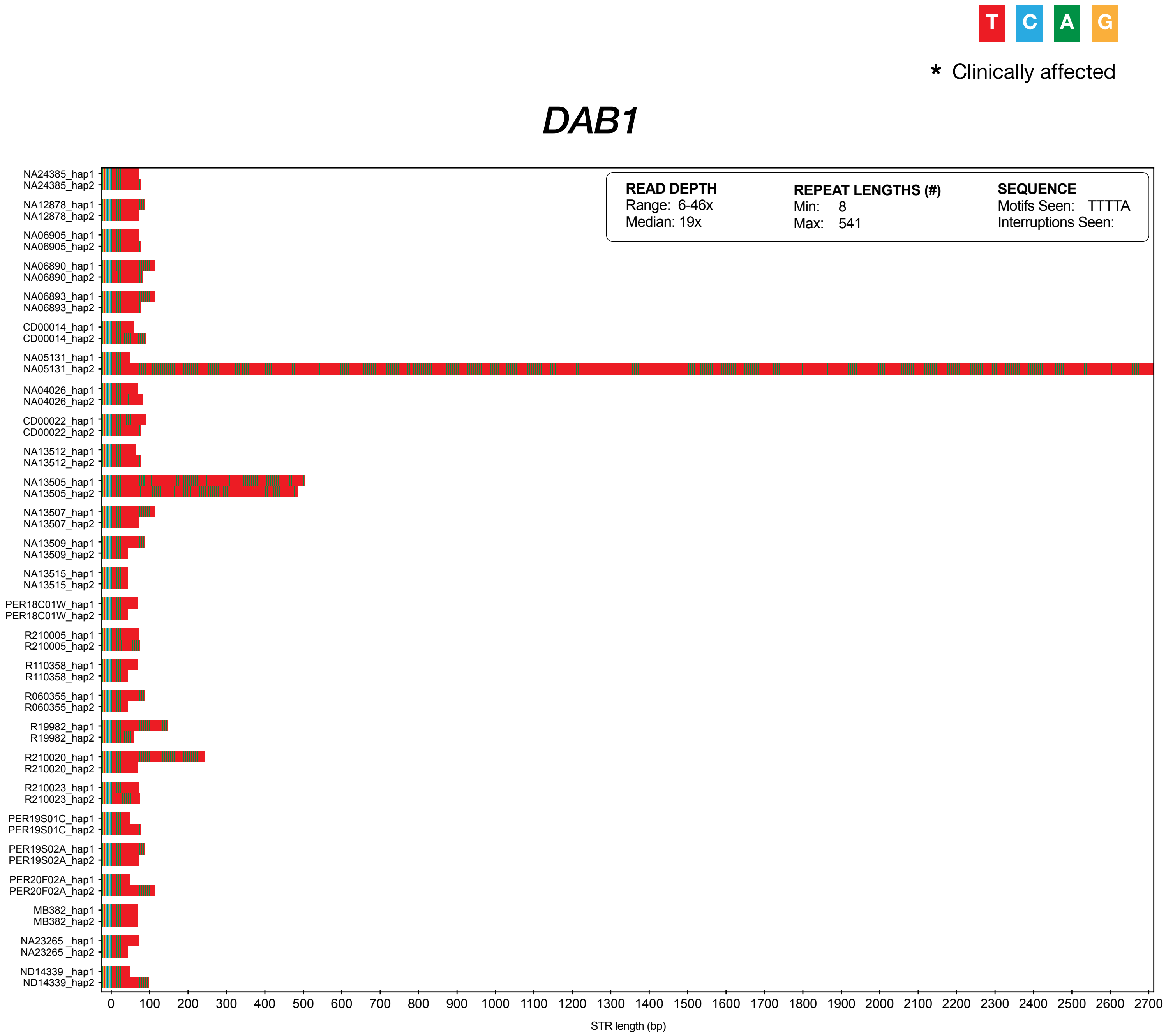
T
C
A
G
* Clinically affected
DAB1
READ DEPTH
Range: 6-46x
Median: 19x
REPEAT LENGTHS (#)
Min: 8
Max: 541
SEQUENCE
Motifs Seen: TTTTA
Interruptions Seen:
NA24385_hap1
NA24385_hap2
NA12878_hap1
NA12878_hap2
NA06905_hap1
NA06905_hap2
NA06890_hap1
NA06890_hap2
NA06893_hap1
NA06893_hap2
CD00014_hap1
CD00014_hap2
NA05131_hap1
NA05131_hap2
NA04026_hap1
NA04026_hap2
CD00022_hap1
CD00022_hap2
NA13512_hap1
NA13512_hap2
NA13505_hap1
NA13505_hap2
NA13507_hap1
NA13507_hap2
NA13509_hap1
NA13509_hap2
NA13515_hap1
NA13515_hap2
PER18C01W_hap1
PER18C01W_hap2
R210005_hap1
R210005_hap2
R110358_hap1
R110358_hap2
R060355_hap1
R060355_hap2
R19982_hap1
R19982_hap2
R210020_hap1
R210020_hap2
R210023_hap1
R210023_hap2
PER19S01C_hap1
PER19S01C_hap2
PER19S02A_hap1
PER19S02A_hap2
PER20F02A_hap1
PER20F02A_hap2
MB382_hap1
MB382_hap2
NA23265 _hap1
NA23265 _hap2
ND14339 _hap1
ND14339_hap2
0 100 200 300 400 500 600 700 800 900 1000 1100 1200 1300 1400 1500 1600 1700 1800 1900 2000 2100 2200 2300 2400 2500 2600 2700
STR length (bp)

ki

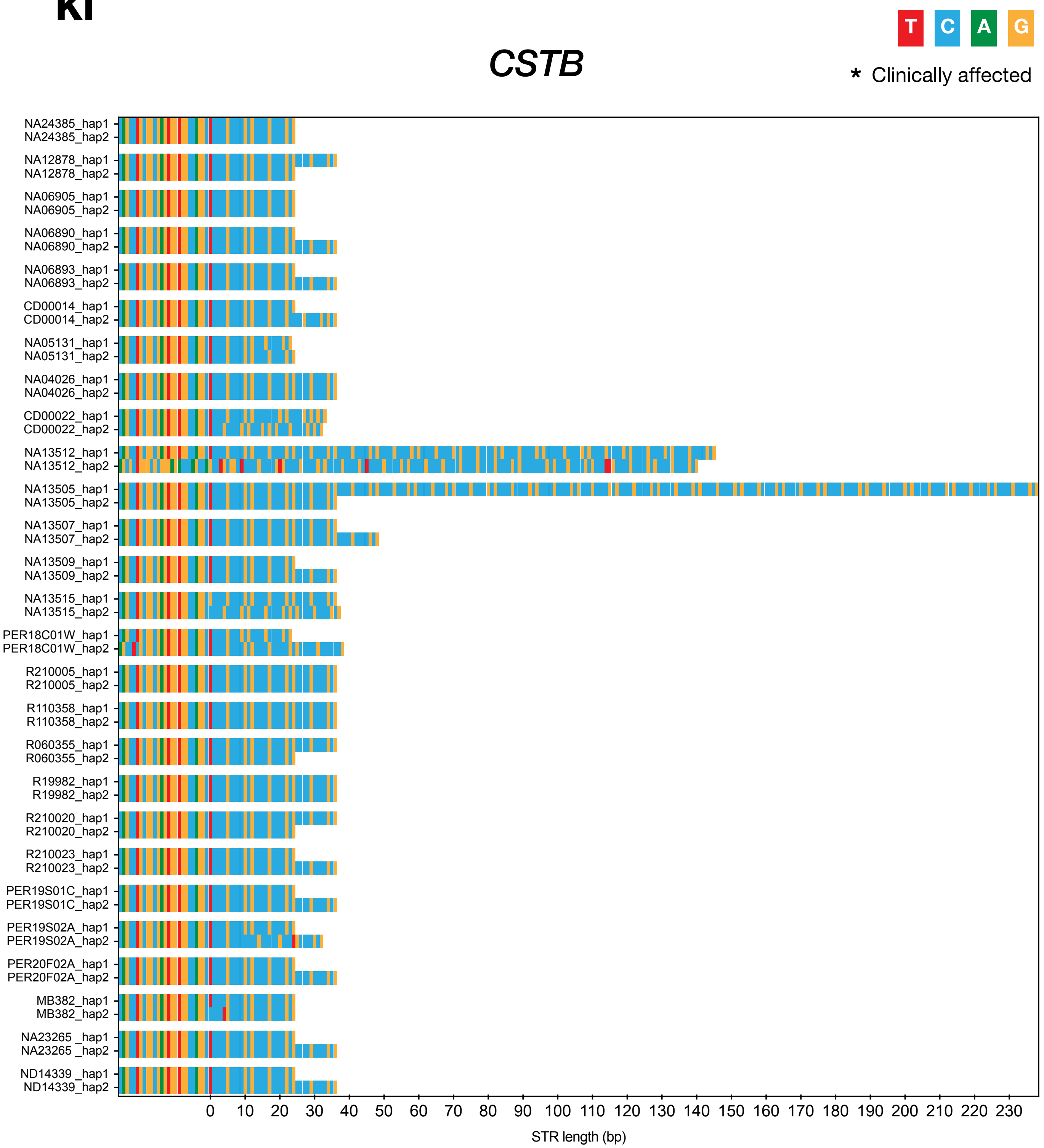


**READ DEPTH**
Range: 7-41x
Median:18x

**REPEAT LENGTHS (#)**
Min: 2
Max: 20

**SEQUENCE**
Motifs Seen: CCCCGCCCCGCG
Interruptions Seen:

**Ii**

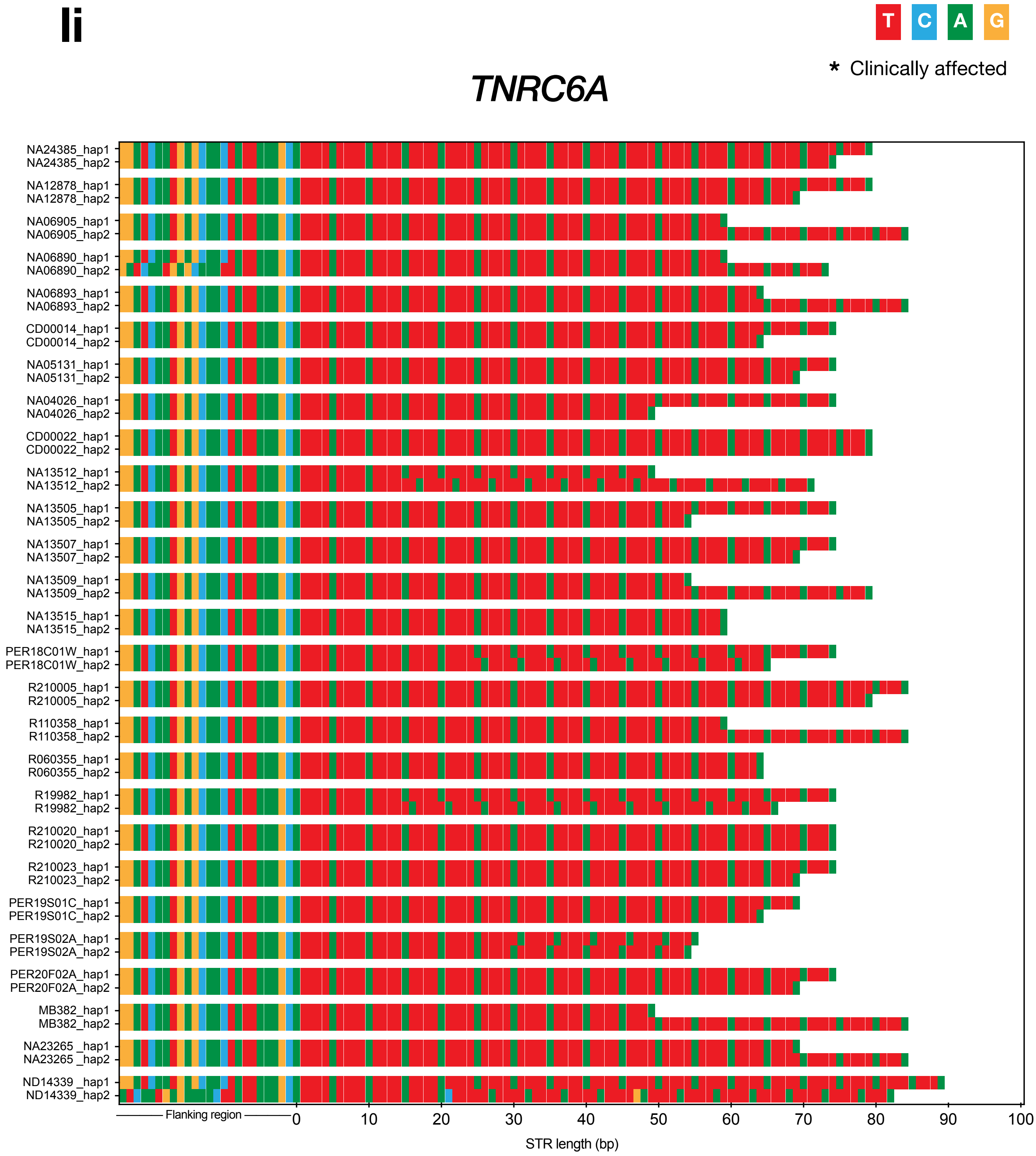


**READ DEPTH**
Range: 7-44x
Median: 18x

**REPEAT LENGTHS (#)**
Min: 9
Max: 18

**SEQUENCE**
Motifs Seen: TTTTA
Interruptions Seen: TTTTTA

# mi

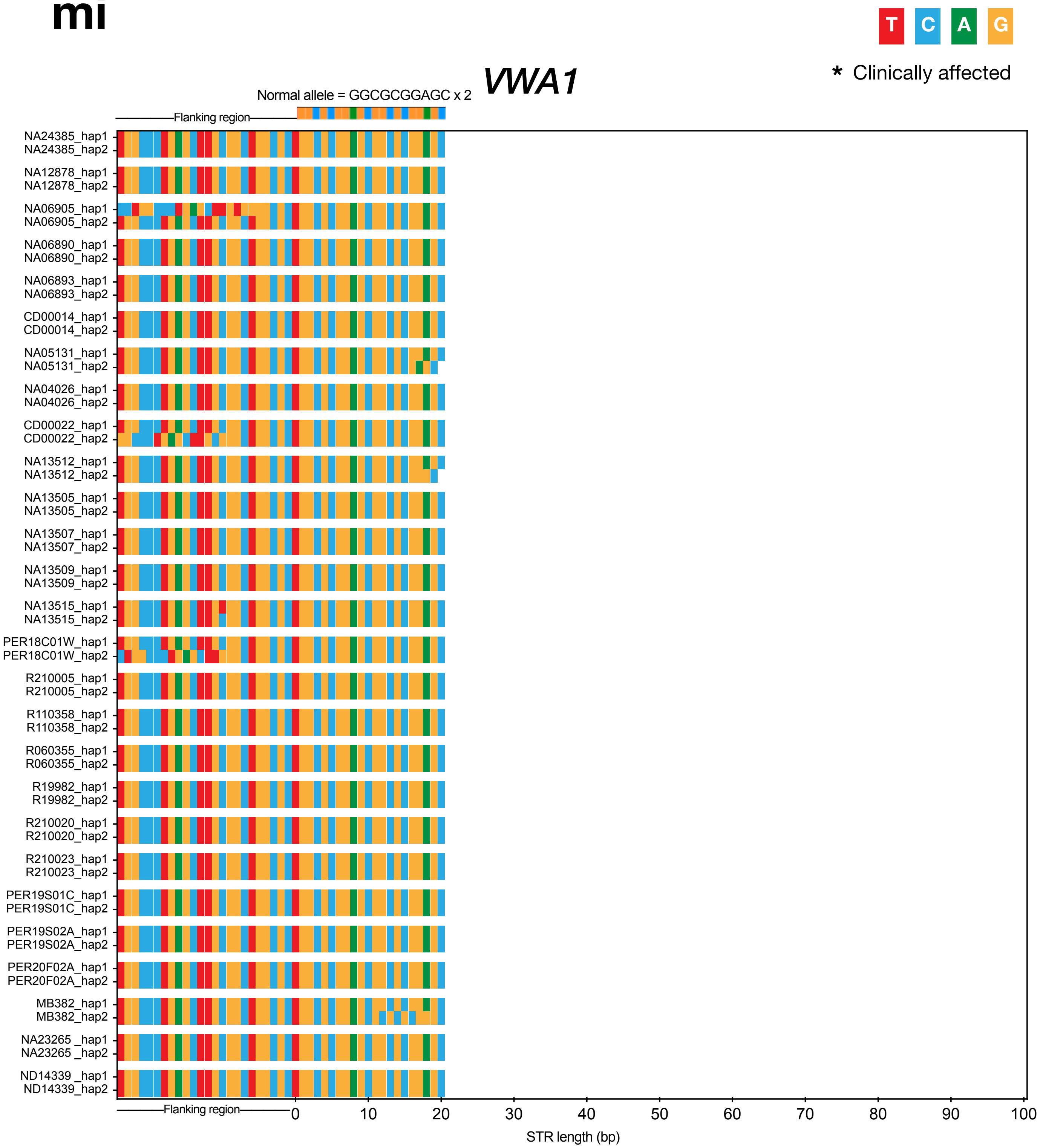


**READ DEPTH**
Range: 5-42x
Median: 17x

**REPEAT LENGTHS (#)**
Min: 2
Max: 2

**SEQUENCE**
Motifs Seen: GGCGCGGAGC
Interruptions Seen:

# ni

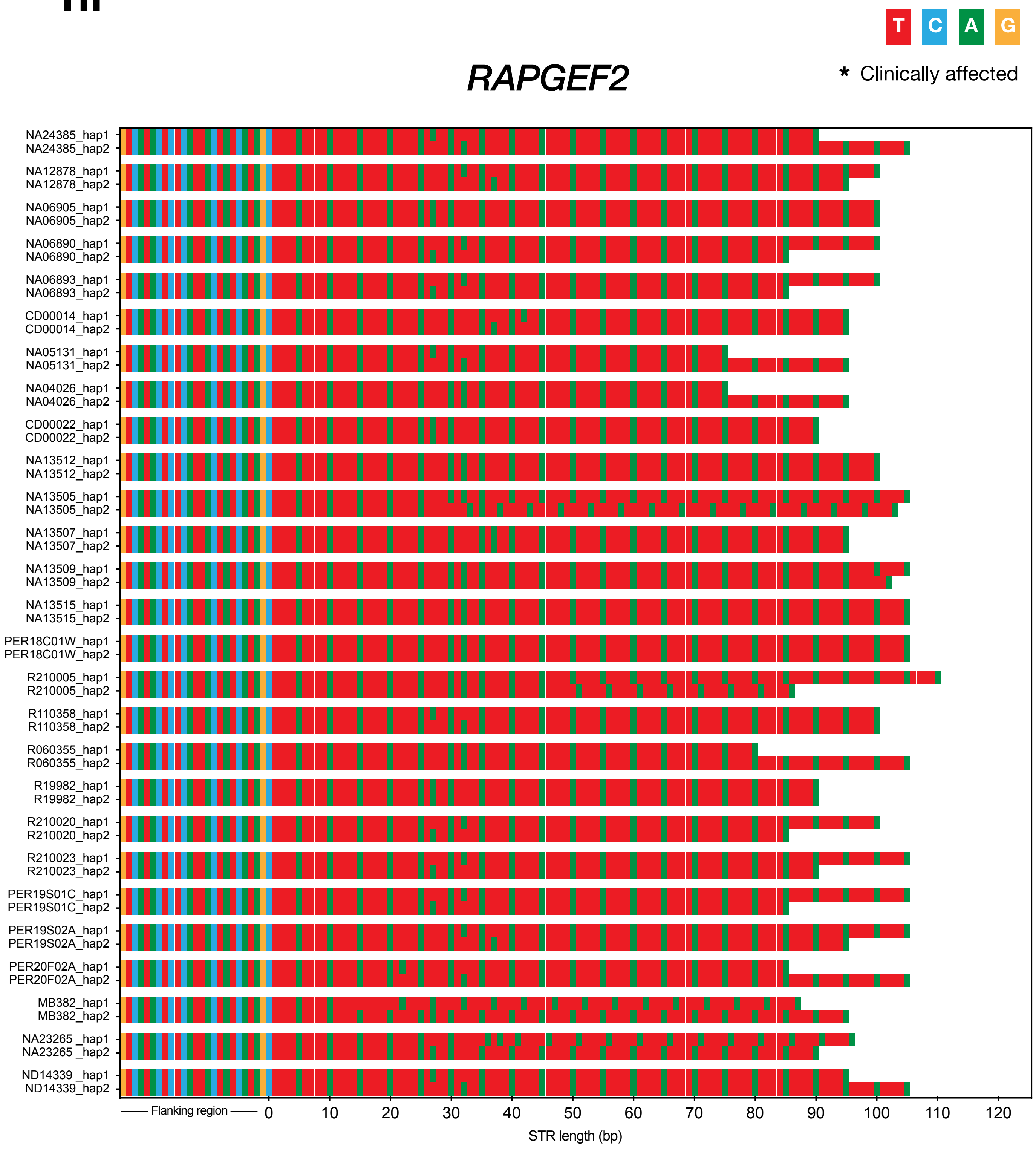


**READ DEPTH**
Range: 6-45x
Median: 20x

**REPEAT LENGTHS (#)**
Min: 15
Max: 22

**SEQUENCE**
Motifs Seen: TTTTA
Interruptions Seen: TATTA, TTTTTA

oi

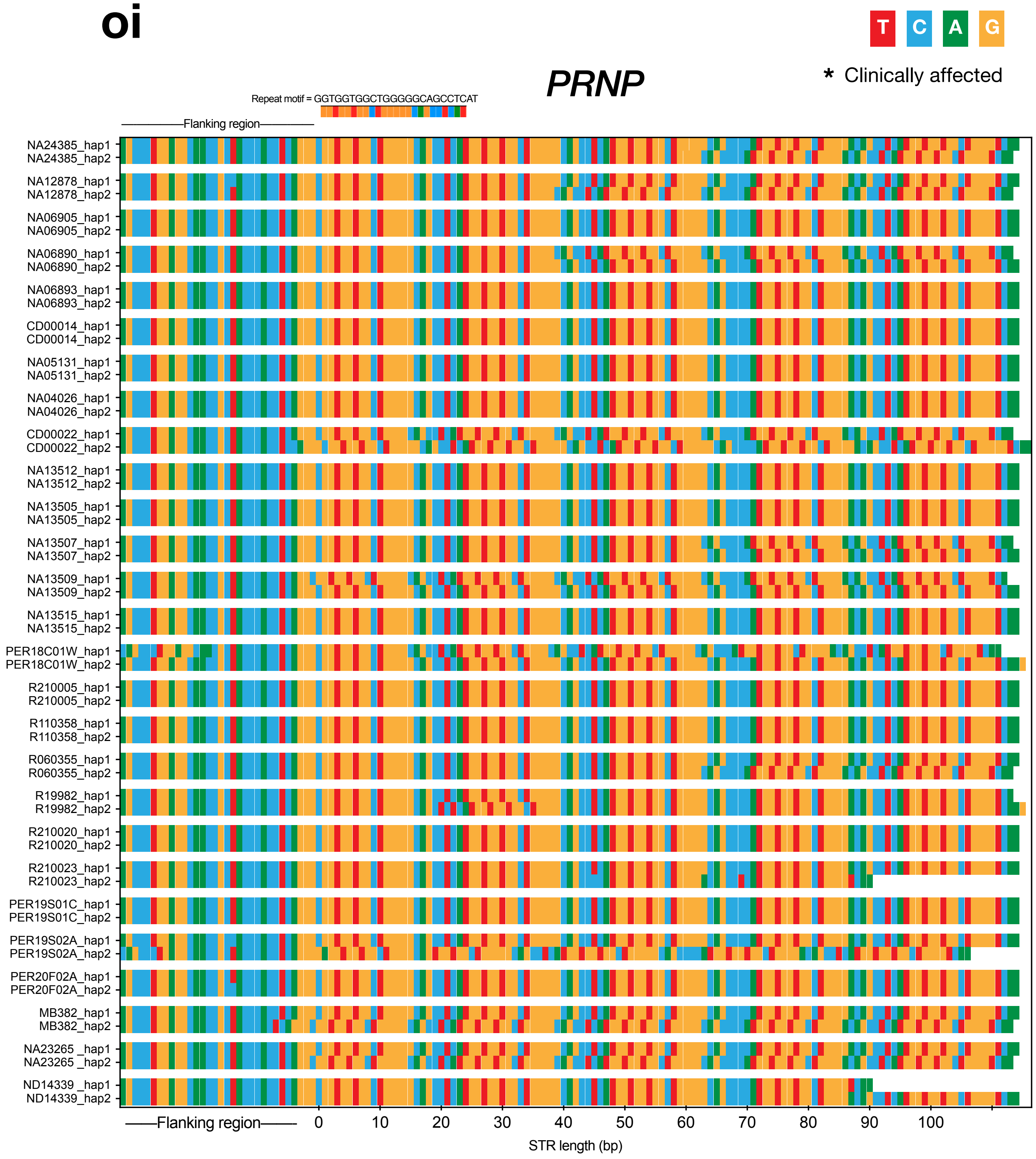


| READ DEPTH | REPEAT LENGTHS (#) | SEQUENCE | |
|---|---|---|---|
| Range: 9-42x | Min: 4 | Motifs Seen: | GGTGGTGGCTGGGGGCAGCCTCAT |
| Median: 19x | Max: 4 | Interruptions Seen: | GGTGGTGGCTGGGGGCAGCCCCAT |

pi

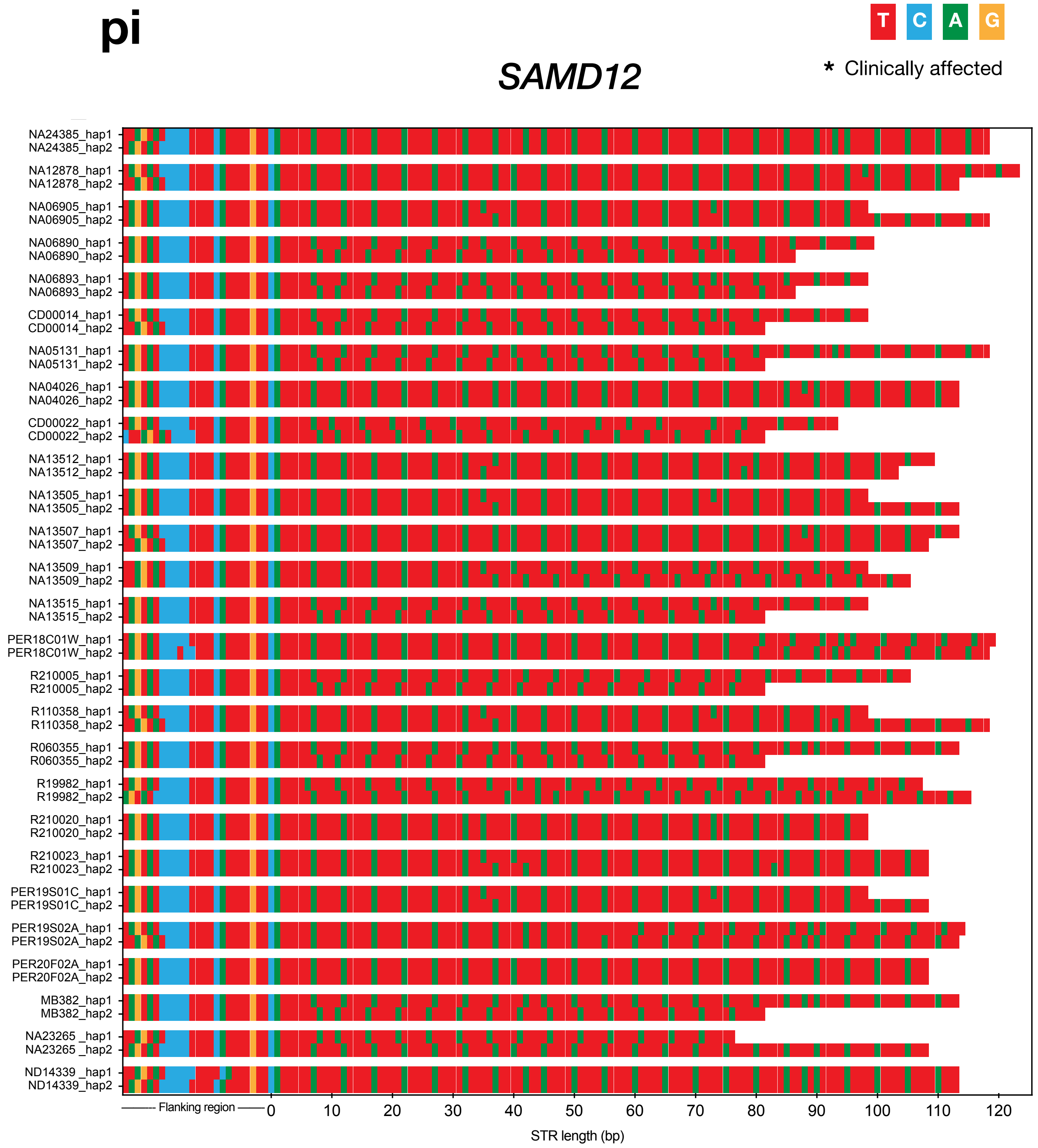


**READ DEPTH**
Range: 10-42x
Median: 21x

**REPEAT LENGTHS (#)**
Min: 14
Max: 24

**SEQUENCE**
Motifs Seen: TTTTA
Interruptions Seen: TTA, TTATA, TTTTTA

qi

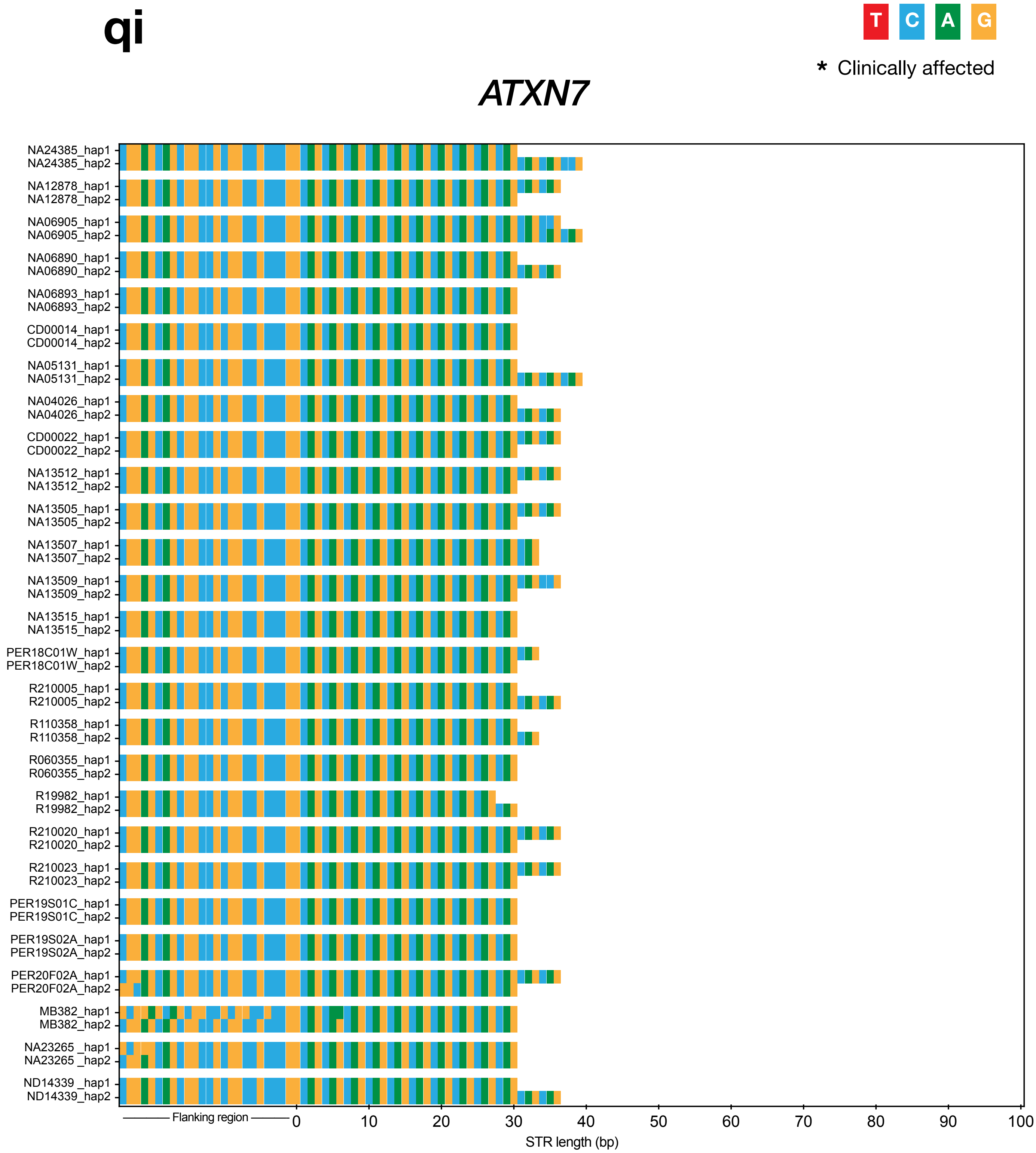


**READ DEPTH**
Range: 7-43x
Median: 21x

**REPEAT LENGTHS (#)**
Min: 9
Max: 13

**SEQUENCE**
Motifs Seen: CAG
Interruptions Seen: CAA

ri

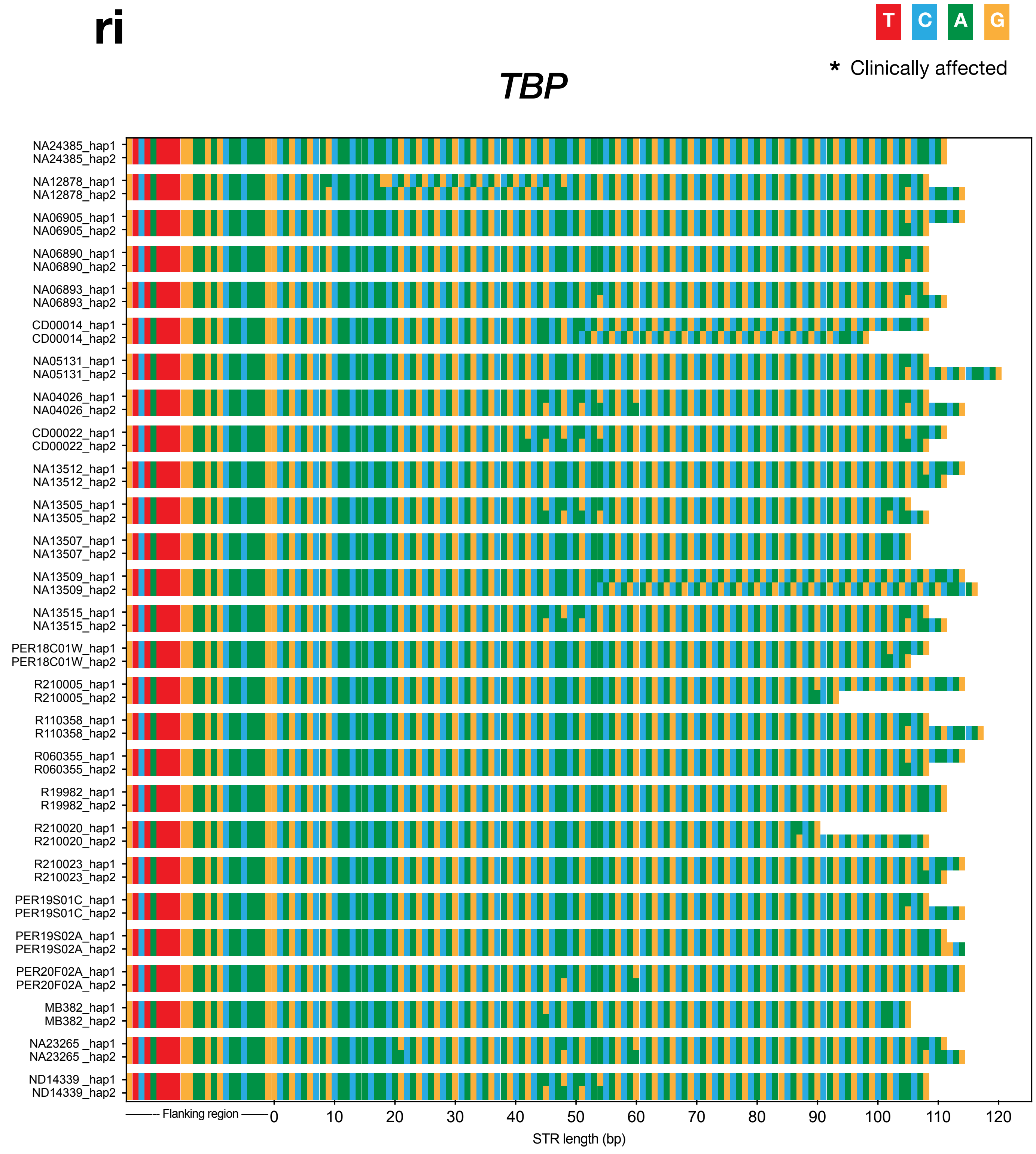


**READ DEPTH**
Range: 7-44x
Median: 19x

**REPEAT LENGTHS (#)**
Min: 30
Max: 40

**SEQUENCE**
Motifs Seen: CAG
Interruptions Seen: CAA

si

*PPP2R2B*

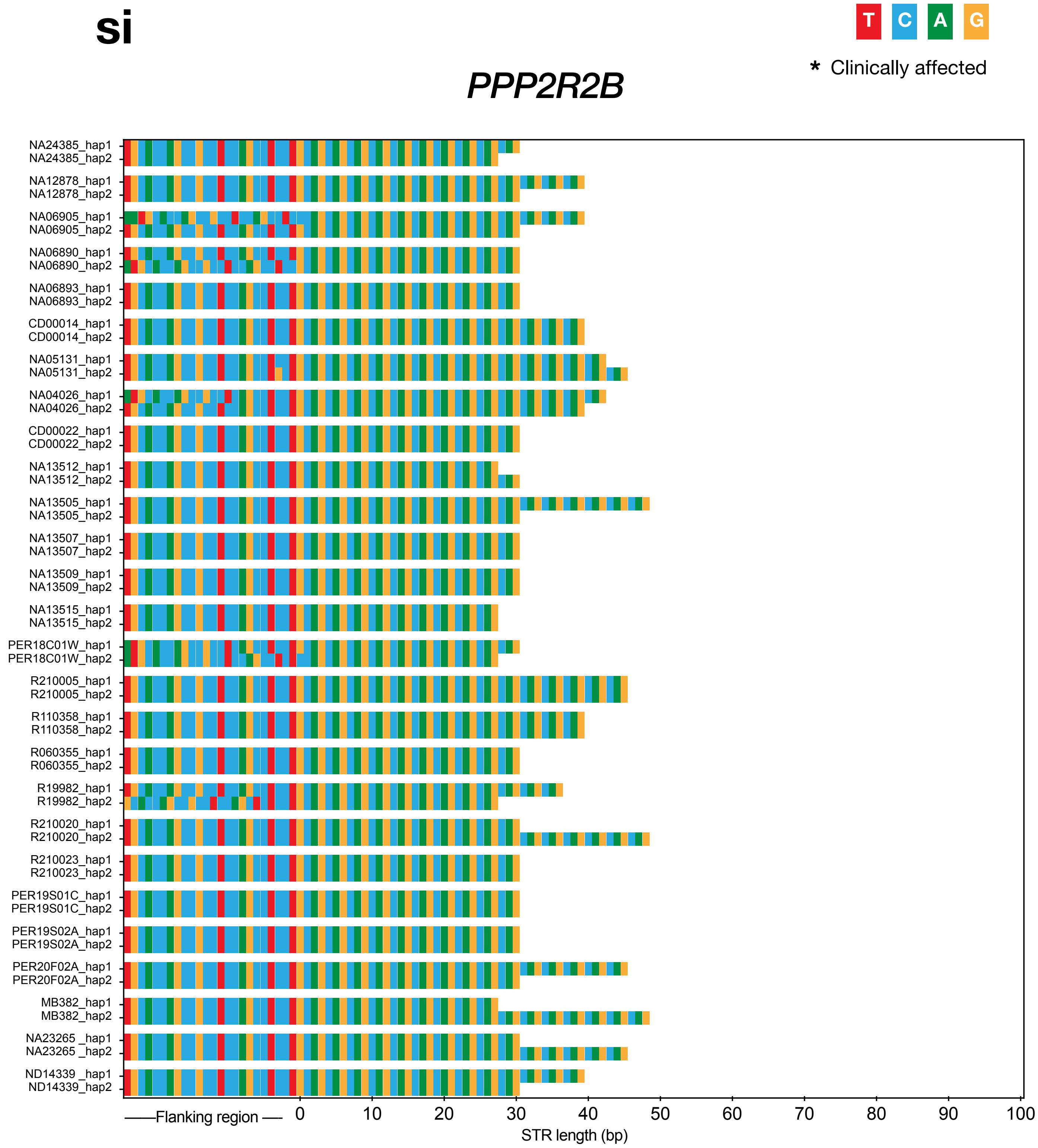


**READ DEPTH**
Range: 6-35x
Median: 20x

**REPEAT LENGTHS (#)**
Min: 9
Max: 16

**SEQUENCE**
Motifs Seen: CAG
Interruptions Seen:

ti

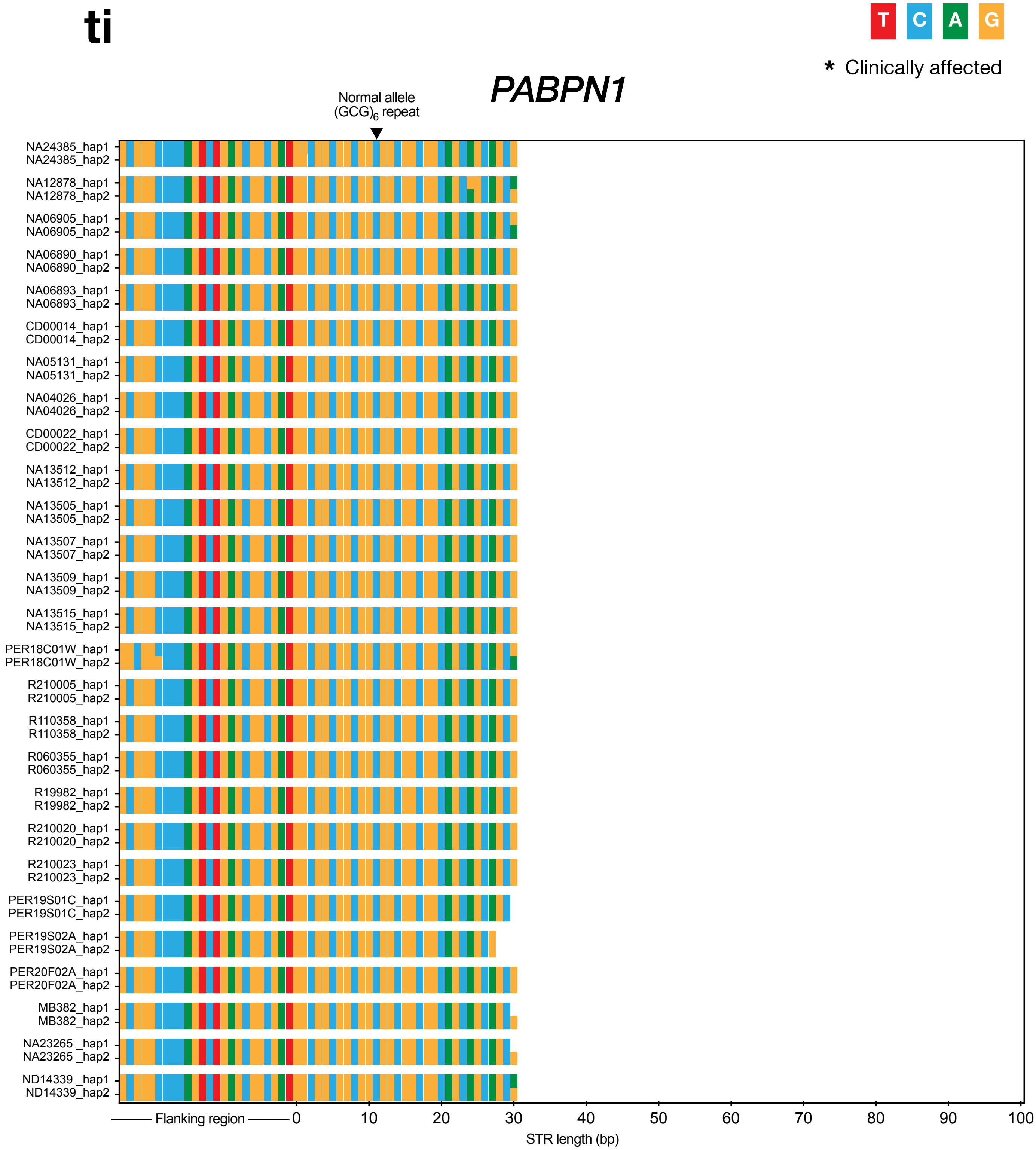


**READ DEPTH**
Range: 6-41x
Median: 19x

**REPEAT LENGTHS (#)**
Min: 6
Max: 6

**SEQUENCE**
Motifs Seen: GCG
Interruptions Seen: GCA

ui

### *NUTM2B*

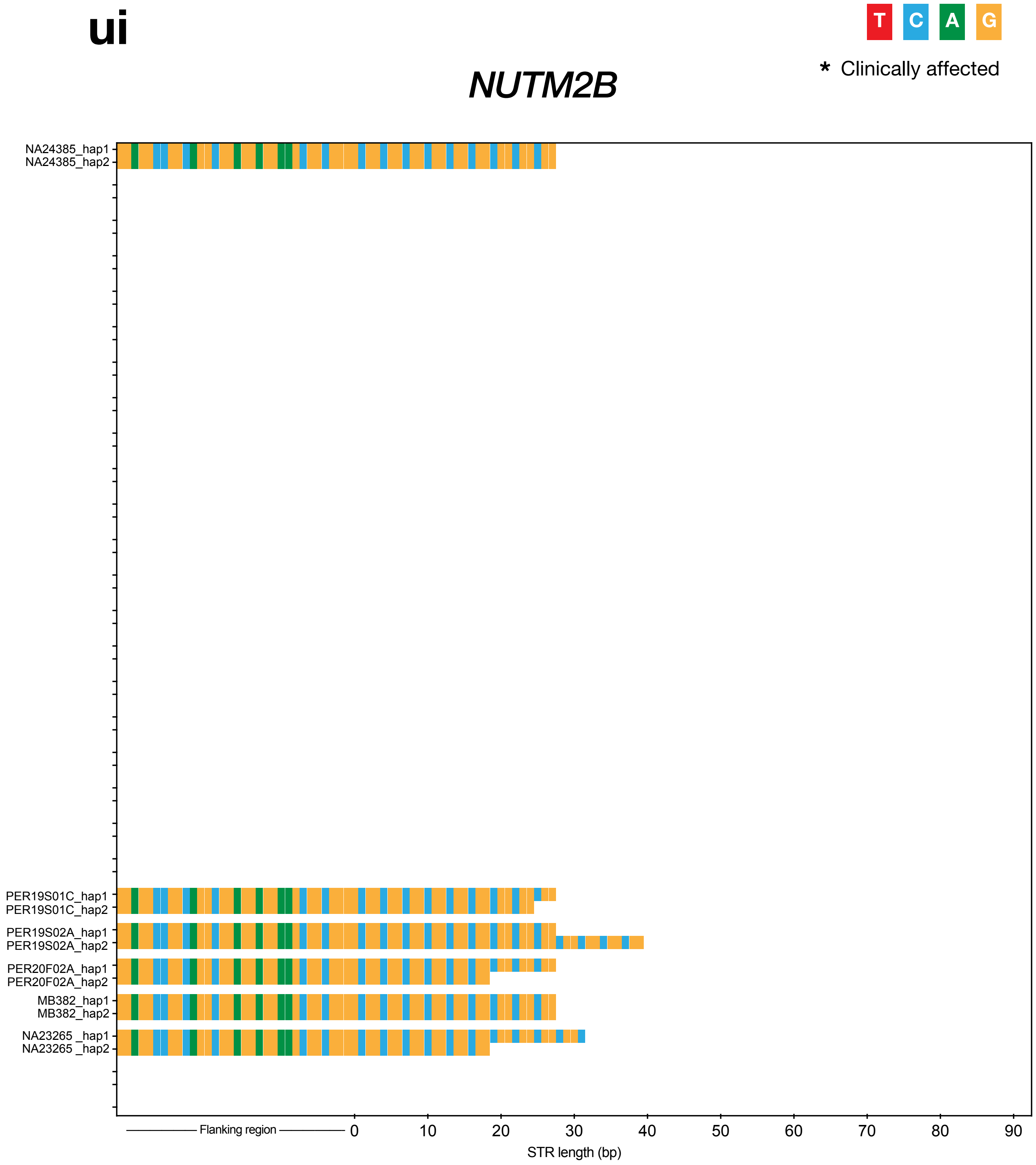


**READ DEPTH**
Range: 14-43x
Median: 20.5x

**REPEAT LENGTHS (#)**
Min: 6
Max: 13

**SEQUENCE**
Motifs Seen: GGC
Interruptions Seen:

**vi**

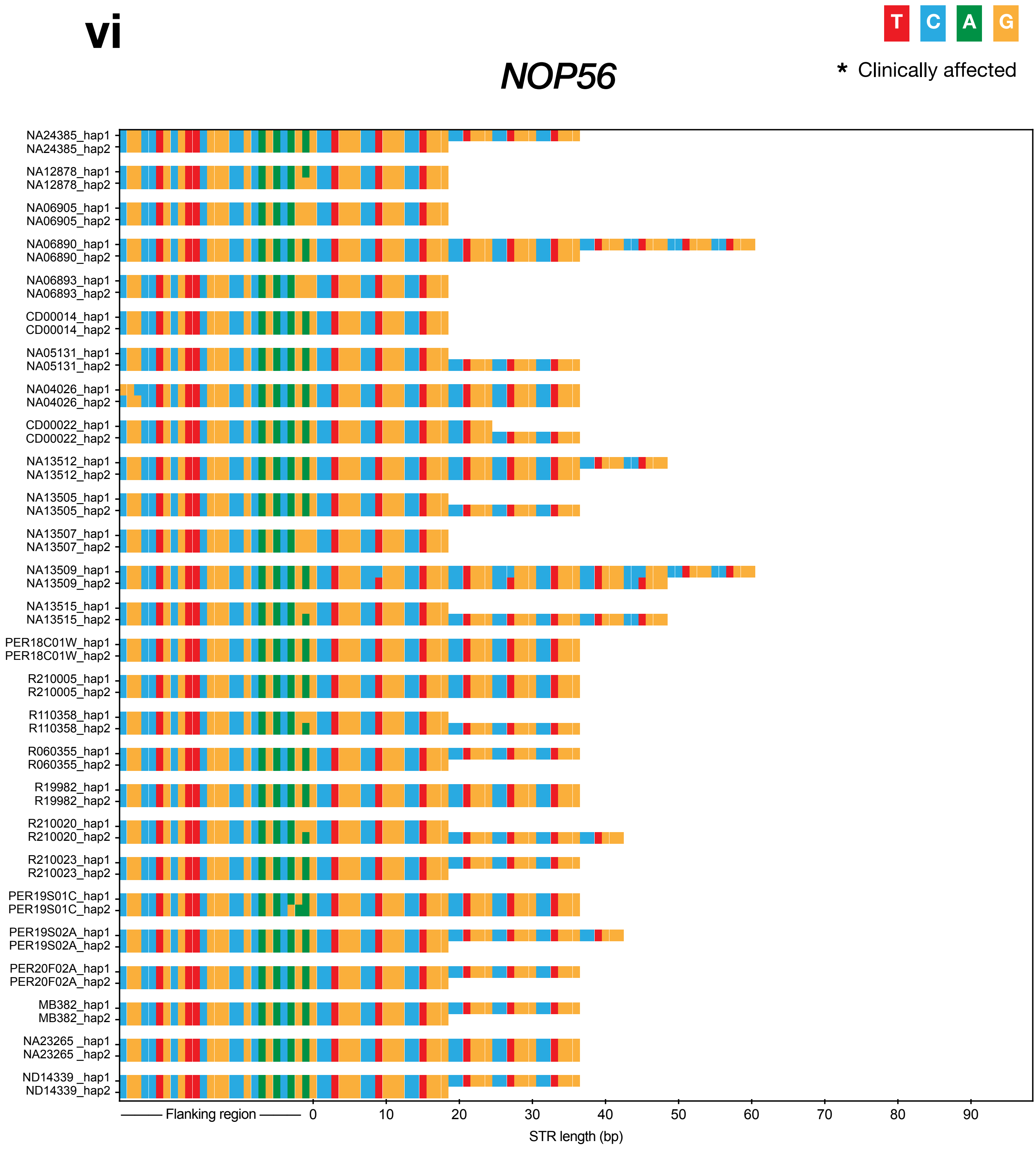


**READ DEPTH**
Range: 6-38x
Median: 20x

**REPEAT LENGTHS (#)**
Min: 3
Max: 10

**SEQUENCE**
Motifs Seen: CCTGGG
Interruptions Seen: CCCGGG

### *MARCHF6*

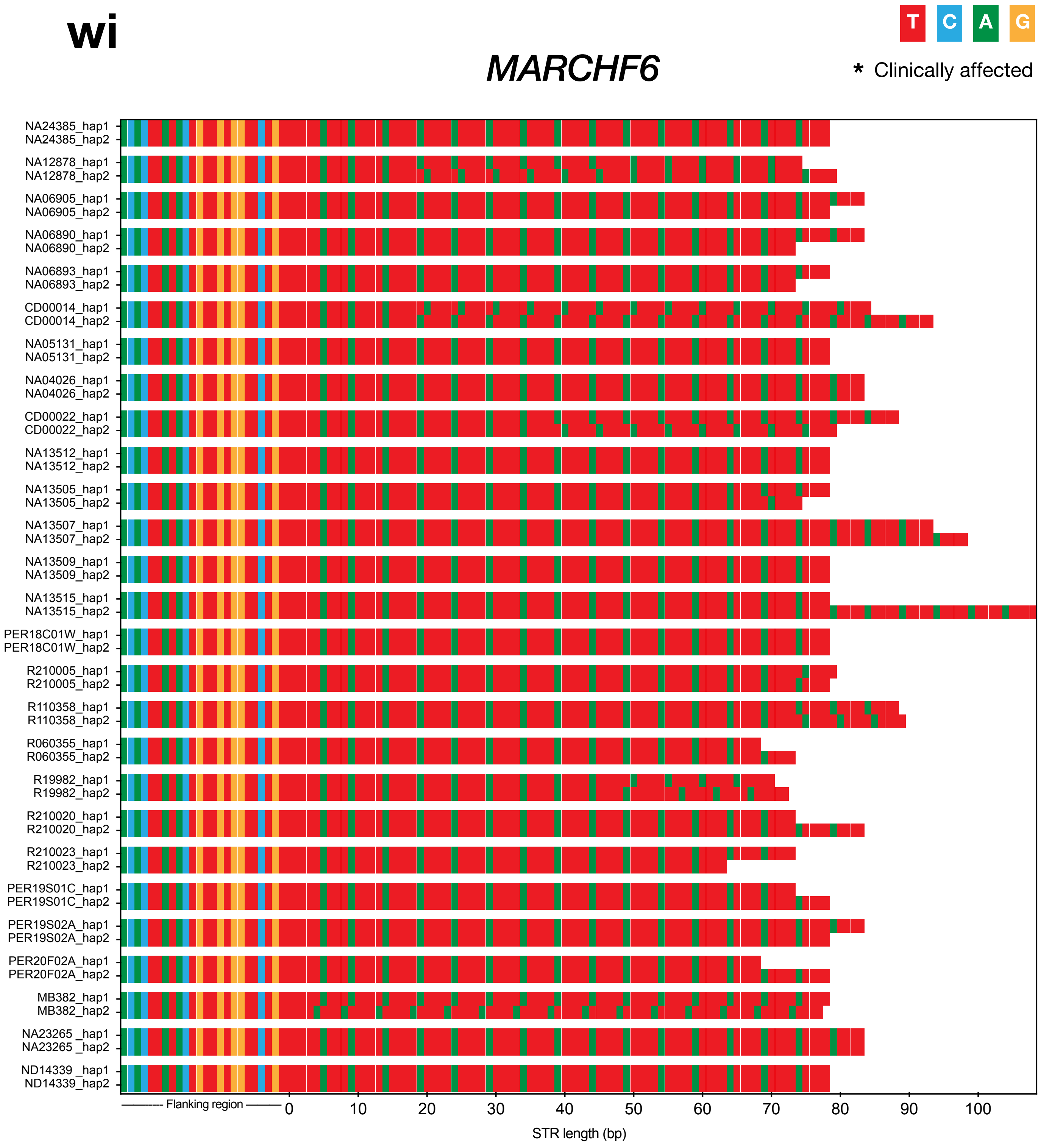


**READ DEPTH**
Range: 6-40x
Median: 18x

**REPEAT LENGTHS (#)**
Min: 12
Max: 21

**SEQUENCE**
Motifs Seen: TTTTA
Interruptions Seen: TTTTTA

xi

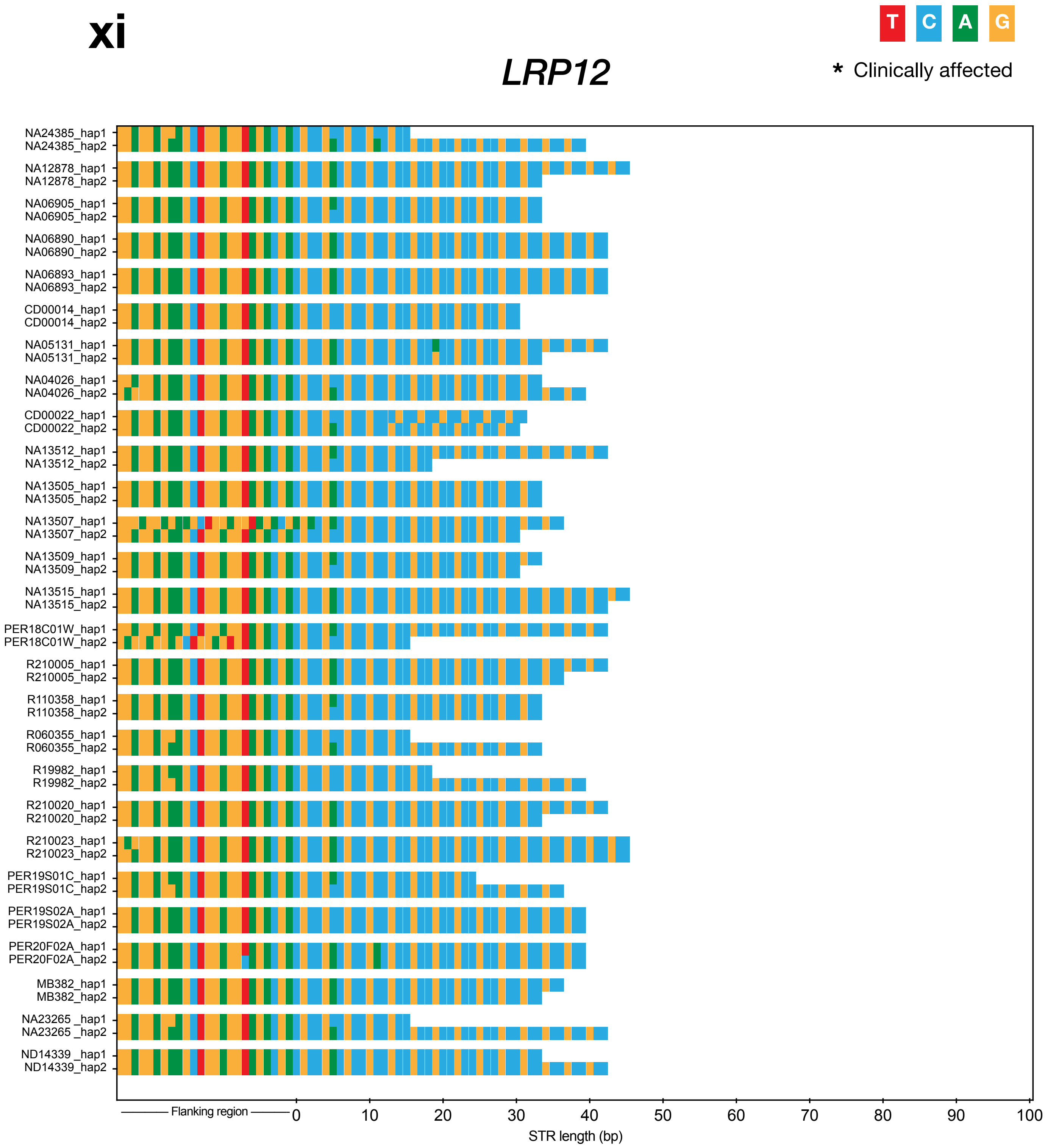


**READ DEPTH**
Range: 8-41x
Median: 19x

**REPEAT LENGTHS (#)**
Min: 5
Max: 15

**SEQUENCE**
Motifs Seen: CCG
Interruptions Seen: ACG, CCA

**yi**

*JPH3*

T C A G

* Clinically affected

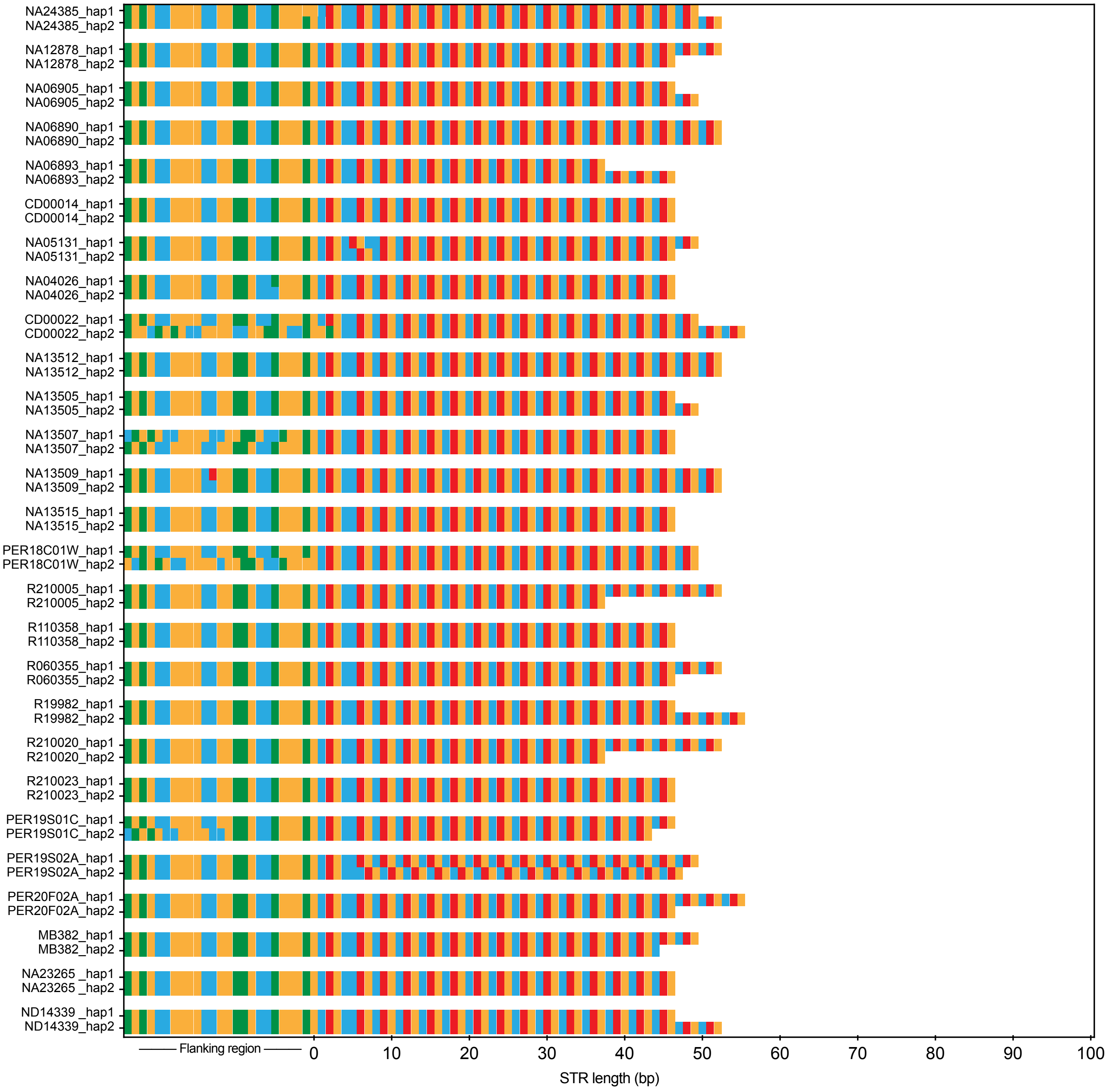


**READ DEPTH**
Range: 8-30x
Median: 18x

**REPEAT LENGTHS (#)**
Min: 12
Max: 18

**SEQUENCE**
Motifs Seen: CTG
Interruptions Seen:

zi

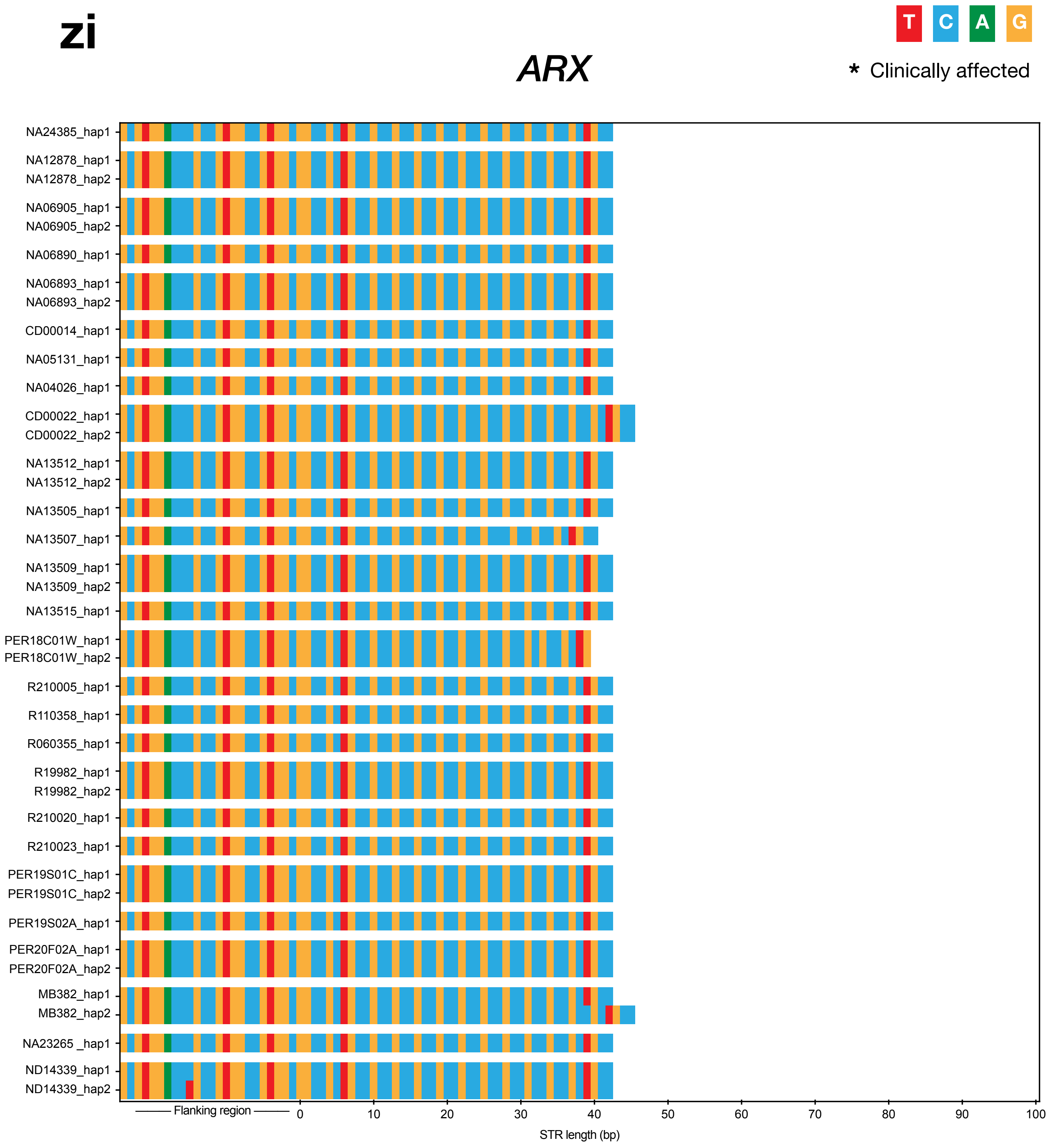


**READ DEPTH**
Range: 4-42x
Median:14x

**REPEAT LENGTHS (#)**
Min: 12
Max: 15

**SEQUENCE**
Motifs Seen: CCG
Interruptions Seen: CTG

**aii**

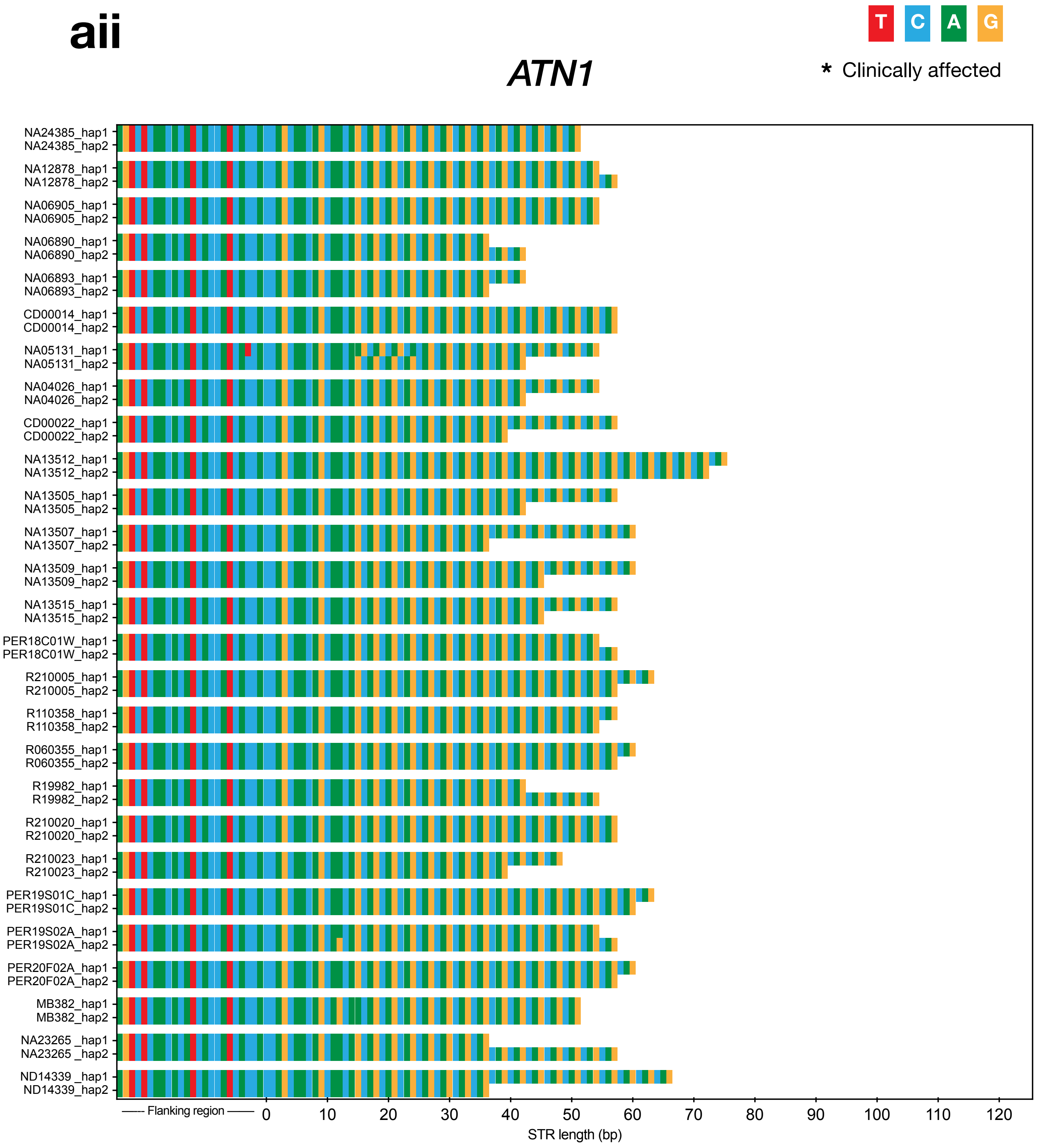


**READ DEPTH**
Range: 6-32x
Median:19x

**REPEAT LENGTHS (#)**
Min: 12
Max: 25

**SEQUENCE**
Motifs Seen: CAG
Interruptions Seen: CAA

#### bii

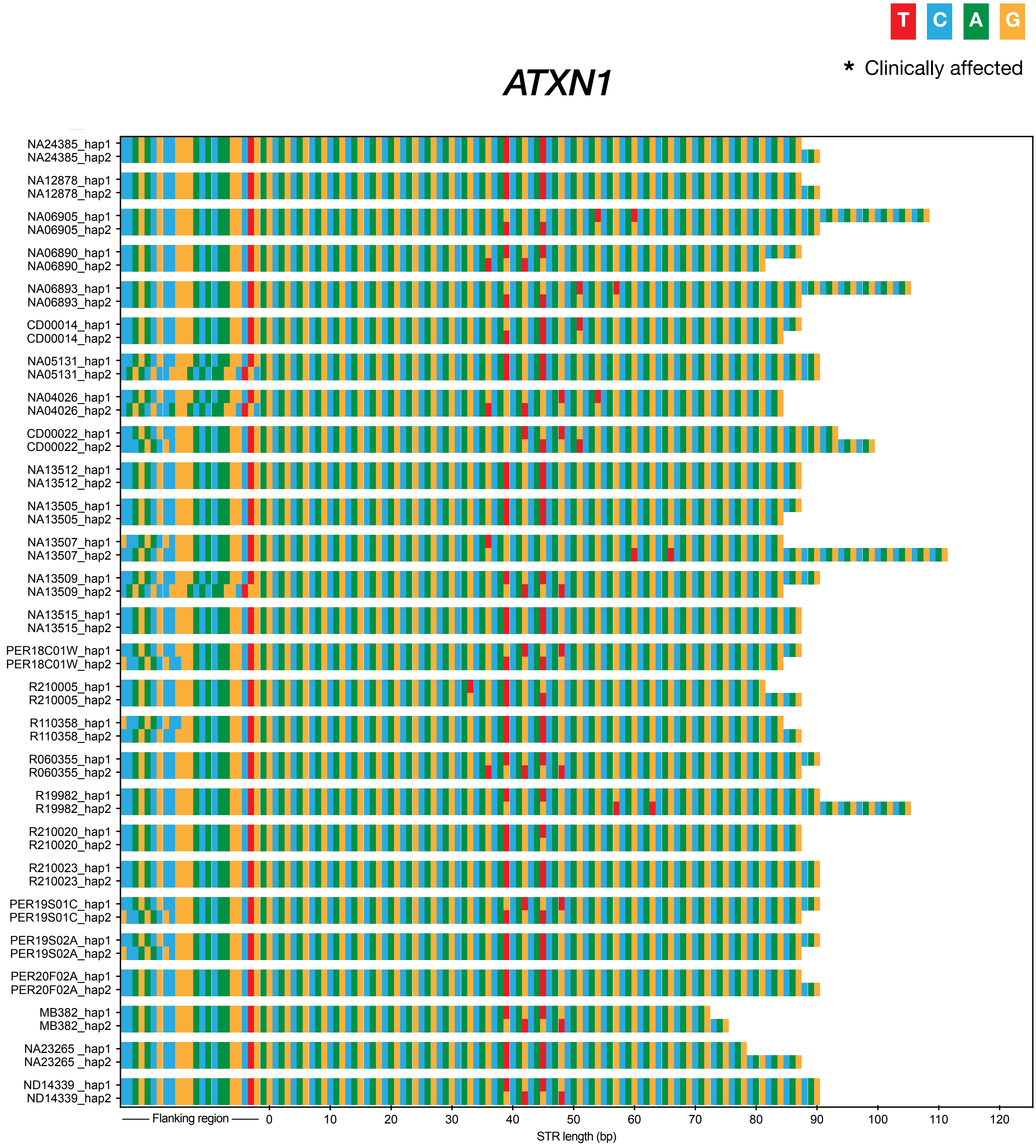
T C A G
* Clinically affected
ATXN1
NA24385_hap1
NA24385_hap2
NA12878_hap1
NA12878_hap2
NA06905_hap1
NA06905_hap2
NA06890_hap1
NA06890_hap2
NA06893_hap1
NA06893_hap2
CD00014_hap1
CD00014_hap2
NA05131_hap1
NA05131_hap2
NA04026_hap1
NA04026_hap2
CD00022_hap1
CD00022_hap2
NA13512_hap1
NA13512_hap2
NA13505_hap1
NA13505_hap2
NA13507_hap1
NA13507_hap2
NA13509_hap1
NA13509_hap2
NA13515_hap1
NA13515_hap2
PER18C01W_hap1
PER18C01W_hap2
R210005_hap1
R210005_hap2
R110358_hap1
R110358_hap2
R060355_hap1
R060355_hap2
R19982_hap1
R19982_hap2
R210020_hap1
R210020_hap2
R210023_hap1
R210023_hap2
PER19S01C_hap1
PER19S01C_hap2
PER19S02A_hap1
PER19S02A_hap2
PER20F02A_hap1
PER20F02A_hap2
MB382_hap1
MB382_hap2
NA23265 _hap1
NA23265 _hap2
ND14339 _hap1
ND14339_hap2
Flanking region
0
10
20
30
40
50
60
70
80
90
100
110
120
STR length (bp)

**cii**

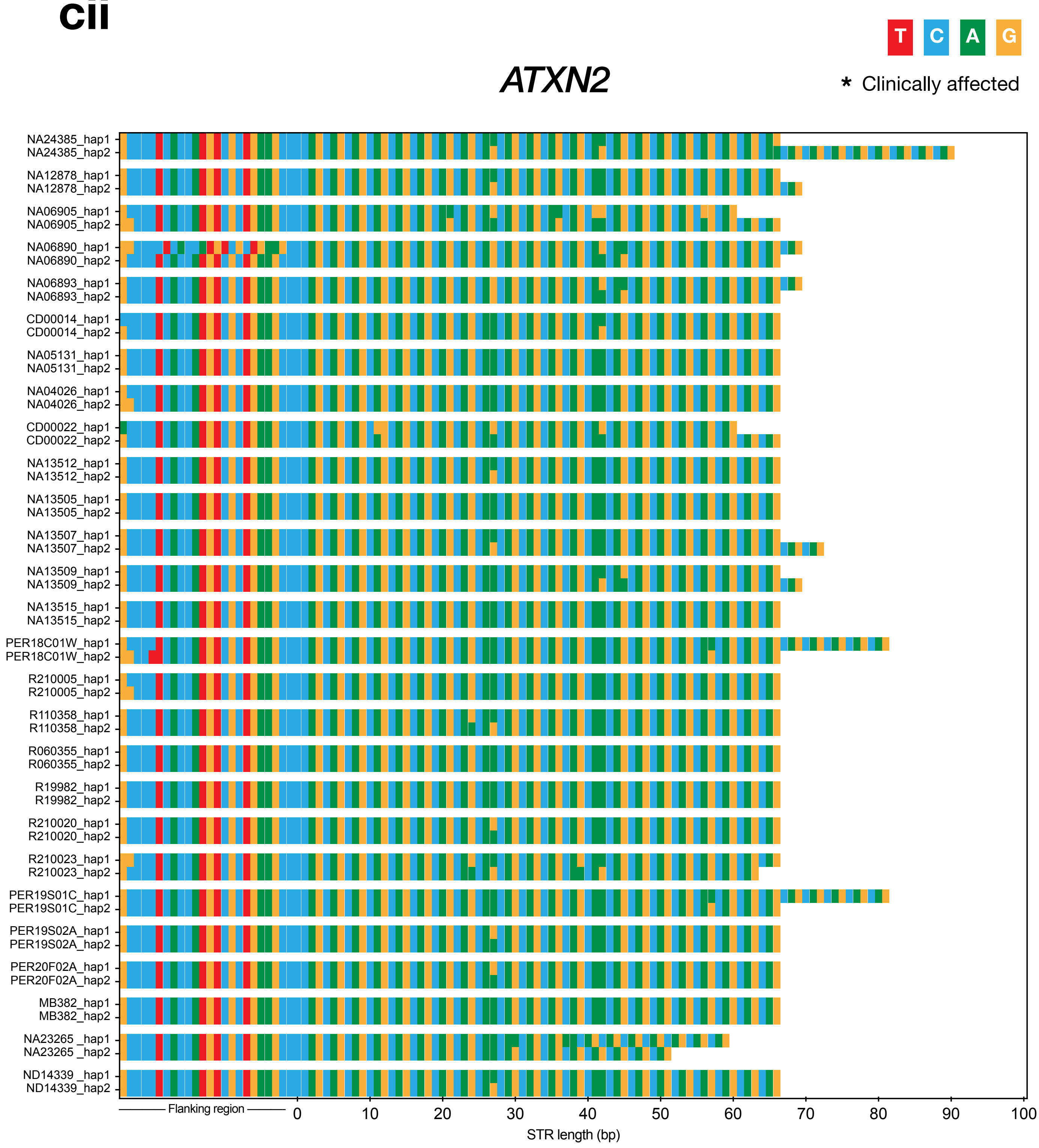


**READ DEPTH**
Range: 8-46x
Median:19x

**REPEAT LENGTHS (#)**
Min: 17
Max: 30

**SEQUENCE**
Motifs Seen: CAG
Interruptions Seen: CAA

**dii**

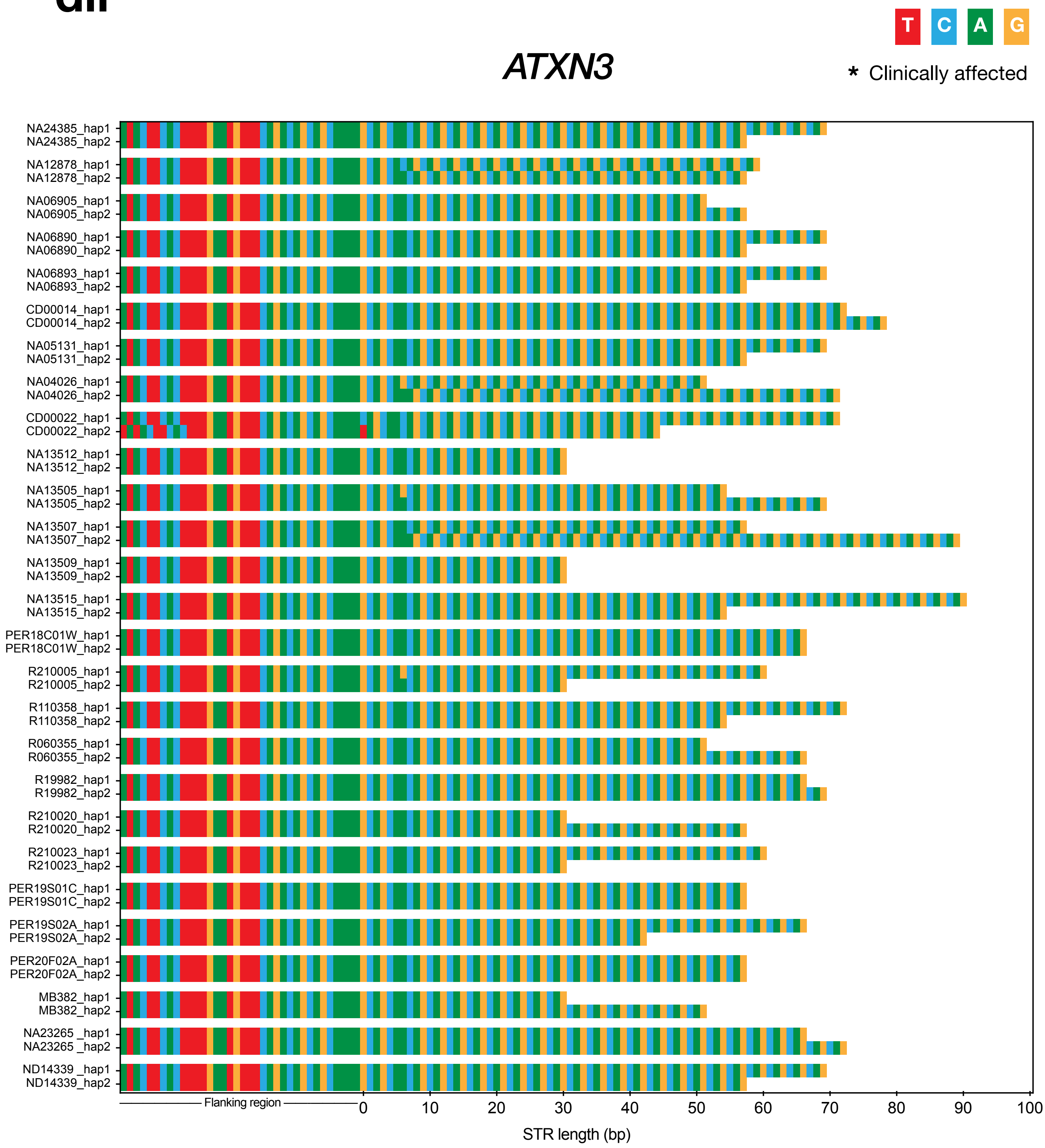


**READ DEPTH**
Range: 8-47x
Median:18x

**REPEAT LENGTHS (#)**
Min: 10
Max: 30

**SEQUENCE**
Motifs Seen: CAG
Interruptions Seen: CAA

**eii**

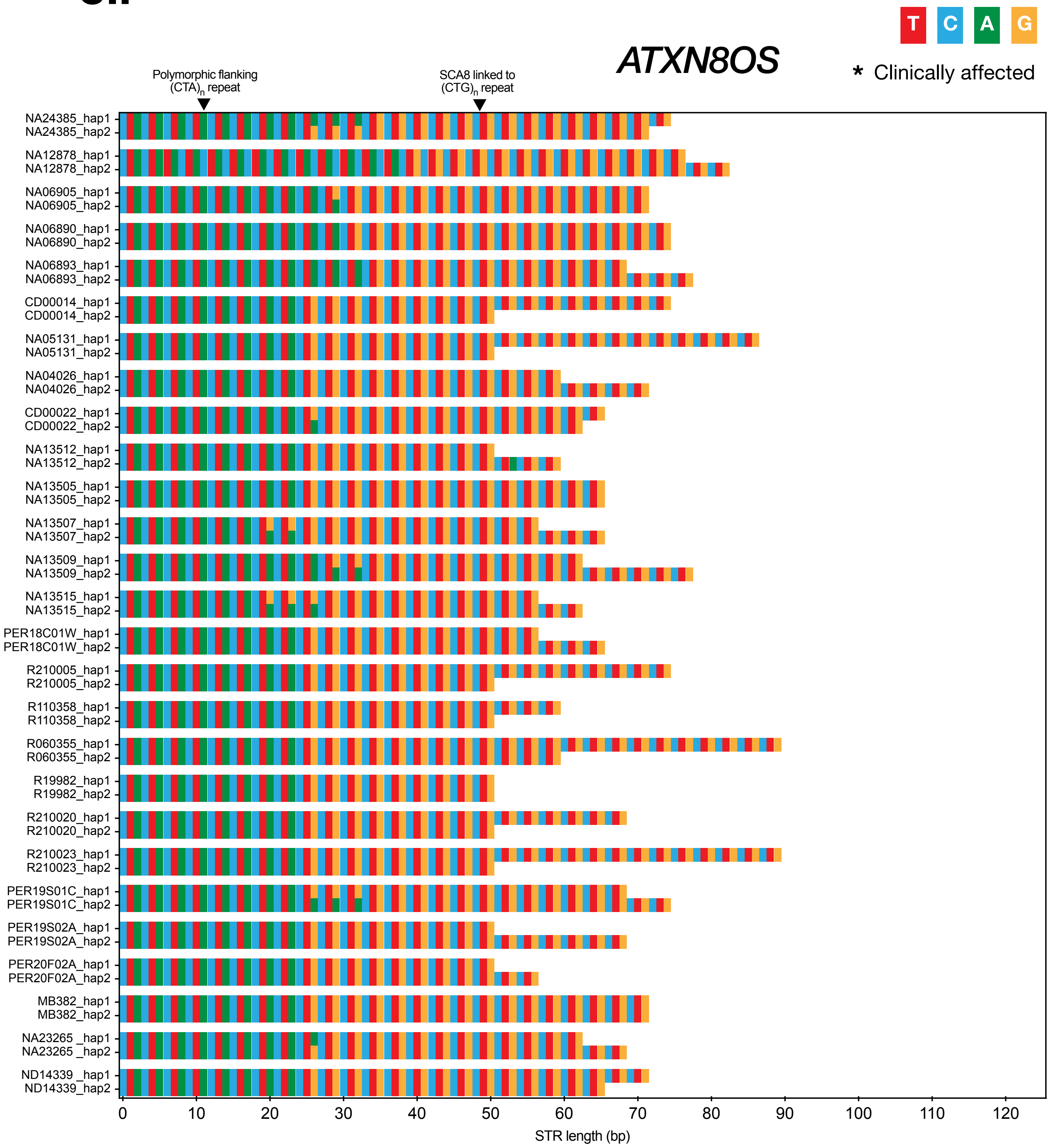


**READ DEPTH**
Range: 5-38x
Median:18x

**REPEAT LENGTHS (#)**
Min: 9
Max: 22

**SEQUENCE**
Motifs Seen: CTG
Interruptions Seen: CTA

**fii**

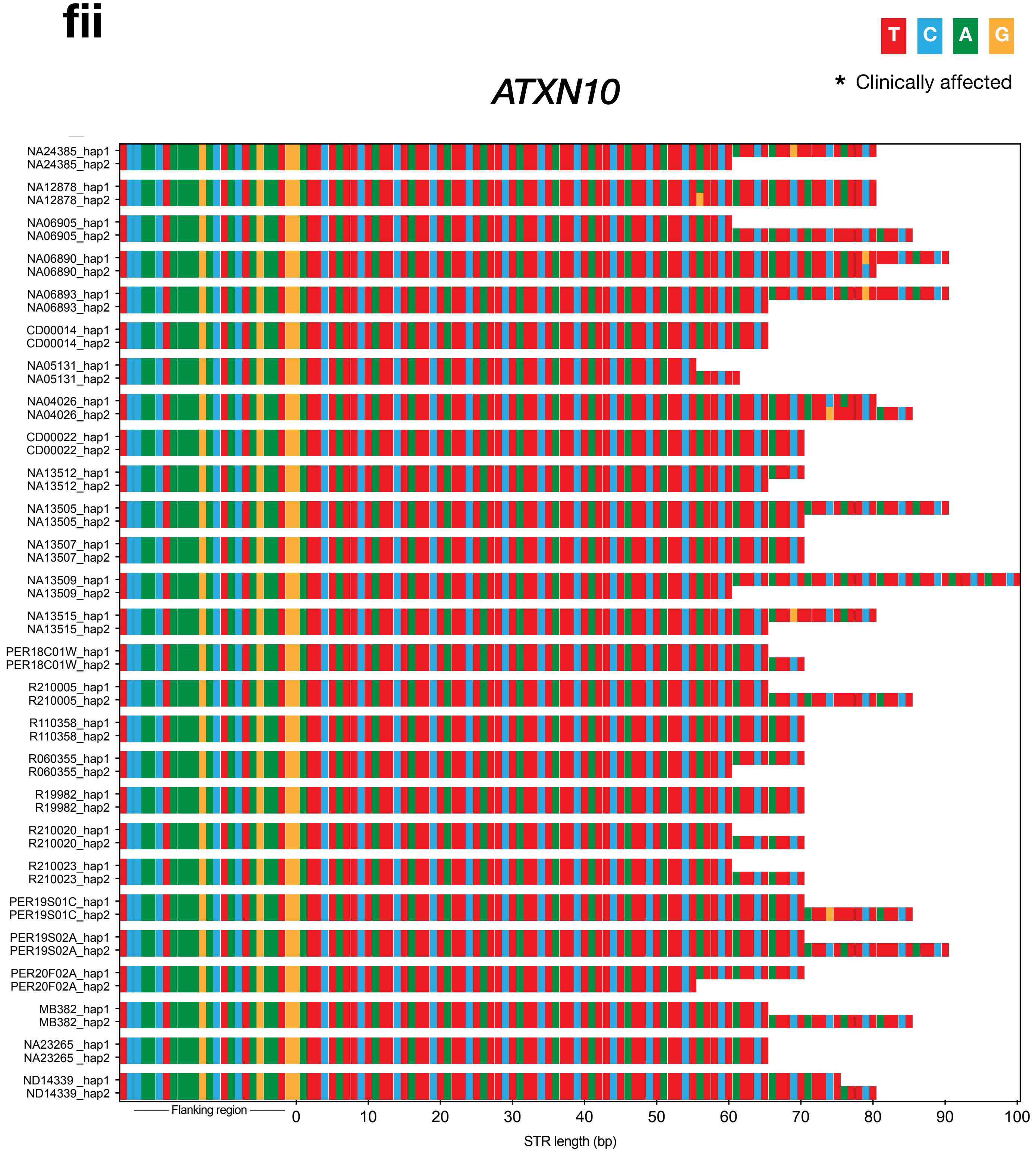


**READ DEPTH**
Range: 7-48x
Median: 19x

**REPEAT LENGTHS (#)**
Min: 11
Max: 20

**SEQUENCE**
Motifs Seen: ATTCT
Interruptions Seen: GTTCT, ATTGT, TTTCT

**gii**

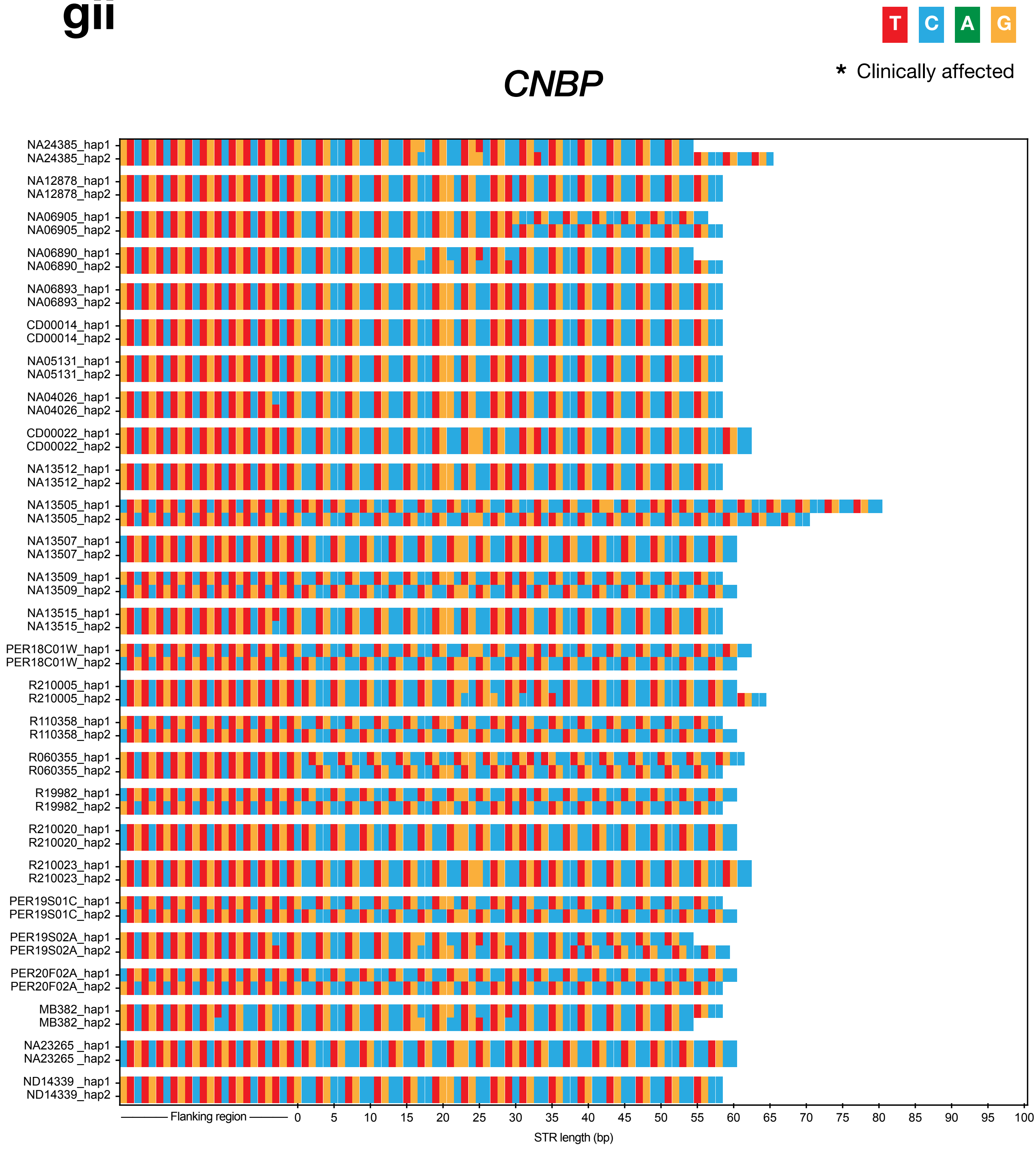


**READ DEPTH**
Range: 8-37x
Median: 19x

**REPEAT LENGTHS (#)**
Min: 14
Max: 19

**SEQUENCE**
Motifs Seen: CCTG
Interruptions Seen: GCTG, TCTG

hii

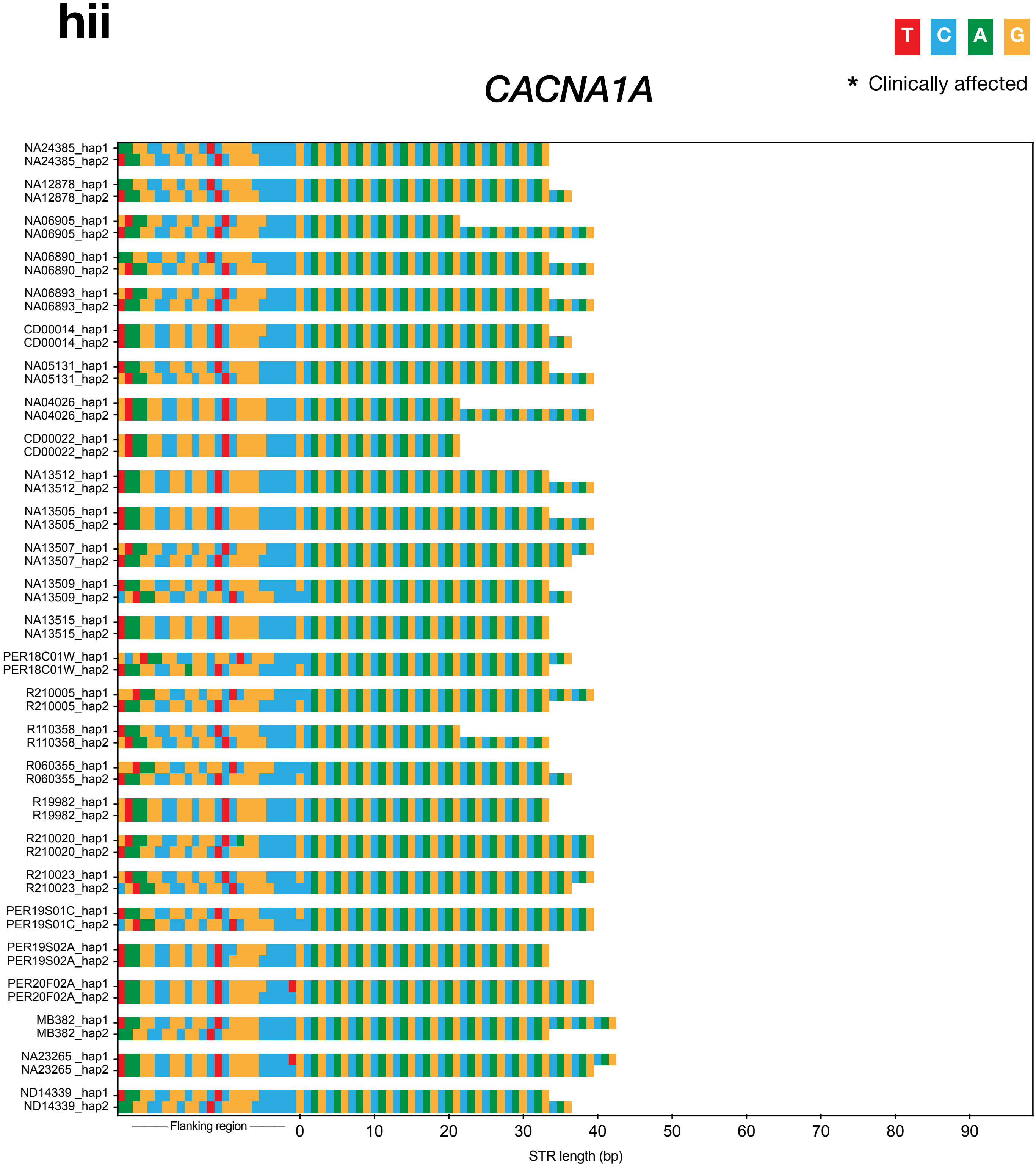


**READ DEPTH**
Range: 4-35x
Median: 22x

**REPEAT LENGTHS (#)**
Min: 7
Max: 14

**SEQUENCE**
Motifs Seen: CAG
Interruptions Seen:

iii

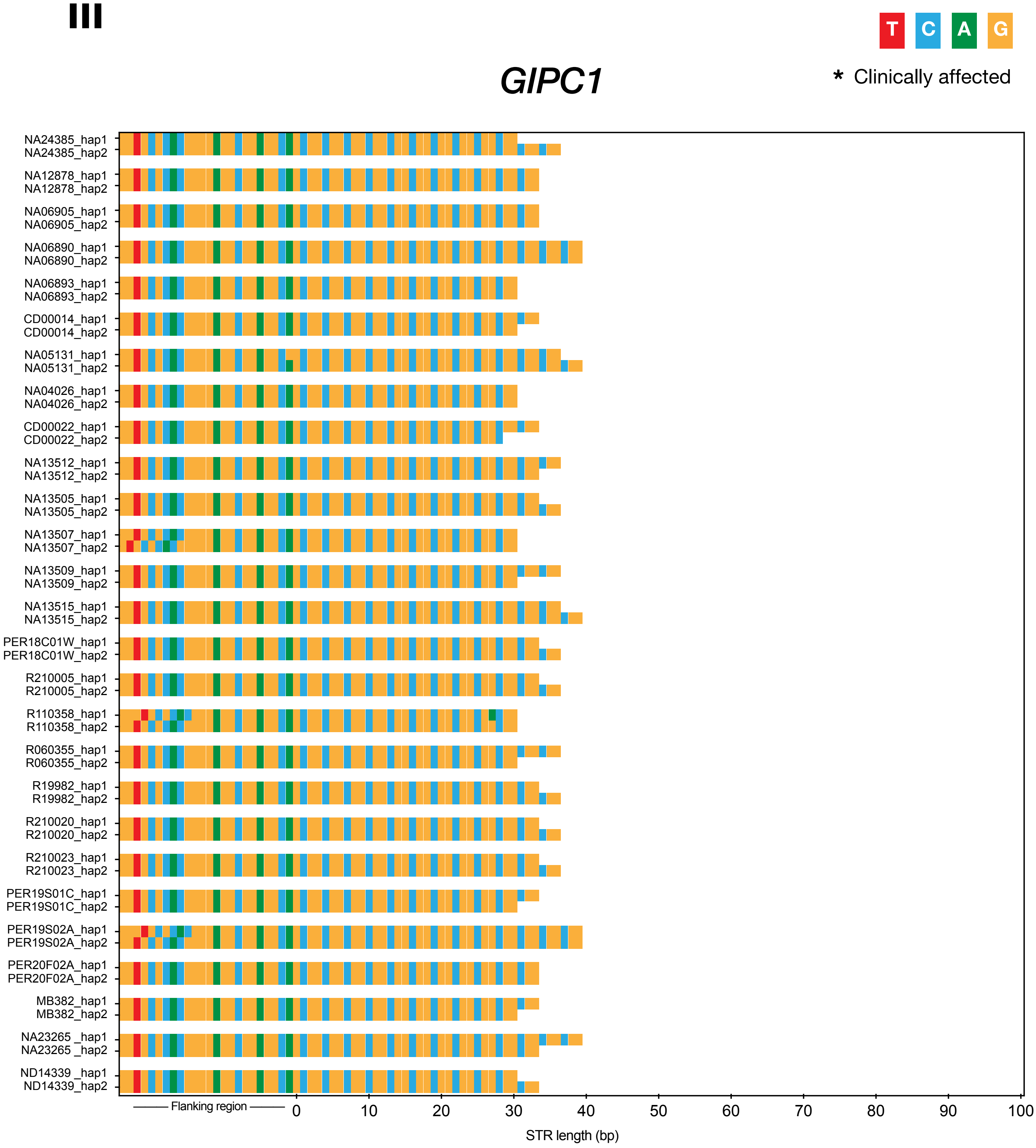


**READ DEPTH**
Range: 7-37x
Median: 21x

**REPEAT LENGTHS (#)**
Min: 10
Max: 13

**SEQUENCE**
Motifs Seen: GGC
Interruptions Seen: CGA

**jii**

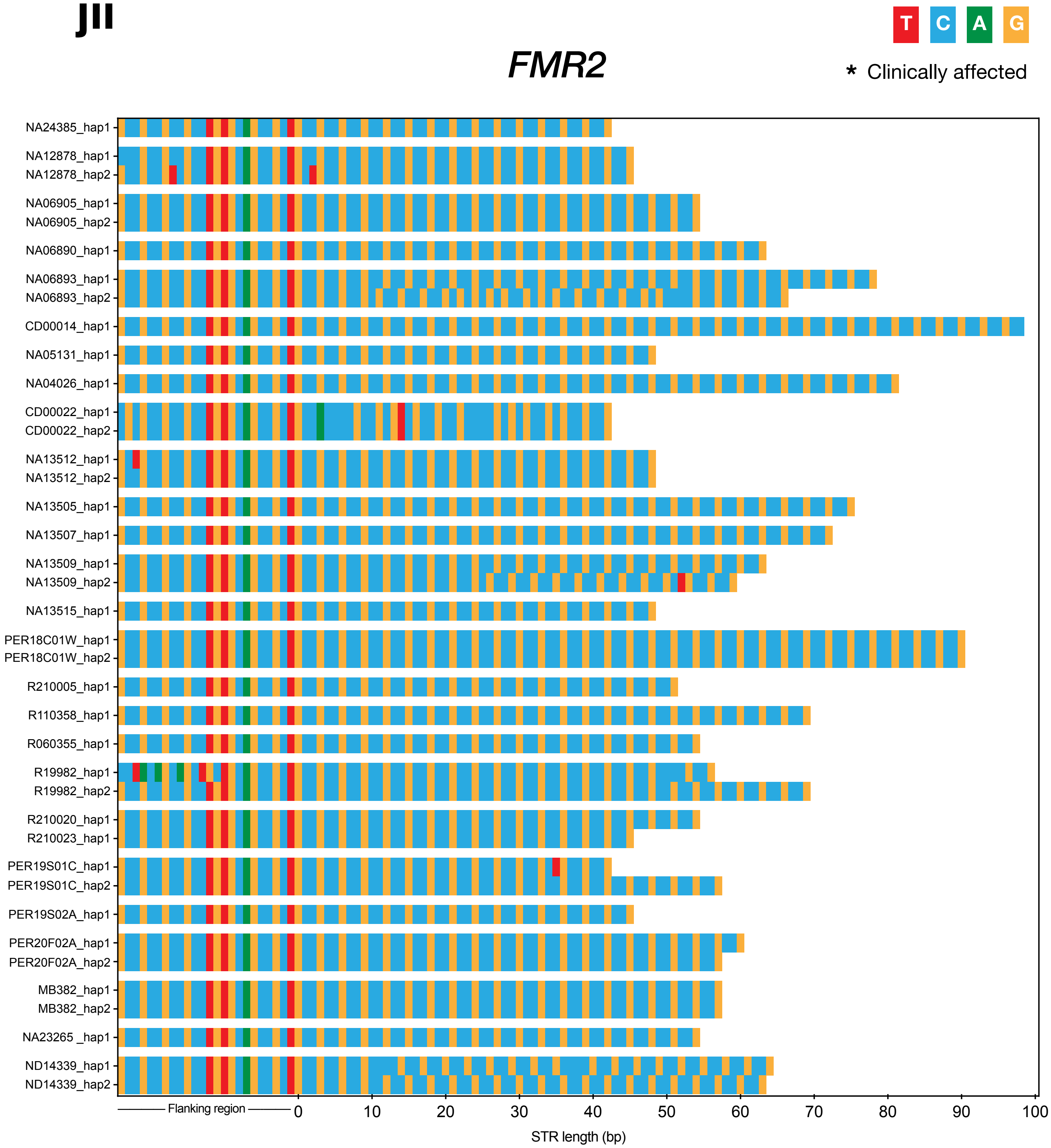


**READ DEPTH**
Range: 5-30x
Median: 11x

**REPEAT LENGTHS (#)**
Min: 13
Max: 33

**SEQUENCE**
Motifs Seen: CCG
Interruptions Seen: CTG
